# Proteomic Signatures and an Injury-Stress Endotype in Myositis-Associated Interstitial Lung Disease

**DOI:** 10.64898/2026.08.04.26359441

**Authors:** Julio A. Huapaya, Peter D. Burbelo, Eric W. Robbins, Xin Tian, Shijinqiu Gao, Sevilay Turan, Salina Gairhe, James M. Ward, Neelam Redekar, Jianliang Li, Gloria Pastor, Neha A. Gupta, Payam Noroozi Farhadi, Kakali Sarkar, Maria Casal-Dominguez, Iago Pinal-Fernandez, Lisa Christopher-Stine, Adam Schiffenbauer, Lisa G. Rider, Andrew L. Mammen, Sonye K. Danoff, Anthony F. Suffredini

## Abstract

**Rationale:** Idiopathic inflammatory myopathy-associated interstitial lung disease (IIM-ILD) is a major cause of morbidity and mortality. We tested whether quantitative myositis-specific autoantibodies and proteomic profiling capture biological heterogeneity and prognosis beyond categorical serology.

**Methods:** Myositis-specific autoantibodies were quantified using the luciferase immunoprecipitation systems assay, and 184 serum proteins were measured in 226 IIM patients; 199 with higher-ILD-risk autoantibodies (Jo-1/MDA5/PL-7/PL-12/EJ), 27 with lower- ILD-risk autoantibodies (Mi-2/NXP2/TIF1γ) and 35 healthy controls. We identified shared and subgroup-specific differences by comparing each subgroup with controls, then correlated quantitative autoantibody and protein levels within higher-risk subgroups. Additional analyses included pathway enrichment, unsupervised clustering, longitudinal lung-function change, and mortality.

**Results:** Higher-ILD-risk subgroups shared interferon-responsive CXCR3 chemokine, IL- 6/JAK/STAT3, and apoptosis signaling. Dominant autoantibody subgroup profiles differed: interferon/CXCR3 chemokine signaling with T-cell activation and monocyte recruitment in anti-Jo-1; proteostasis/antigen-processing and vascular/cellular stress signals in anti-MDA5; IL- 6/macrophage and profibrotic signals in anti-PL-12; and apoptotic and innate immune activation with metabolic/redox-stress signals in anti-PL-7. Within higher-ILD-risk subgroups, autoantibody levels correlated with interferon-response, profibrotic, and metabolic/vascular proteins (r=0.40- 0.74; nominal p<0.05). Unsupervised clustering identified four proteomic endotypes beyond autoantibody type, including an injury-stress endotype associated with worse lung function and poorer survival, and a chemokine/checkpoint-high endotype with relatively preserved lung function. Across 203 participants with 38 deaths, a weighted 10-protein score was associated with all-cause mortality (HR, 3.28; 95% CI, 2.12-5.08; p<0.001).

**Conclusions:** Integrated quantitative autoantibodies and proteomic profiling revealed shared inflammatory biology, autoantibody-associated signatures, and an injury-stress endotype associated with poor survival in IIM-ILD, supporting risk stratification beyond categorical serology.

## Introduction

Idiopathic inflammatory myopathy-associated interstitial lung disease (IIM-ILD) is a major driver of morbidity and mortality in patients with myositis(1–4). Unlike idiopathic pulmonary fibrosis (IPF), IIM-ILD often affects younger individuals(5) and may stabilize or improve with immunosuppressive therapy(1–4, 6). However, a substantial proportion of patients develop progressive pulmonary fibrosis despite treatment(7, 8). A critical gap remains in identifying patients at highest risk for progression before irreversible lung injury occurs.

Myositis-specific autoantibodies are central to IIM-ILD classification and define distinct clinical phenotypes(6, 9, 10). Anti-Jo-1, anti-PL-7, anti-PL-12, and anti-MDA5 are strongly associated with ILD(5, 10, 11), whereas anti-Mi-2, anti-NXP2, and anti-TIF1γ are less strongly associated with ILD(11–13). However, substantial heterogeneity remains within autoantibody-defined groups, suggesting that categorical serology alone does not fully capture disease biology, severity, or prognosis. Although translational studies suggest that some myositis autoantibodies may contribute to tissue-specific injury(14–17), most clinical assays report autoantibodies categorically and vary in diagnostic and quantitative performance(18, 19). The luciferase immunoprecipitation systems (LIPS) assay provides quantitative measurements across a broad dynamic range(20, 21), enabling assessment of whether autoantibody levels relate to downstream circulating protein patterns.

Proteomic studies have identified inflammatory pathways, prognostic proteins, and molecular endotypes across multiple diseases including ILD and myositis cohorts(22–38). However, the relationship between quantitative myositis autoantibody levels and circulating protein profiles has not been systematically defined across multiple adult ILD-risk subgroups or linked to lung function and survival. We therefore integrated LIPS-based quantitative autoantibody profiling with targeted serum proteomics in an IIM-ILD-enriched cohort to identify shared and subgroup-associated signatures, proteomic endotypes and outcome-related protein patterns.

## Methods

### Study Cohort and biomarkers measurements

We analyzed 264 serum samples from 261 individuals, including 226 adults with IIM and 35 age-, sex-, and race-matched healthy controls recruited from the Johns Hopkins Myositis Center, the National Institute of Environmental Health Sciences, and the National Institute of Arthritis and Musculoskeletal and Skin Diseases between 2010 and 2020. IIM diagnoses were confirmed by the treating specialists. All patients fulfilled the 2017 EULAR/ACR classification criteria(39). Patients were categorized by autoantibody group into higher-ILD-risk groups (i.e., Jo-1, MDA5, PL-7, PL-12, EJ) and lower-ILD-risk groups (Mi-2, NXP2, TIF1γ)(13). ILD was defined by high-resolution CT abnormalities and/or pulmonary function test showing restriction with impaired gas transfer. Institutional review boards approved the study, and all participants provided written informed consent.

### Targeted proteomics with proximity extension assay (PEA)

Serum levels of 184 proteins were measured using the Olink Inflammation and Organ Damage proximity-extension assay panels (Supplementary Table S1). Myositis-specific autoantibodies were quantified using the luciferase immunoprecipitation systems assay as previously described (20, 21). Additional cohort, assay, quality-control, and protein-nomenclature details are provided in the Supplementary Material.

### Statistical Analysis

Differentially expressed proteins were identified using linear models after ComBat batch correction, with significance defined by a 5% false discovery rate and an absolute mean fold-change threshold of 1.25. Spearman correlations were calculated between continuous quantitative LIPS autoantibody levels and protein levels. Pathway enrichment was evaluated using MSigDB Hallmark gene sets(40) and Ingenuity Pathway Analysis (Qiagen, Germantown, MD)(41)

Proteomic endotypes were identified by k-means clustering of scaled, batch-corrected baseline protein data, with the number of clusters selected using the elbow method. Uniform Manifold Approximation and Projection (UMAP) was used for visualization only.

Mortality-associated proteins were prioritized using adjusted Cox proportional hazards and random survival forest models. Candidate 5- and 10-protein scores were evaluated after adjustment for age, sex, and ILD status, with additional adjustment for baseline forced vital capacity (FVC%) predicted. Hazard ratios were reported per 1-SD increase, and model discrimination was internally validated using 1,000 bootstrap resamples with correction for optimism. Exploratory random forest and LASSO models evaluated whether baseline protein profiles predicted subsequent lung-function decline or improvement at 6–24 months; outcome definitions and modeling details are provided in the online supplement(42, 43). All analyses were performed using R version 4.6.0 (R Foundation for Statistical Computing) . Additional statistical details are provided in the Supplementary Material.

## Results

### Cohort and global proteomic differences

The cohort included 199 patients with higher-ILD-risk autoantibodies, 27 patients with lower- ILD-risk autoantibodies, and 35 age-, sex-, and race-matched healthy controls. Baseline demographic, clinical, pulmonary, laboratory, and treatment characteristics stratified by autoantibody group are shown in Table 1. Full characteristics are provided in Supplementary Table S2. Of the 264 samples assayed from 261 participants, proteomic data were retained after quality control for 250 samples (94.7%) on the Inflammation panel and 246 samples (93.2%) on the Organ Damage panel.

**Table 1.**
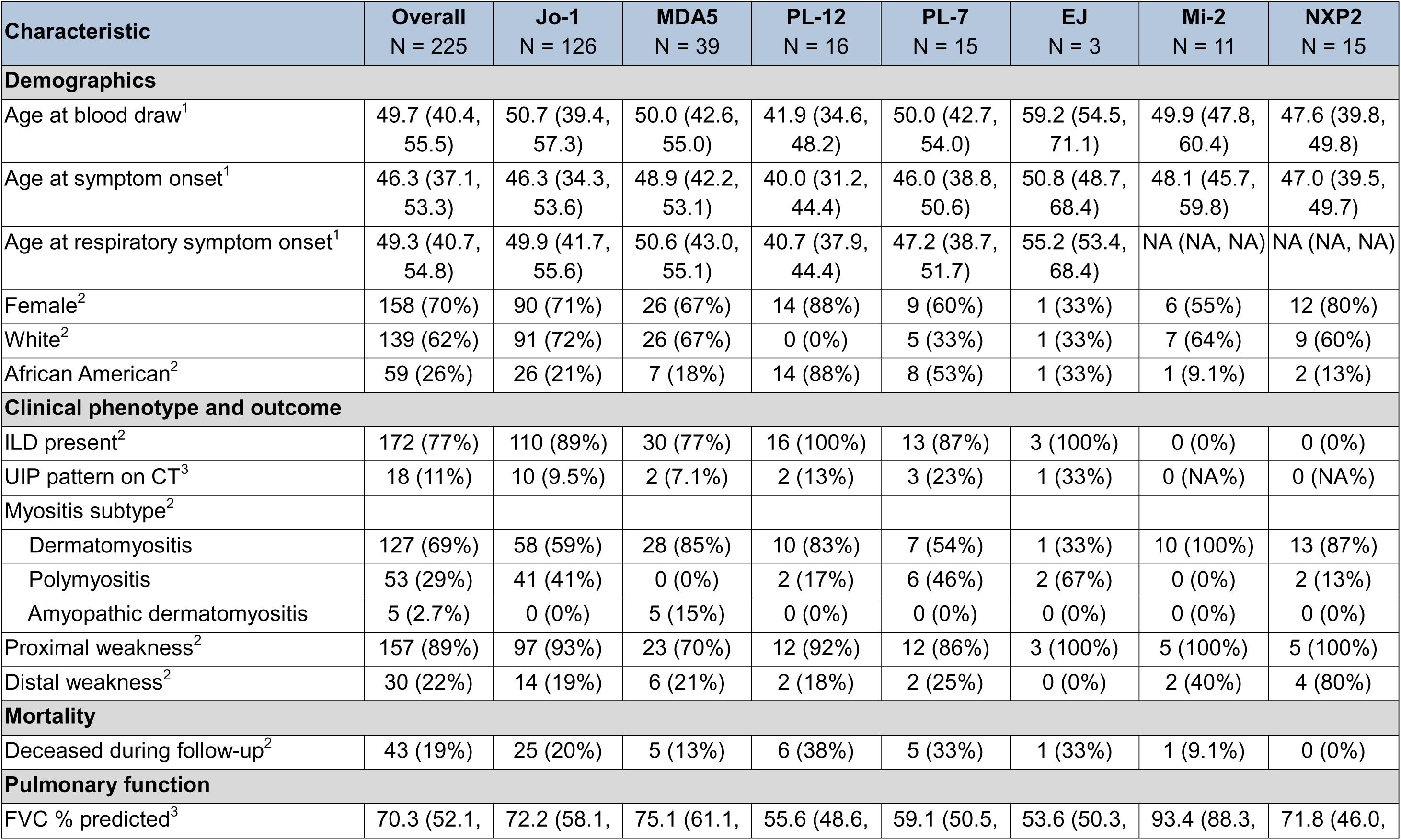
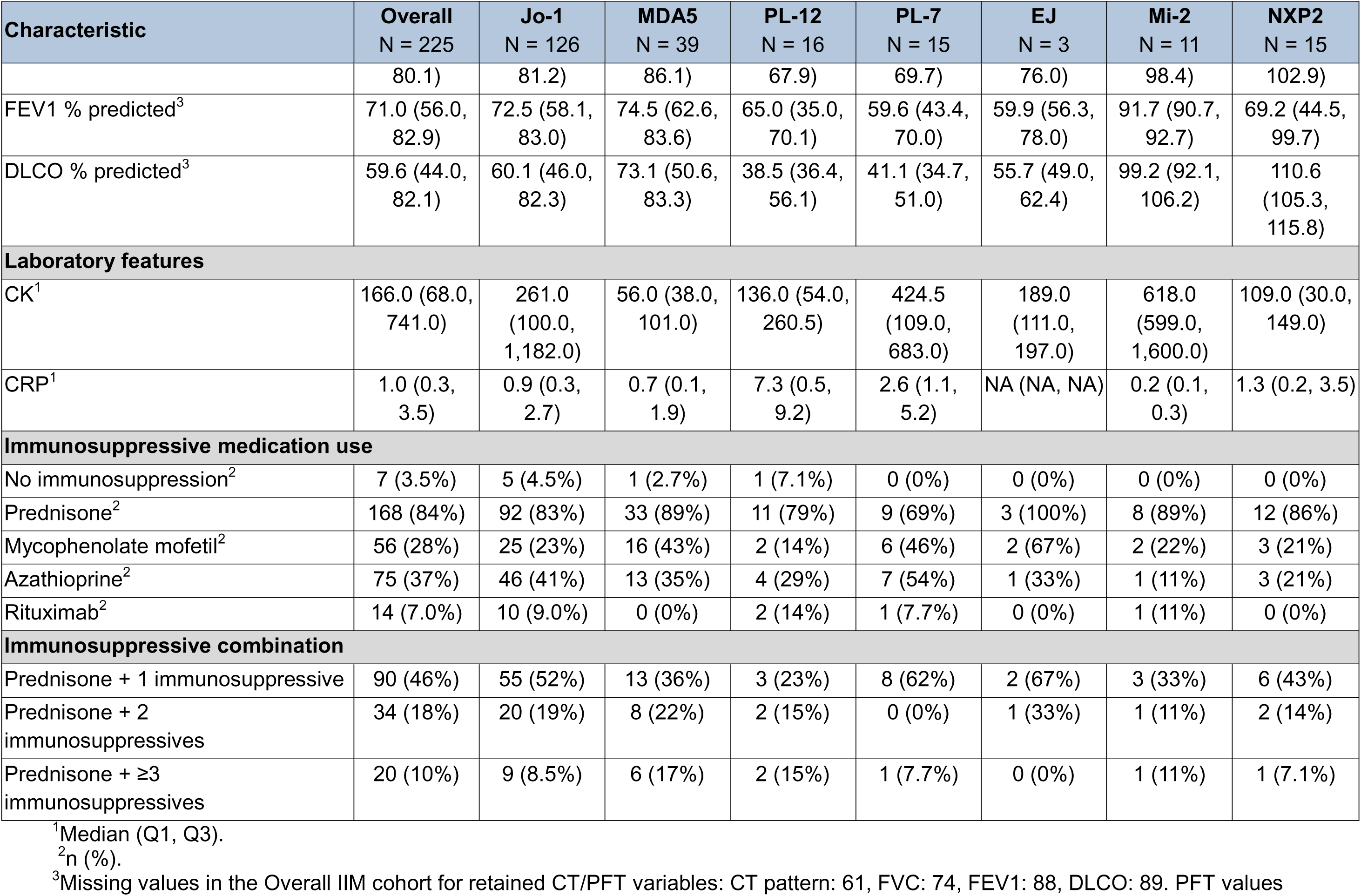
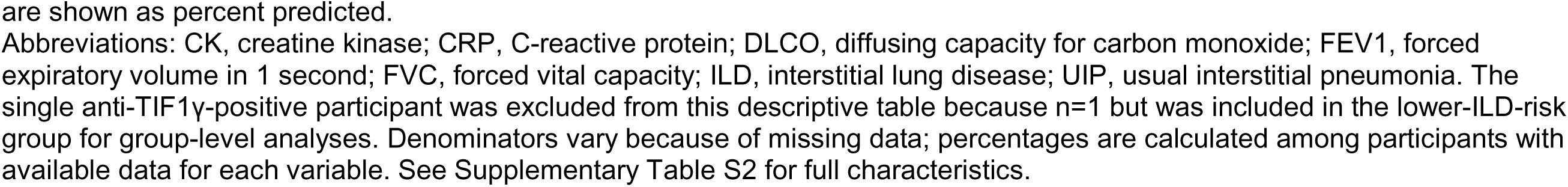
Selected clinical characteristics and outcomes stratified by autoantibody group.

Patients with higher-ILD-risk autoantibodies (i.e., Jo-1, MDA5, PL-7, PL-12, and EJ) had 88 differentially expressed proteins compared with healthy controls (69 upregulated and 19 downregulated; Figure 1A). Direct comparison with the lower-ILD-risk group identified 31 differentially expressed proteins (30 upregulated and 1 downregulated; Figure 1B). Patients with lower-ILD-risk autoantibodies (i.e., Mi-2, NXP2, and TIF1γ) had 70 differentially expressed proteins compared with controls (41 upregulated and 29 downregulated; Supplementary Figure S1).

**Figure 1.**
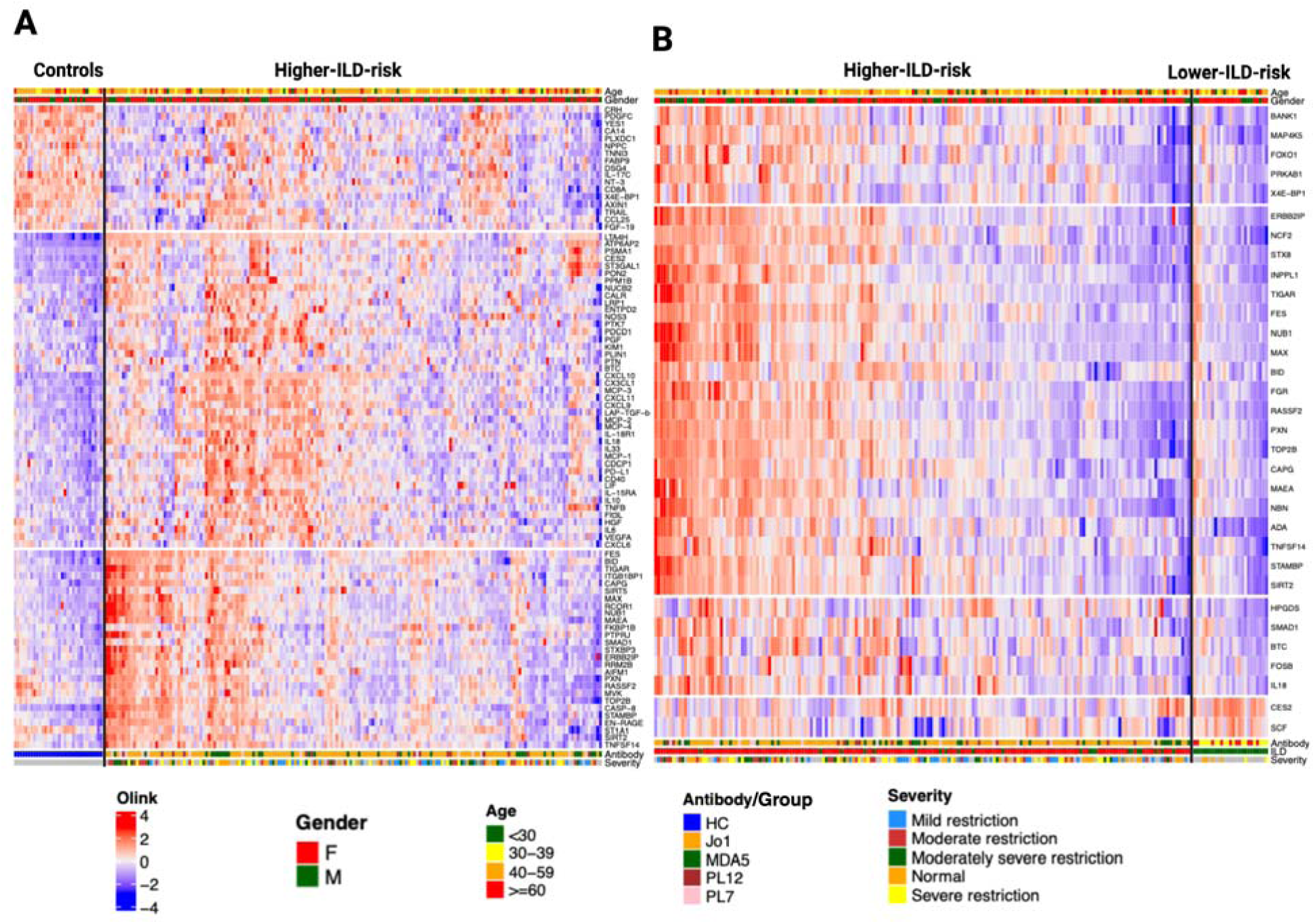
Differentially expressed proteins in higher-ILD-risk autoantibody groups compared with controls and lower-ILD-risk autoantibody groups. (A) Heatmap of differentially expressed serum proteins comparing patients with higher-ILD-risk autoantibodies with matched healthy controls. Columns represent individual participants and rows represent proteins identified as differentially expressed in the group-level comparison. Protein abundance is shown as normalized Olink NPX values scaled by protein, with higher relative expression shown in red and lower relative expression shown in blue. Column annotations indicate age group, sex, autoantibody type, and ILD severity. Higher-ILD-risk autoantibody groups included anti-Jo-1, anti-MDA5, anti-PL-12, and anti-PL-7. The EJ participant was included in the corresponding group-level analyses but is not displayed separately due to the small sample size. (B) Heatmap of differentially expressed serum proteins comparing patients with higher-ILD-risk versus lower-ILD-risk autoantibodies. Columns represent individual participants and rows represent proteins identified as differentially expressed in this comparison. Column annotations indicate age group, sex, autoantibody type, ILD status, and ILD severity. Lower-ILD-risk autoantibody groups included anti-Mi-2 and anti-NXP2. The single anti-TIF1γ-positive participant was included in the lower-ILD-risk group-level comparison but is not displayed separately because n=1.

Across higher-ILD-risk autoantibody subgroups, we observed a shared proteomic signature characterized by increased caspase-8 (CASP8), leukotriene A4 hydrolase (LTA4H), interferon-inducible chemokines CXCL9, CXCL10, and CXCL11, monocyte chemoattractant proteins MCP-3/CCL7, MCP-2/CCL8, and MCP-4/CCL13, and inflammatory/remodeling mediators including IL-6, IL-10, lymphotoxin-alpha/TNF-β (TNFB/LTA), IL-33, LAP-TGF-β1, VEGFA, and HGF when compared to controls. Additional upregulated proteins included CES2, FES, and ATP6AP2. The most consistently downregulated proteins included cardiac troponin I (TNNI3), corticotropin-releasing hormone (CRH), eIF4E-binding protein 1/4E-BP1 (EIF4EBP1), C-type natriuretic peptide (NPPC), CD8 alpha chain (CD8A), YES1, IL-17C, AXIN1, PDGFC, and neurotrophin-3 (NT-3/NTF3). Representative proteins illustrating the shared higher-ILD-risk signature are shown in Figure 2A. Within this shared interferon-responsive signature, CXCL9 is more closely associated with IFN-γ/type II interferon signaling, whereas CXCL10 and CXCL11 may reflect both type I and type II interferon activity.

**Figure 2.**
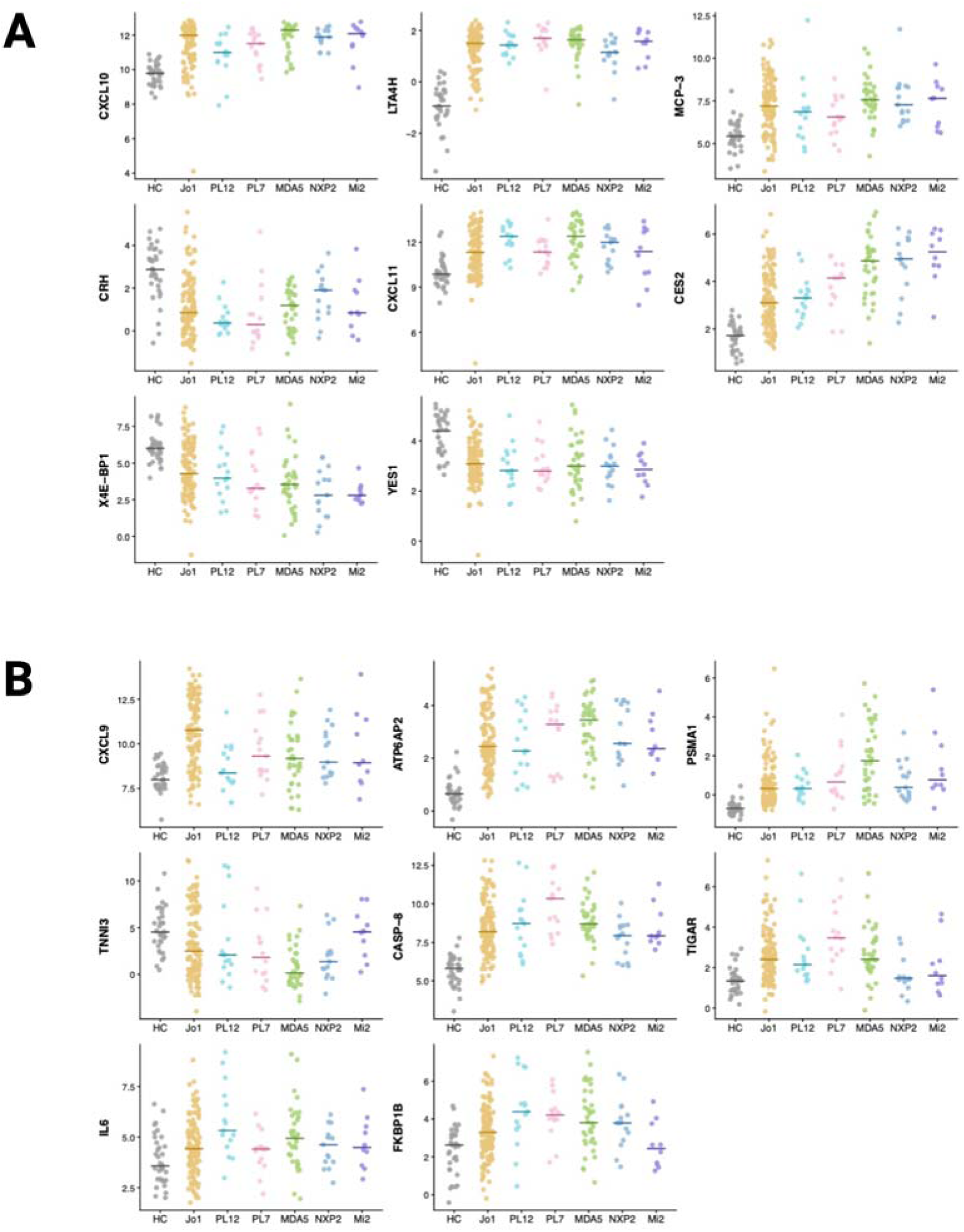
Shared and autoantibody-associated protein expression patterns across myositis autoantibody subgroups. (A) Dot plots showing selected proteins demonstrating shared up- or down-regulated patterns across myositis autoantibody subgroups compared with healthy controls. Proteins shown include CXCL10, LTA4H, MCP-3, CRH, CXCL11, CES2, 4E-BP1, and YES1. These proteins were selected to illustrate a shared inflammatory signature, including chemokine and leukotriene-related proteins, along with selected downregulated structural or regulatory proteins. (B) Dot plots showing selected proteins with prominent autoantibody-associated expression patterns across healthy controls and myositis autoantibody subgroups. Proteins shown include CXCL9, ATP6AP2, PSMA1, TNNI3, CASP8, TIGAR, IL-6, and FKBP1B. These proteins were selected to illustrate subgroup-enriched patterns, including Jo-1-associated chemokine/interferon activation, MDA5-associated proteostasis and antigen-processing signals, PL-7-associated apoptotic/innate immune activation, and PL-12-associated cytokine signaling. EJ and TIF1γ groups are not displayed separately because of small sample sizes. Each point represents an individual participant, and horizontal lines indicate group medians. Protein abundance is shown as Olink normalized protein expression. Abbreviations: HC, healthy control; IIM, idiopathic inflammatory myopathy; NPX, normalized protein expression.

Patients with lower-ILD-risk autoantibodies showed partial overlap with the shared inflammatory signature and interferon response, including elevations in CES2, LTA4H, MCP-3/CCL7, CASP8, CXCL10, CXCL11, ATP6AP2, PSMA1, MCP-2/CCL8, and ST3GAL1 when compared to controls (Supplementary Figure S1). In the direct comparison, higher-ILD-risk patients had greater abundance of proteins linked to injury and stress, including adenosine deaminase (ADA), MAP4K5, NCF2, FES, BANK1, 4E-BP1/EIF4EBP1, TIGAR, RASSF2, betacellulin (BTC), and syntaxin-8 (STX8) (Figure 1B). These findings suggest that interferon-response and other cytokine proteomic signals are shared across IIM autoantibody groups, whereas higher-ILD-risk autoantibodies show greater activation of apoptotic, oxidative-stress, and remodeling-related pathways. Analyses stratified by ILD status are shown in Supplementary Figures S2 and S3.

Pathway analyses supported these group-level findings (Supplementary Figure S4). Hallmark enrichment analysis identified IL-6/JAK/STAT3 signaling, inflammatory response, interferon-gamma response, interferon-alpha response, and allograft rejection among the top enriched pathways in both higher-ILD-risk and lower-ILD-risk autoantibody groups compared with controls. Thus, both type I and type II interferon-response programs were represented. Ingenuity Pathway Analysis similarly identified pathogen-induced cytokine-storm signaling and macrophage classical activation among the top pathways in both groups (Supplementary Figures S5 and S6), with greater enrichment of IL-6-type cytokine/JAK signaling in the higher- ILD-risk group.

Network-level enrichment analysis further supported a coordinated inflammatory and injury-remodeling architecture across autoantibody-defined comparisons. Highly overlapping pathway modules centered on chemokine-mediated recruitment; IL-6/JAK/STAT3 and TNF/NF-κB signaling; and downstream tissue-injury response programs involving programmed cell death, TGF-β receptor signaling, focal adhesion, and PI3K/AKT-related growth-factor signaling. Several autoantibody-defined groups showed partial overlap within these modules without reaching formal pathway-level significance. Together, these findings support interferon-responsive CXCR3 chemokine signaling, IL-6/JAK/STAT3 inflammation, and apoptosis, with inflammation-injury-remodeling pathways differentially amplified across serologic subgroups.

## Autoantibody-associated proteomic signatures among higher-ILD-risk subgroups

Proteomic profiles were also examined within each higher-ILD-risk autoantibody group. Compared with healthy controls, anti-Jo-1 patients demonstrated the strongest interferon/CXCR3 chemokine profile. CXCL9 was the most upregulated protein (Figure 2B) and was accompanied by increased CASP8, LTA4H, CXCL10, CXCL11, FES, ATP6AP2, MCP-3, CES2, and CAPG. This CXCL9-dominant pattern supports a prominent IFN-γ/type II interferon-responsive component in anti-Jo-1 disease. Within anti-Jo-1-positive patients, quantitative anti-Jo-1 levels correlated with CXCL9, CXCL10, CXCL11, IL-15RA, MCP-4, PDCD1, and MCP-2 (Figure 3), supporting interferon/CXCR3 chemokine signaling, T-cell activation, and monocyte recruitment.

**Figure 3.**
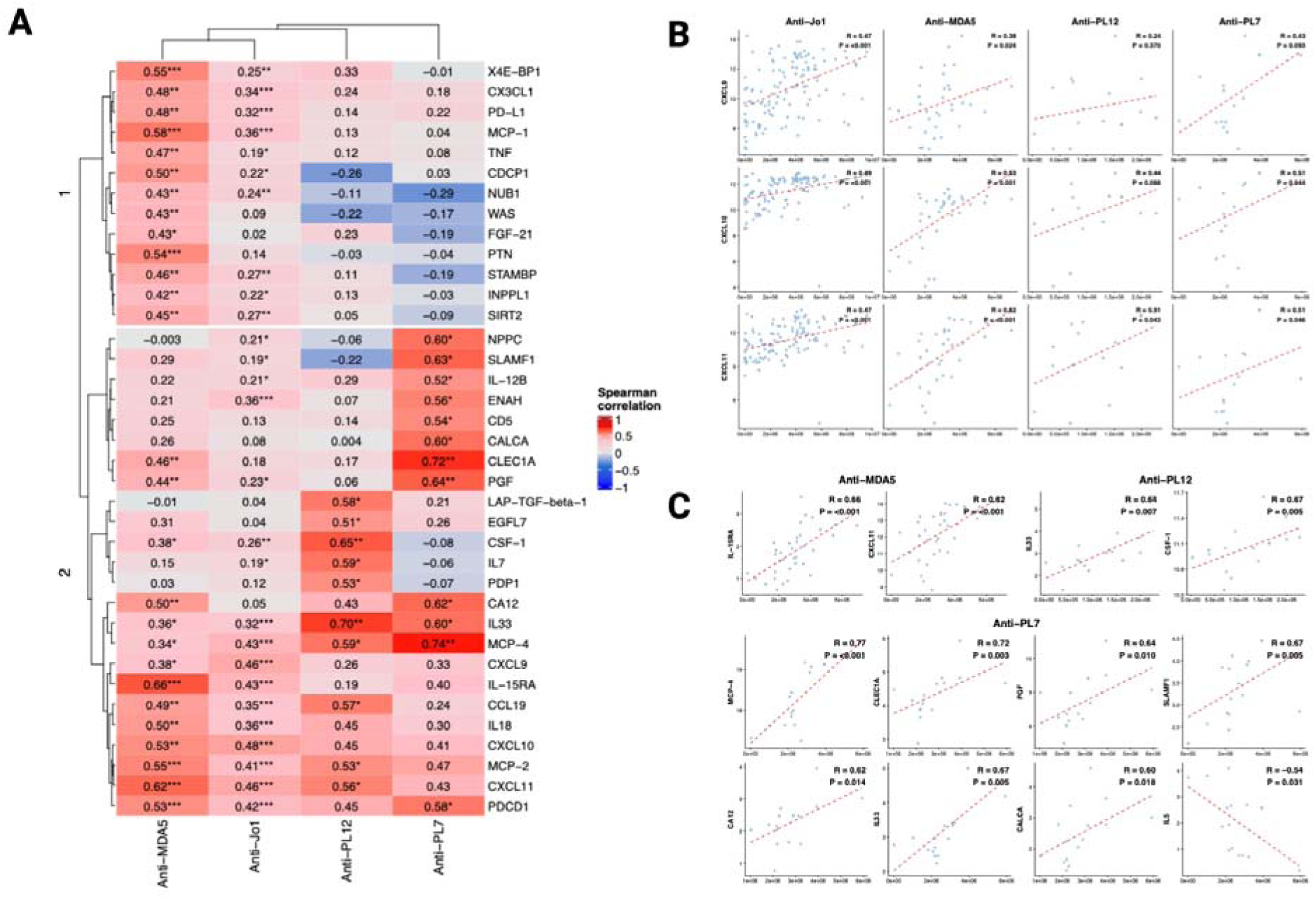
Quantitative autoantibody levels correlate with shared and autoantibody- associated serum proteins. (A) Heatmap showing Spearman correlations between quantitative autoantibody levels and selected serum proteins across higher-ILD-risk autoantibody subgroups. Red indicates positive correlation and blue indicates negative correlation. Values within cells represent Spearman correlation coefficients, and asterisks indicate nominal statistical significance. Proteins were clustered according to similarity in correlation patterns, highlighting shared and autoantibody- associated relationships between autoantibody burden and circulating inflammatory, immune- regulatory, vascular, and tissue-stress proteins. (B) Scatter plots showing correlations between quantitative autoantibody levels and interferon- related chemokines CXCL9, CXCL10, and CXCL11 across anti-Jo-1, anti-MDA5, anti-PL-12, and anti-PL-7 subgroups. (C) Scatter plots showing selected autoantibody-protein correlations in anti-MDA5, anti-PL-12, and anti-PL-7 subgroups. Proteins were selected to illustrate representative associations involving immune-regulatory, cytokine, endothelial, and tissue-stress proteins. Each point represents an individual participant. Dashed lines represent fitted trend lines. Spearman correlation coefficients and nominal P values are shown within scatter plot panels. Protein abundance is shown as Olink normalized protein expression. Abbreviations: ILD, interstitial lung disease; NPX, normalized protein expression. *P<0.05; **P<0.01; ***P<0.001; nominal P values.

Compared with healthy controls, anti-MDA5 patients demonstrated an immune-stress profile characterized by interferon-related inflammation, proteostasis and antigen-processing signals, and vascular/cellular stress markers. Among the autoantibody subgroups, anti-MDA5 showed the strongest CXCL10 increase, consistent with a prominent interferon-responsive component, although CXCL10 is not specific to type I interferon. ATP6AP2 and PSMA1 showed the greatest relative upregulation in this group, whereas TNNI3 was the most strongly downregulated protein across all autoantibody subtypes (Figure 2B). Within anti-MDA5-positive patients, quantitative anti-MDA5 levels correlated with IL-15RA, CXCL11, CXCL10, PTN, CDCP1, TNF, PGF, EIF4EBP1, NUB1, WAS, and INPPL1 (Figure 3).

Compared with healthy controls, anti-PL-12 patients showed comparatively greater involvement of cytokine, macrophage/growth-factor, epithelial, and profibrotic markers. Upregulated proteins included IL-6, CXCL11, LTA4H, FES, CES2, MCP-3, and LAP-TGF-β1. IL-6 and FKBP1B showed the highest relative upregulation in this subgroup (Figure 2B). Within anti-PL-12-positive patients, quantitative anti-PL-12 levels correlated with IL-7, LAP-TGF- β 1, CSF1, PDP1, and EGFL7 (Figure 3), suggesting a biological profile distinct from the chemokine-dominant pattern observed in anti-Jo-1 patients.

Anti-PL-7 patients showed the most prominent apoptosis and innate immune activation profile among antisynthetase subtypes, with a distinct metabolic/redox-stress component compared with healthy controls. CASP8 was the most upregulated protein (Figure 2B), with additional elevation of FES, S100A12/EN-RAGE, TIGAR (Figure 2B), ATP6AP2, ST3GAL1, and CXCL10. Within the anti-PL-7-positive patients, quantitative anti-PL-7 levels correlated with MCP-4, CLEC1A, PGF, CA12, NPPC, ENAH, SLAMF1, CALCA, and IL-12B (Figure 3), supporting monocyte recruitment, endothelial signaling, and vascular or neuroimmune stress pathways.

Correlations between quantitative autoantibody levels and protein abundance are summarized in Figure 3A. Higher anti-Jo-1 levels were associated predominantly with CXCR3 chemokines and immune-regulatory or monocyte-recruiting proteins; anti-MDA5 with interferon-responsive and vascular/cellular-stress proteins; anti-PL-12 with IL-7, CSF1, LAP-TGF-β1, PDP1, and EGFL7; and anti-PL-7 with monocyte-recruitment, endothelial, and stress-related proteins. Selected chemokine and non-chemokine associations are shown in Figures 3B and 3C, respectively. No significant associations were observed between quantitative autoantibody levels and baseline pulmonary function (Supplementary Figure S7).

## Proteomic clustering identifies distinct injury-stress and immune-activation endotypes

Unsupervised proteomic clustering of 204 patients identified four distinct endotypes not fully captured by autoantibody class. Cluster-specific proteomic patterns are shown in Figure 4A, UMAP visualization in Figure 4B, and survival by cluster in Figure 4C. Selected characteristics are provided in Table 2, with full characteristics in Supplementary Table S3. A focused heatmap of 38 representative proteins further illustrates lower disease-associated protein abundance in Cluster 1, intermediate immune activation in Cluster 2, injury-stress signaling in Cluster 3, and broad chemokine/checkpoint activation in Cluster 4 (Supplementary Figure S8).

**Figure 4.**
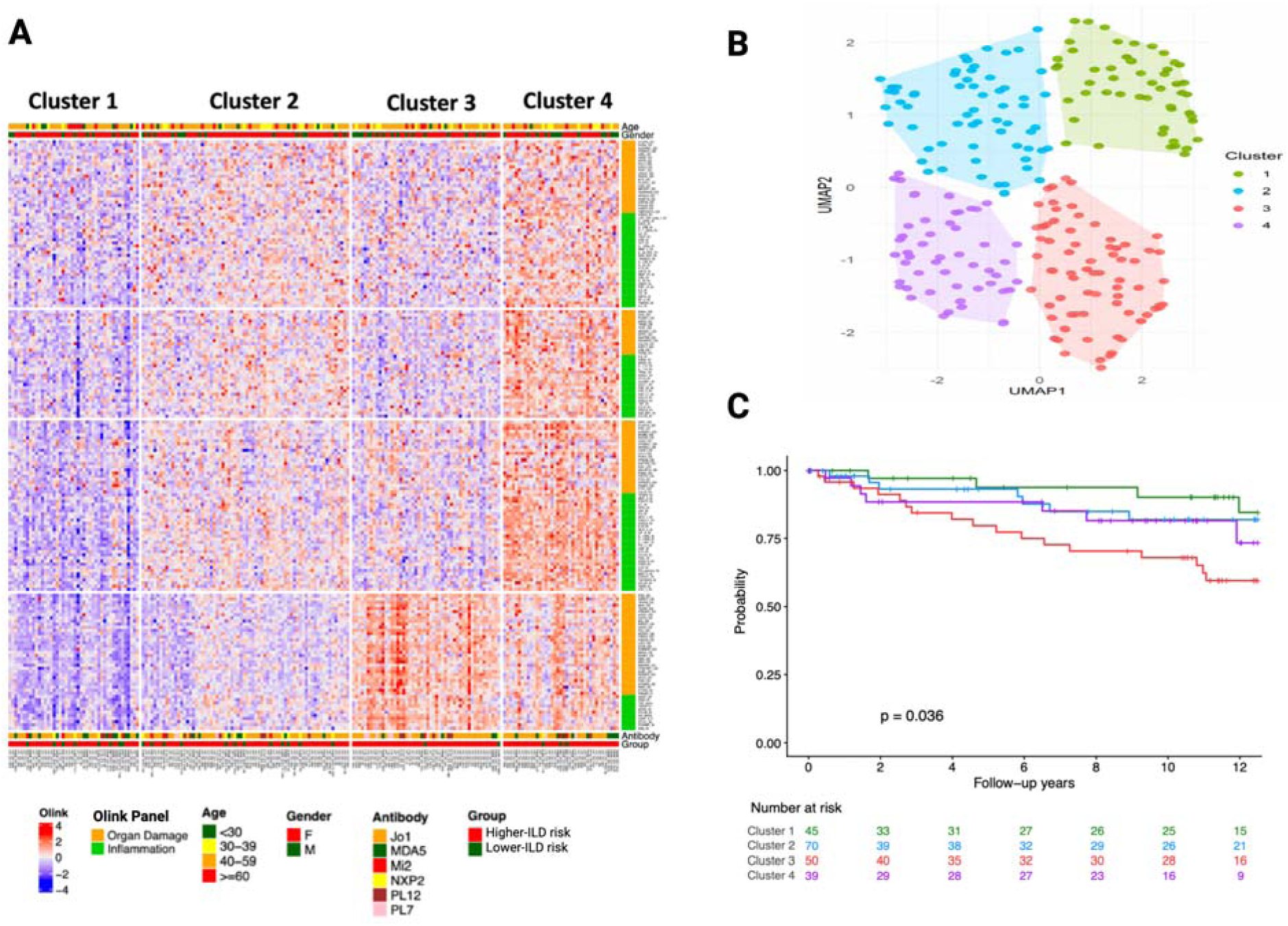
Proteomic clustering identifies distinct injury-stress and immune-activation endotypes. (A) Heatmap from unsupervised clustering of targeted serum proteomic profiles. Columns represent individual participants and rows represent proteins. Protein abundance is shown as normalized Olink NPX values scaled by protein, with higher relative expression shown in red and lower relative expression shown in blue. Row annotation indicates Olink panel type, including Inflammation and Organ Damage proteins. Column annotations indicate age group, sex, autoantibody type, and ILD-risk group. Four proteomic clusters were identified among 204 patients. Cluster 3 showed higher expression of proteins related to inflammatory stress, apoptosis, injury, and remodeling pathways, consistent with an injury-stress proteomic profile. Cluster 4 showed relatively higher expression of chemokine, checkpoint, and costimulatory proteins, consistent with a chemokine/checkpoint-high immune-activation profile. (B) UMAP visualization of patient-level proteomic profiles after scaling. Each point represents an individual patient and is labeled according to the four clusters identified by k-means clustering of the scaled, batch-corrected protein-expression data. Shaded polygons indicate the spatial distribution of patients within each cluster. Cluster 1 is shown in green, Cluster 2 in sky blue, Cluster 3 in red, and Cluster 4 in purple. (C) Kaplan-Meier curves showing all-cause survival according to proteomic cluster assignment. Cluster 3 showed poorer survival compared with other clusters. The number at risk is shown below the plot. P value was calculated using the log-rank test. Abbreviations: ILD, interstitial lung disease; NPX, normalized protein expression.

**Table 2.**
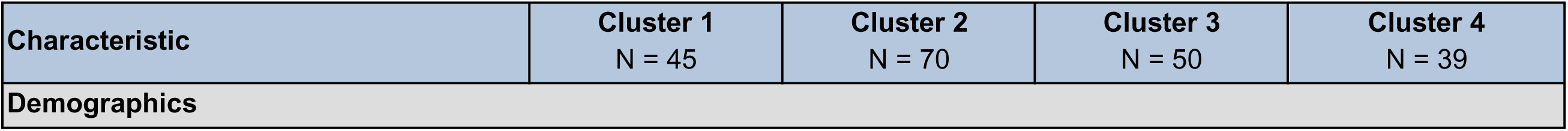
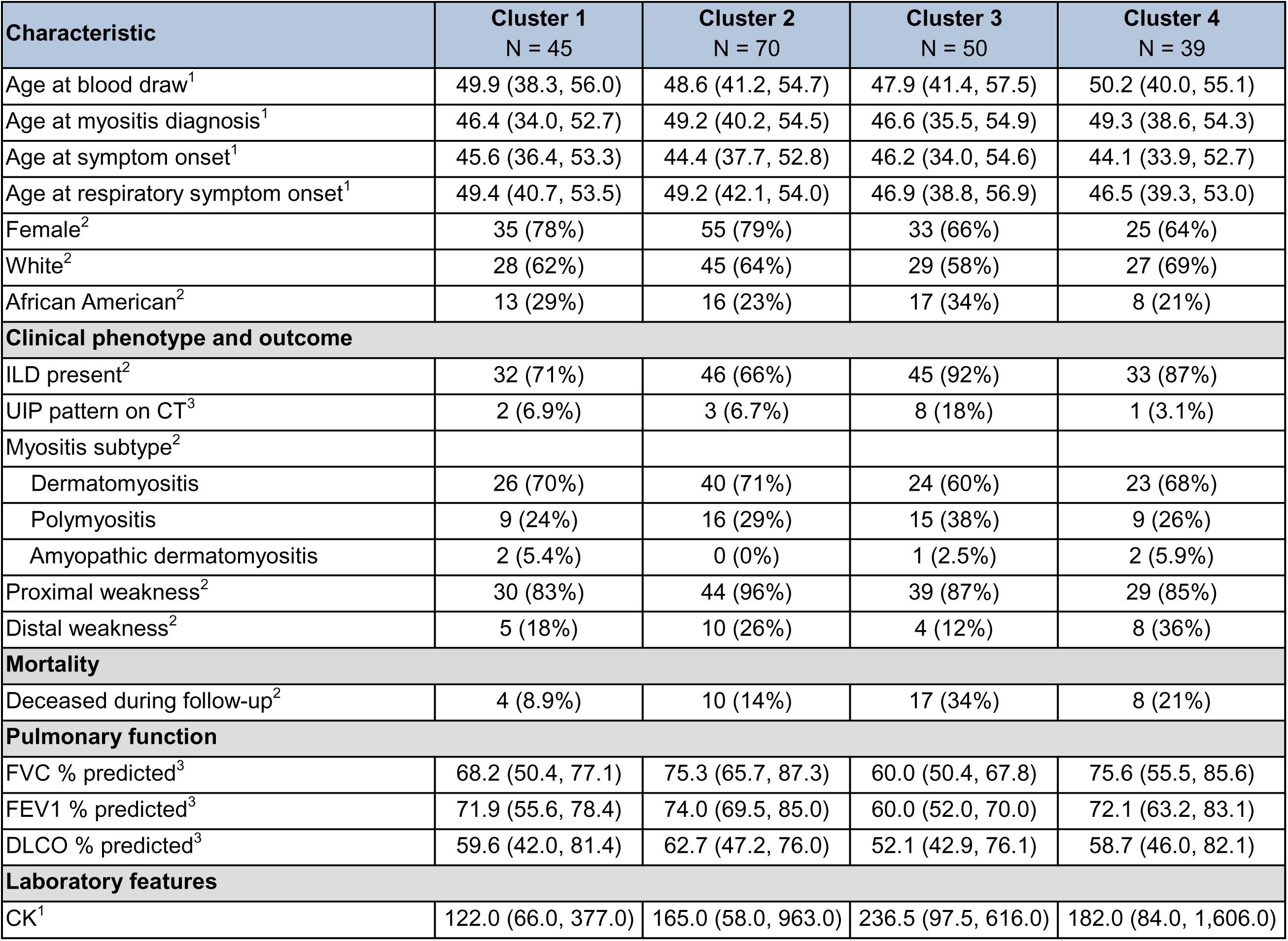
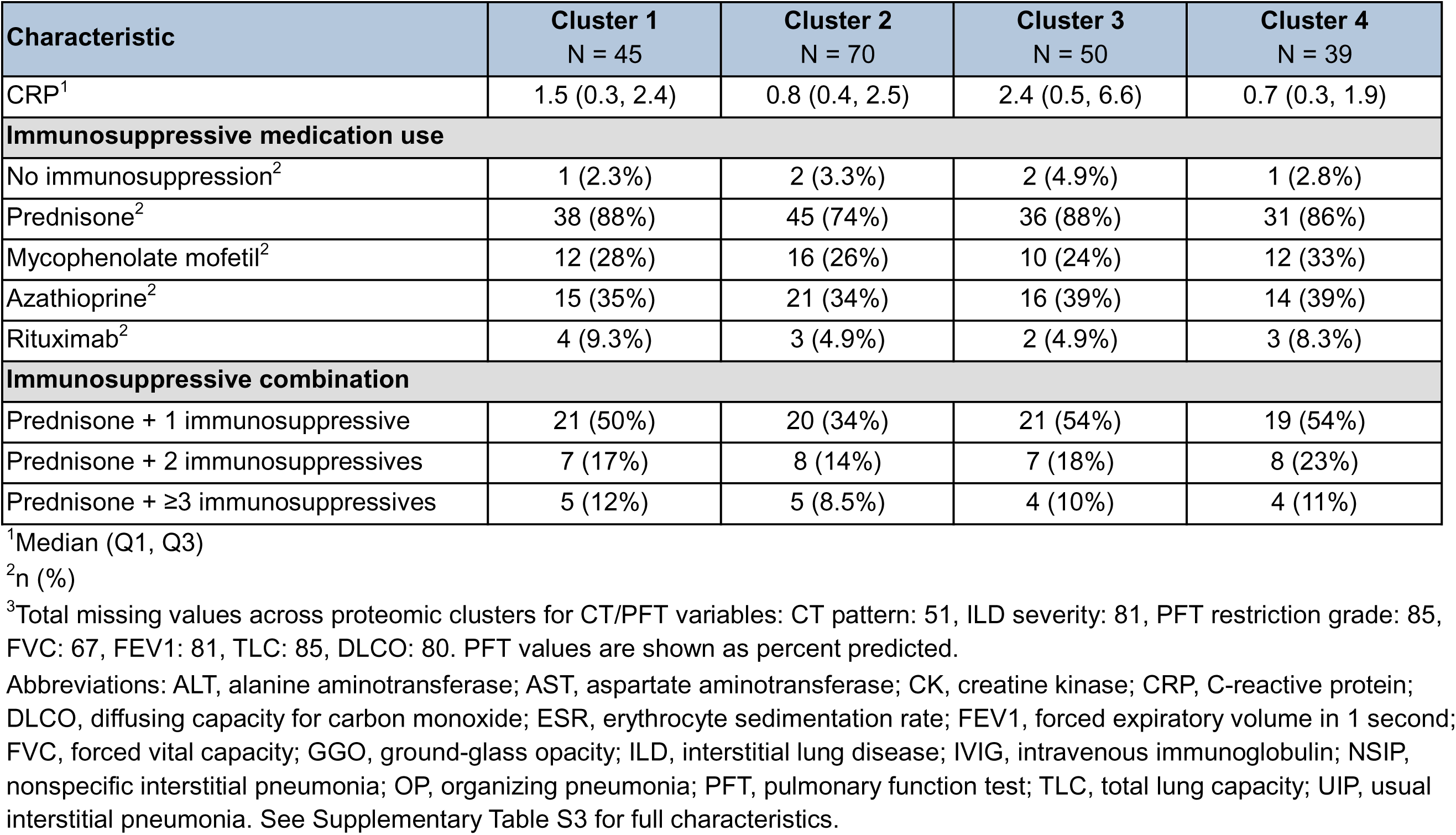
Baseline clinical characteristics stratified by proteomic cluster.

Autoantibody classes were distributed across clusters, although partial enrichment was observed. Most PL-7-positive patients clustered in Cluster 3, Mi-2- and NXP2-positive patients were largely restricted to Clusters 1 and 2, and Jo-1- and MDA5-positive patients were distributed across multiple clusters. Quantitative anti-Jo-1 levels were higher in Clusters 3–4 than in Clusters 1–2 (Supplementary Figure S9). Although Cluster 1 had the lowest relative disease-associated protein abundance, all four clusters remained substantially different from healthy controls.

Cluster 3 represented a distinct injury-stress endotype. CASP8, FES, TIGAR, NUB1, BID, NCF2, PXN, OSM, CAPG, and EN-RAGE were increased relative to controls and the other clusters, implicating apoptosis, oxidative/metabolic stress, cytoskeletal remodeling, innate immune activation, and inflammatory-stress signaling. Cluster 3 was associated with worse lung function and poorer survival (Table 2; Figure 4C).

Cluster 4 showed a chemokine/checkpoint-high immune-activation profile, with prominent increases in CXCL9, CXCL10, CXCL11, MCP-3, MCP-2, IL-6, PD-L1, PDCD1, TNFRSF9, IL-15RA, CD40, and PGF. Despite this prominent immune activation, Cluster 4 had relatively preserved lung function compared with Cluster 3. These proteomic patterns remained detectable despite prevalent baseline immunosuppression. Detailed differential-expression and pairwise cluster comparisons are provided in the Supplementary Results.

## Proteomic mortality scores identify patients at increased risk of all-cause mortality

Survival analyses included 206 participants with 38 deaths for the 5-protein score and 203 participants with 38 deaths for the 10-protein weighted score. FVC-adjusted analyses included 149 participants with 33 deaths and 147 participants with 33 deaths, respectively. Cox and random survival forest analyses were used to prioritize proteins for candidate composite scores (Figure 5; Supplementary Table S4 and Supplementary Figure S10). The unweighted 5-protein score included IL-7, OSM, VEGFA, IL-6, and TGF-α, whereas the 10-protein weighted score additionally included CRH, IL-17C, TNF-β, CD244, and NOS3.

**Figure 5.**
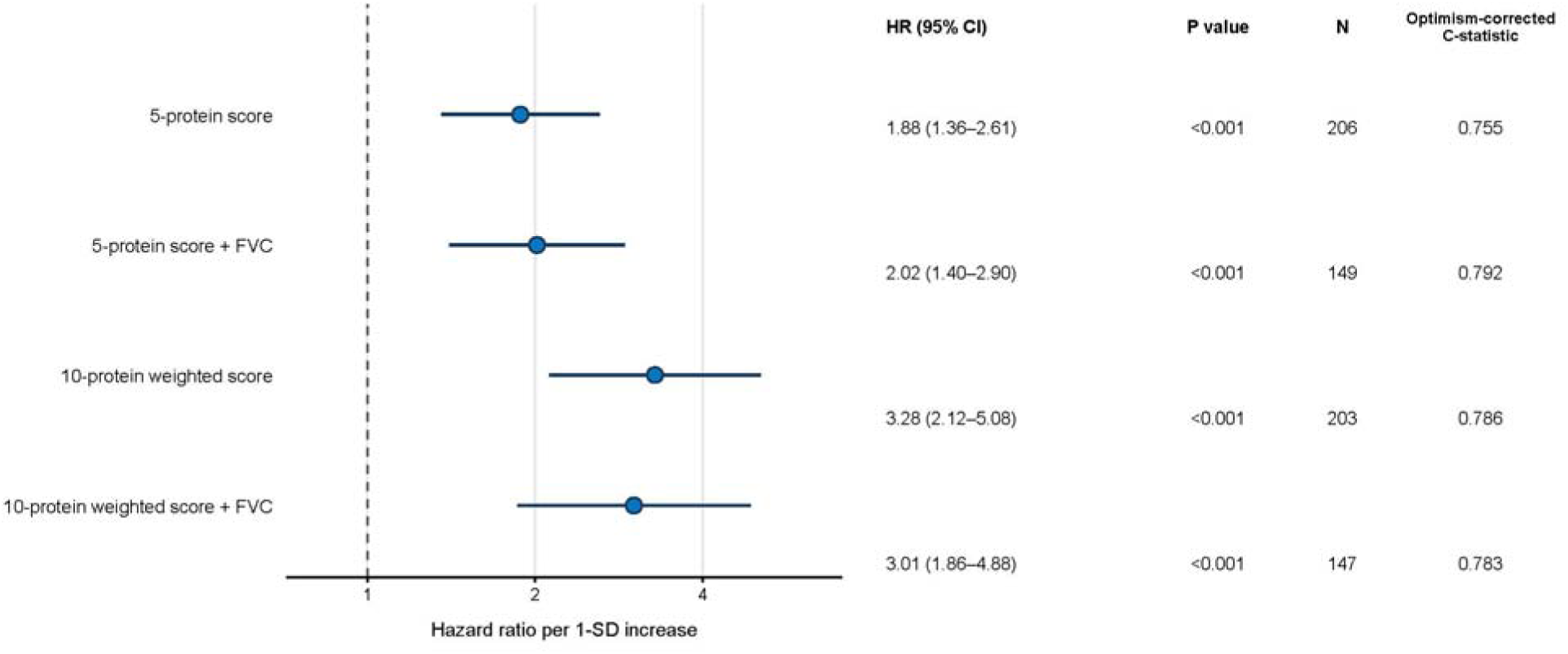
Proteomic survival scores are associated with all-cause mortality after adjustment for ILD status, with and without additional adjustment for baseline FVC. Forest plot showing the adjusted associations of the 5-protein score and 10-protein weighted score with all-cause mortality. The 5-protein score was calculated as the unweighted sum of IL- 7, OSM, VEGFA, IL-6, and TGF-α. The 10-protein weighted score was calculated as a Cox coefficient-weighted linear combination of these five proteins plus CRH, IL-17C, TNF-β, CD244, and NOS3. All models were adjusted for age, sex, and ILD status. Models additionally adjusted for baseline FVC were restricted to participants with available FVC measurements. Hazard ratios and 95% confidence intervals were obtained from the original Cox proportional hazards models and are reported per 1-SD increase, using reference SDs estimated from the corresponding full analysis samples and applied to both the primary and FVC-adjusted models (3.63 for the 5-protein score and 1.31 for the 10-protein weighted score). Points represent hazard ratios, and horizontal lines represent 95% confidence intervals; estimates are displayed on a logarithmic scale. C-statistics describe the discrimination of the complete adjusted models and were internally validated using 1,000 bootstrap resamples with correction for optimism. For the 5-protein score models, the score was held fixed during bootstrapping. For the 10-protein weighted score models, the protein weights and resulting score were re-estimated within each bootstrap resample because the weights were derived from the survival outcome. All 1,000 bootstrap iterations were successfully completed for each model. Abbreviations: CI, confidence interval; FVC, forced vital capacity; HR, hazard ratio; ILD, interstitial lung disease.

Patients with a baseline 5-protein score 3.63 units higher, equivalent to 1 SD, had an 88% higher hazard of all-cause mortality after adjustment for age, sex, and ILD status (HR, 1.88; 95% CI, 1.36–2.61; p<0.001). The association remained significant after additional adjustment for baseline FVC % predicted (HR, 2.02; 95% CI, 1.40–2.90; p<0.001).

Patients with a baseline 10-protein weighted score 1.31 units higher, equivalent to 1 SD, had a 3.28-fold hazard of all-cause mortality after adjustment for age, sex, and ILD status (HR, 3.28; 95% CI, 2.12–5.08; p<0.001). The association remained significant after additional adjustment for baseline FVC % predicted (HR, 3.01; 95% CI, 1.86–4.88; p<0.001). Optimism-corrected C-statistics were 0.755 and 0.792 for the 5-protein models without and with FVC, respectively, and 0.786 and 0.783 for the corresponding 10-protein models.

Exploratory quartile analyses suggested potentially nonlinear associations for several score components. OSM showed the strongest survival separation and was also enriched in the injury-stress endotype, linking the mortality and clustering analyses (Supplementary Figure S11 and Supplementary Results).

## Exploratory longitudinal lung-function analysis

Matched proteomic and longitudinal pulmonary-function data were available for 62, 56, 47, and 45 patients at 6, 12, 18, and 24 months, respectively. At 12 months, 6 patients met the primary definition of decline, 18 improved, and 32 remained stable. Given the limited number of decline events, these analyses were considered exploratory. Across complementary random forest and LASSO analyses, SERPINA9 was the most reproducible candidate marker of decline, whereas improvement-associated signals were less stable. No protein was consistently identified across all follow-up time points and outcome definitions (Supplementary Figures S12–S15 and Supplementary Results).

## Discussion

In this study, we integrated quantitative myositis-specific autoantibodies and targeted serum proteomics profiling to characterize biological heterogeneity in an IIM-ILD-enriched cohort. Three findings are central. First, IIM patients, particularly those with higher-ILD-risk autoantibodies, shared a broad interferon-responsive inflammatory signature marked by CXCR3 chemokine signaling, IL-6/JAK/STAT3 signaling, monocyte-recruiting factors, and apoptotic mediators. Second, quantitative autoantibody levels correlated with distinct proteomic patterns, suggesting that quantitative autoantibody burden may provide biological information beyond categorical serologic status. Third, unsupervised proteomic clustering identified endotypes that were not fully explained by autoantibody class, including an injury-stress endotype associated with impaired lung function and poorer survival.

Prior ILD proteomic studies have distinguished connective tissue disease-ILD (CTD-ILD) from IPF and identified validated survival proteins and molecular endotypes beyond conventional diagnosis and baseline physiology (22, 23, 25, 29–32). Our study extends this framework within IIM by resolving distinct proteomic states among patients who may share the same myositis autoantibody. It also addresses a gap in prior myositis molecular studies, which often had limited representation of antisynthetase autoantibodies strongly associated with ILD. By including 199 patients with higher-ILD-risk autoantibodies spanning anti-Jo-1, anti-MDA5, anti-PL-7, anti-PL-12, and anti-EJ, we integrated quantitative serology with proteomic endotypes and outcomes in subgroups directly relevant to pulmonary risk.

The shared inflammatory signature observed across higher-ILD-risk autoantibody groups is consistent with prior proteomic and transcriptomic studies showing that interferon-related chemokines, innate immune activation, intracellular signaling, and tissue-remodeling programs are recurrent features of adult and juvenile myositis (27, 35–37, 44, 45). PI3K/AKT-related signaling also overlapped with pathways identified in prior integrative transcriptomic and proteomic studies of dermatomyositis, supporting shared intracellular signaling across myositis phenotypes(27, 44). CXCL9 is more strongly associated with IFN-γ/type II interferon signaling, whereas CXCL10 and CXCL11 can reflect both type I and type II interferons. The CXCL9- dominant anti-Jo-1 profile is therefore consistent with prominent type II interferon signaling in antisynthetase syndrome and anti-Jo-1 myositis (46, 47), while CXCL10 and CXCL11 have also been linked to inflammatory burden and an ILD-associated dermatomyositis endotype (48).

Longitudinal juvenile dermatomyositis studies have shown that some proteomic abnormalities persist despite treatment, while others re-emerged during treatment tapering(36, 37). Consistent with this distinction, our findings suggest that inflammatory activation alone does not define the highest-risk biology. The injury-stress endotype points to a broader program in which immune activation converges with apoptosis, oxidative or metabolic stress, vascular remodeling, and tissue-repair pathways. This framework may help explain why patients with similar serologic risk can differ substantially in lung function, trajectory, and survival.

Autoantibody-associated proteomic patterns provided additional evidence of biological heterogeneity within IIM-ILD. Although interferon-response and other cytokine signals were shared, the dominant signals differed by autoantibody specificity. Anti-Jo-1 disease showed a prominent interferon/CXCR3 chemokine profile with evidence of T-cell activation and monocyte recruitment, whereas anti-MDA5 disease showed stronger signals related to immune stress, proteostasis, antigen processing, and vascular or cellular stress. Prior plasma proteomic studies identified KRT19 as a candidate biomarker in anti-MDA5-positive dermatomyositis and independently associated SPP1/osteopontin with rapidly progressive ILD(33, 34). Myositis-specific-autoantibody-stratified serum mass spectrometry linked anti-MDA5-positive juvenile dermatomyositis to type I interferon and immunoproteasome pathways and identified coagulation-related proteins and SFTPD among MDA5- or ILD-associated signals(38). Longitudinal Olink profiling of juvenile dermatomyositis, anti-MDA5-positive patients showed elevated circulating SFTPA1 and SFTPA2(36). Together, these data support a combined epithelial-injury, vascular/coagulation, macrophage, interferon, and proteostasis model of anti- MDA5-associated lung disease. Our ATP6AP2/PSMA1-predominant pattern and correlations between quantitative anti-MDA5 levels and vascular or cellular-stress proteins extend this model to continuous autoantibody burden. Anti-PL-12 disease showed greater involvement of IL-6, macrophage/growth-factor, epithelial, and profibrotic signals, while anti-PL-7 disease showed prominent apoptosis and innate immune activation with a metabolic/redox-stress component. These subgroup patterns should not be interpreted as discrete mechanisms, given overlap across serologic groups; rather, they suggest that shared immune-injury signals may be differentially amplified across autoantibody-defined subsets.

The clustering analysis further supports clinically relevant biology beyond traditional autoantibody categories. Autoantibody specificity contributed to cluster composition but did not determine it. Cluster 3 combined apoptosis, oxidative/metabolic stress, cytoskeletal remodeling, innate immune activation, and inflammatory-stress signaling and was associated with worse lung function and survival. Subtype-resolved muscle multiomics similarly separated interferon biology from structural, extracellular-matrix, and metabolic/endothelial injury programs across IIM subtypes (45). Although tissue and circulating protein directions need not match, these data support redox/metabolic stress, proteostasis, and remodeling as plausible components of the circulating high-risk endotype. In contrast, Cluster 4 showed marked chemokine/checkpoint activation with relatively preserved lung function, indicating that inflammatory abundance alone did not identify the most physiologically severe state. Comparable endotyping studies in IPF and non-IPF ILD identified molecular classes with divergent outcomes (30, 32), although our injury- stress endotype was biologically distinct from their more epithelial-, macrophage-, and remodeling-dominant profiles. Higher anti-Jo-1 levels in Clusters 3-4 may indicate greater proteomic activation but should not be interpreted as directly indicating worse prognosis.

Longitudinal lung-function analyses were exploratory because data were available in a smaller subset and relatively few patients met decline criteria. SERPINA9 emerged as a candidate marker of decline, whereas improvement-associated signals were less stable; no protein was consistent across all time points and outcome definitions. In contrast, the survival analyses were more robust. Both mortality scores remained associated with survival after adjustment for ILD status and baseline FVC, suggesting that the proteomic signal was not fully explained by baseline pulmonary impairment. Their inflammatory, vascular, endocrine/stress, and injury-related components differed from epithelial and dysregulated-repair signatures reported in broader ILD cohorts(25, 31, 33), potentially reflecting systemic IIM biology, targeted panel composition, or both. Exploratory quartile analyses suggested potentially nonlinear relationships, particularly for OSM, whereas inverse associations involving CRH and TNF-β may reflect treatment exposure, disease severity, immunosuppression, or residual confounding rather than direct biological effects. Because the scores were developed and internally validated within this cohort, they remain hypothesis generating and require external validation.

These findings support interpreting quantitative autoantibody measurements together with downstream proteomic patterns. Quantitative autoantibody levels were associated with interferon, immune-regulatory, profibrotic, and metabolic/vascular proteins. The enrichment of interferon-related chemokines and IL-6/JAK/STAT3 signaling is notable because these pathways are biologically plausible and pharmacologically targetable. Distinct inflammatory and injury-stress signals also remained detectable despite prevalent immunosuppression, potentially reflecting ongoing disease activity, accumulated injury, treatment-resistant biology, or confounding by indication. A non-IPF ILD study found that molecular endotype modified the observed association between immunosuppressant exposure and survival, although treatment was not randomized (32). Our study was not designed to evaluate treatment response, but prospective serial sampling could determine whether proteomic endotypes and quantitative autoantibody levels track disease activity or therapeutic response (36, 37).

Several limitations should be considered. The targeted serum panels did not comprehensively capture pulmonary epithelial and fibroproliferative proteins, and the absence of IPF or non-IIM CTD-ILD comparators prevented assessment of disease specificity. The tissue sources of circulating proteins remain uncertain. Treatment exposure was heterogeneous, most measurements were cross-sectional, and longitudinal pulmonary-function data were available in a subset. Finally, the clustering, random forest analyses, and mortality scores require external validation, and differences in specimen type, platform, analyte coverage, and preprocessing may limit comparisons across studies.

## Conclusion

In summary, integrated quantitative autoantibodies and targeted serum proteomic profiling revealed shared inflammatory biology, autoantibody-associated signatures, and proteomic endotypes in IIM-ILD. An injury-stress endotype was associated with lung impairment and poor survival, supporting further evaluation of risk stratification beyond categorical serology and biologically informed longitudinal and therapeutic studies.

## Author Contributions

**Conceptualization:** J.A. Huapaya, S.K. Danoff, A.F. Suffredini

**Methodology:** J.A. Huapaya, P.D. Burbelo, J. Ward, N.R. Redekar, X. Tian, S. Gao

**Formal analysis:** J.A. Huapaya, X. Tian, S. Gao, J. Ward, N.R. Redekar

**Investigation:** J.A. Huapaya, P.D. Burbelo, E.W. Robbins, S. Turan, S. Gairhe, G. Pastor, N. Gupta, P.N. Farhadi, K. Sarkar

**Resources:** J.L. Li, M. Casal-Dominguez, I. Pinal-Fernandez, L. Christopher-Stine, A. Schiffenbauer, L.G. Rider, A. Mammen, S.K. Danoff, A.F. Suffredini

**Data curation:** J.A. Huapaya, E.W. Robbins, N. Gupta, S. Turan, S. Gairhe, G. Pastor, J. Ward, N.R. Redekar

**Software:** J. Ward, N.R. Redekar

**Validation:** J.A. Huapaya, P.D. Burbelo, X. Tian, S. Gao, J. Ward, N.R. Redekar

**Visualization:** J.A. Huapaya, X. Tian, S. Gao, J. Ward, N.R. Redekar

**Writing –** original draft: J.A. Huapaya

**Writing –** review and editing: All authors

**Supervision:** J.A. Huapaya, P.D. Burbelo, J.L. Li, L.G. Rider, A. Mammen, S.K. Danoff, A.F. Suffredini

**Project administration:** J.A. Huapaya

All authors reviewed and approved the final manuscript and agree to be accountable for their respective contributions.

## Impact Statement

By integrating quantitative myositis-specific autoantibodies with serum proteomics, this study identifies molecular endotypes that capture clinically relevant heterogeneity beyond categorical serology in myositis-associated interstitial lung disease. The injury-stress endotype and mortality-associated protein score establish a biologically grounded framework for risk stratification and future biomarker development.

## Data Availability

All data produced in the present study are available upon reasonable request to the authors

## Acknowledgements

We thank all the members of the Johns Hopkins Myositis Center and the patients who generously contributed to this study. We also acknowledge the Johns Hopkins Myositis Center Biobank team for their essential contributions in processing, storing, and managing patient samples. Finally, we thank Kelly Byrne for her assistance with the submission process. Figures 1-5 were assembled using BioRender.com.

## AI tools

OpenAI ChatGPT was used solely to assist with language editing of the manuscript. It was not used to generate or analyze study data, perform statistical analyses, or create or modify figures. No identifiable patient information or other sensitive nonpublic data were entered. All AI-assisted text was critically reviewed and revised by the authors, who take full responsibility for the final content.

## Data availability statement

Data are available upon request.

