## Supplementary Material for "Proteomic Signatures and an Injury-Stress Endotype in Myositis-Associated Interstitial Lung Disease"

**Supplementary Material Contents**

**Supplementary Methods: pages 2-4**

-Study cohort and clinical definitions: page 2

-Targeted serum proteomics: page 2

-Autoantibody quantification: page 2

-Differential-expression and correlation analyses: pages 2-3

-Pathway enrichment: page 3

-Proteomic clustering: page 3

-Lung-function trajectory analyses: page 3

-Mortality analyses: pages 3-4

**Supplementary Results: pages 5-6**

-Detailed proteomic cluster comparisons: page 5

-Exploratory survival analyses of individual score components: page 5

-Exploratory longitudinal lung-function analyses: pages 5-6

**Supplementary Figures: pages 7-27**

-Supplementary Figure S1: page 7

-Supplementary Figure S2: page 8

-Supplementary Figure S3: page 9

-Supplementary Figure S4: page 10

-Supplementary Figure S5: pages 11-12

-Supplementary Figure S6: pages 13-14

-Supplementary Figure S7: page 15

-Supplementary Figure S8: page 16

-Supplementary Figure S9: page 17

-Supplementary Figure S10: page 18

-Supplementary Figure S11: page 19

-Supplementary Figure S12: pages 20-21

-Supplementary Figure S13: pages 22-23

-Supplementary Figure S14: pages 24-25

-Supplementary Figure S15: pages 26-27

**Supplementary Tables: pages 28-44**

-Supplementary Table S1. Protein nomenclature: pages 28-35

-Supplementary Table S2. Full baseline clinical characteristics stratified by autoantibody group: pages 36-39

-Supplementary Table S3. Full clinical characteristics and outcomes stratified by proteomic cluster: pages 40-42

-Supplementary Table S4. Individual protein Cox proportional hazards models for all-cause mortality: pages 43-44

-Supplementary References: page 45

**Supplementary Methods**

**Study cohort and clinical definitions**

We analyzed 264 serum samples from 261 individuals, including 226 adults with IIM and 35 age-, sex-, and race-matched healthy controls recruited from the Johns Hopkins Myositis Center, the National Institute of Environmental Health Sciences, and the National Institute of Arthritis and Musculoskeletal and Skin Diseases between 2010 and 2020. Three participants had duplicate samples; one sample was retained for participant-level analyses.

IIM diagnoses were confirmed by the patients’ treating specialists using clinical, serologic, and histopathologic findings. Patients were categorized into higher-ILD-risk autoantibody groups, comprising Jo-1, MDA5, PL-7, PL-12, and EJ, and lower-ILD-risk groups, comprising Mi-2, NXP2, and TIF1γ(1). ILD was defined by high-resolution CT abnormalities consistent with IIM-associated ILD and/or physiologic restriction with impaired gas transfer, adjudicated by pulmonary and radiology review.

The study was approved by the institutional review boards of the participating centers (IRB00455938, IRB002669, 94E0165, and 11E0099), and all participants provided written informed consent.

**Targeted serum proteomics**

Circulating proteins were measured using the proximity-extension assay platform (Olink, Waltham, MA), which reports normalized protein expression values on a log2 scale. The Inflammation and Organ Damage panels were used, comprising 184 analytes. Samples failing vendor-defined quality-control criteria were excluded.

Proteins are reported in the manuscript using the corresponding Olink assay labels. Their Uniprot, gene symbols, Olink ID, and common aliases are provided in Supplementary Table S1.

**Autoantibody quantification**

Myositis-specific autoantibodies were quantified using the luciferase immunoprecipitation systems assay, a quantitative method with a broad dynamic range. The assay procedures and validation have been described previously(2, 3) .

**Differential-expression and correlation analyses**

Between-group analyses compared higher- and lower-ILD-risk autoantibody groups and individual autoantibody-defined subgroups with healthy controls. Plate effects were adjusted using ComBat while preserving variation attributable to the biological covariates of interest.

Differentially expressed proteins were identified using linear models implemented in the limma package in R. Statistical significance was defined by a false discovery rate of 5% and an absolute mean fold change ≥1.25. Within each evaluable higher-ILD-risk subgroup, Spearman correlations were calculated between continuous quantitative autoantibody levels and normalized protein expression values. Correlation P values were nominal unless otherwise specified.

**Pathway enrichment**

Over-representation analysis of differentially expressed proteins was performed using the MSigDB Hallmark gene sets(4) and Ingenuity Pathway Analysis (Qiagen, Germantown, MD(5). Analyses evaluated biological processes, canonical pathways, predicted upstream regulators, and biological functions associated with differentially expressed proteins.

**Proteomic clustering**

Unsupervised clustering was performed using k-means applied to scaled, batch-corrected baseline protein-expression data from patients with complete proteomic data. The number of clusters was selected using the elbow method. Clinical characteristics, autoantibody composition, ILD status, pulmonary function, and survival were subsequently compared across the four clusters. Biological interpretation of each cluster was assigned manually based on cluster-associated differential expression patterns and the established functions of the enriched proteins. The selected representative proteins shown in Supplementary Figure S8 were used to illustrate the identified molecular programs and were not used to derive the clusters. UMAP was used to visualize patient-level similarities in two dimensions but was not used to determine cluster membership. Because anti-Jo-1-positive participants were represented across all four clusters, an exploratory analysis compared quantitative anti-Jo-1 levels between Clusters 1–2 and Clusters 3–4. Raw anti-Jo-1 levels were log10-transformed and compared using the Wilcoxon rank-sum test.

**Lung-function trajectory analyses**

Associations between baseline variables, including age and protein levels, and lung-function change were explored at 6, 12, 18, and 24 months using random forest models and least absolute shrinkage and selection operator–penalized logistic regression. Separate models evaluated decline versus all other categories and improvement versus all other categories. Random forests ranked variables according to predictive importance, whereas LASSO was used as a complementary variable-selection approach.

Lung-function decline was defined as an absolute decrease from baseline of ≥10 percentage points in FVC % predicted and/or ≥15 percentage points in DLCO % predicted. A less stringent sensitivity definition used absolute decreases of ≥5 percentage points in FVC % predicted and/or ≥10 percentage points in DLCO % predicted(6, 7). Improvement was defined using the corresponding thresholds in the opposite direction.

**Mortality analyses**

Associations between baseline protein levels and all-cause mortality were evaluated using Cox proportional hazards models adjusted for age, sex, and ILD status. Random survival forests were used as a complementary nonparametric approach to prioritize mortality-associated proteins.

Proteins supported by the Cox and random survival forest analyses were selected for candidate composite scores. The 5-protein score was calculated as the unweighted sum of IL-7, OSM, VEGFA, IL-6, and TGF-α. The 10-protein score additionally included CRH, IL-17C, TNF-β, CD244, and NOS3 and was weighted using Cox regression coefficients.

Composite-score analyses required complete data for all score components and included covariates. The full survival dataset contained 43 deaths. Complete-case requirements reduced the analytic samples to 206 participants with 38 deaths for the 5-protein score, 149 with 33 deaths for the FVC-adjusted 5-protein model, 203 with 38 deaths for the 10-protein weighted score, and 147 with 33 deaths for the corresponding FVC-adjusted model. Scores were evaluated using Cox models adjusted for age, sex, and ILD status, with additional models adjusted for baseline FVC % predicted among participants with available FVC measurements. Hazard ratios were reported per 1-SD increase using reference standard deviations of 3.63 for the 5-protein score and 1.31 for the 10-protein weighted score.

Model discrimination was summarized using C-statistics and internally validated using 1,000 bootstrap resamples with correction for optimism. The 5-protein score was held fixed during bootstrapping. For the 10-protein score, protein weights and the resulting score were re-estimated within each bootstrap sample because the weights were derived from the survival outcome.

Each of the 10 candidate proteins was additionally categorized into quartiles to visualize potential nonlinear relationships. Survival was estimated using Kaplan–Meier methods and compared using log-rank tests. These analyses were exploratory and were not adjusted for multiple comparisons.

All analyses were performed using R version 4.6.0 (R Foundation for Statistical Computing).

**Supplementary Results**

**Detailed proteomic cluster comparisons**

Relative to healthy controls, 89, 83, 102, and 133 proteins were differentially expressed in Clusters 1 through 4, respectively. Thus, the lower relative protein abundance observed in Cluster 1 should not be interpreted as a normal proteomic profile; all four clusters remained substantially different from healthy controls. Pairwise cluster comparisons showed the greatest separation between Clusters 1 and 4, with 175 proteins higher in Cluster 4, and between Clusters 2 and 4, with 140 proteins higher in Cluster 4.

Cluster 1 had the lowest overall abundance of disease-associated proteins among the four clusters but remained substantially different from healthy controls, with 89 differentially expressed proteins. Cluster 2 showed an intermediate immune/cytokine activation pattern, with FGF21, TNF-β, Flt3L, CCL19, IL-12B, TNFRSF9, and CCL4 increased relative to Cluster 1 and/or controls, but without the magnitude of injury-stress activation observed in Cluster 3 or the broad chemokine/checkpoint activation observed in Cluster 4.

Among 117 anti-Jo-1-positive participants with available data, quantitative anti-Jo-1 levels were higher in Clusters 3–4 combined (n=54; median raw level, 4,190,151 light units [LU]) than in Clusters 1–2 combined (n=63; median raw level, 2,334,124 LU; p<0.001; Supplementary Figure S9).

**Exploratory survival analyses of individual score components**

Kaplan–Meier curves were generated according to quartiles of each of the 10 candidate mortality-associated proteins (Supplementary Figure S11). Nominal survival differences were present for seven proteins, although the associations were not uniformly monotonic. OSM showed the strongest separation, driven primarily by poorer survival in the highest quartile. Higher TGF-α and NOS3 levels also generally identified groups with poorer survival, whereas lower CRH and TNF-β levels were associated with poorer outcomes. IL-7 and VEGFA showed significant overall quartile differences but less clearly ordered relationships. IL-6, IL-17C, and CD244 did not show significant overall quartile separation.

OSM, a component of both mortality scores, was also enriched in the injury-stress endotype, linking the survival and clustering analyses. These analyses were exploratory, used nominal log-rank p values, and were not adjusted for multiple comparisons.

**Exploratory longitudinal lung-function analyses**

Longitudinal pulmonary-function data were available for 64, 60, 51, and 50 patients at 6, 12, 18, and 24 months, respectively. Of these, 62, 56, 47, and 45 patients, respectively, could be matched to the Olink proteomic dataset and were included in the proteomic analyses. At 12 months, under the primary definition, 6 patients met criteria for decline, 18 for improvement, and 32 remained stable. Under the less stringent sensitivity definition, 9 patients met criteria for decline, 32 for improvement, and 15 remained stable.

Given the limited number of patients meeting decline criteria, these analyses were considered exploratory (Supplementary Figures S12–S15). Across complementary random forest and LASSO analyses, SERPINA9 was the most reproducible candidate marker of decline: it ranked first in the 6-month random forest analyses under both outcome definitions and was selected by LASSO under both 12-month definitions. NRTN and ADGRG1 were also selected under both 12-month definitions, whereas IL-7 and PON2 were selected under both 18-month definitions.

Improvement-associated signals were less stable. SCF and CXCL5 were selected under both 6-month definitions, while PLIN1 ranked among the leading 12-month random forest candidates under both definitions. No protein was consistently identified across all follow-up time points and outcome definitions. These findings identify candidate biomarkers of longitudinal lung-function change but require validation in larger longitudinal cohorts.

**Supplementary Figures**

**Supplementary Figure S1. Differentially expressed proteins in lower-ILD-risk autoantibody groups compared with controls.**

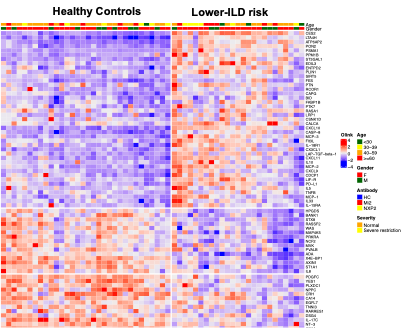

Heatmap of differentially expressed serum proteins comparing patients with lower-ILD-risk autoantibodies with matched healthy controls. Columns represent individual participants and rows represent proteins identified as differentially expressed in this group-level comparison. Protein abundance is shown as normalized Olink NPX values scaled by protein, with higher relative expression shown in red and lower relative expression shown in blue. Column annotations indicate age group, sex, autoantibody type, and ILD severity. Lower-ILD-risk autoantibody groups included anti-Mi-2 and anti-NXP2.
Abbreviations: ILD, interstitial lung disease; NPX, normalized protein expression

The TIF1γ participant was included in the corresponding group-level analyses but is not displayed separately due to the small sample size.

**Supplementary Figure S2. Differentially expressed proteins in higher-ILD-risk autoantibody patients with ILD compared with controls.**

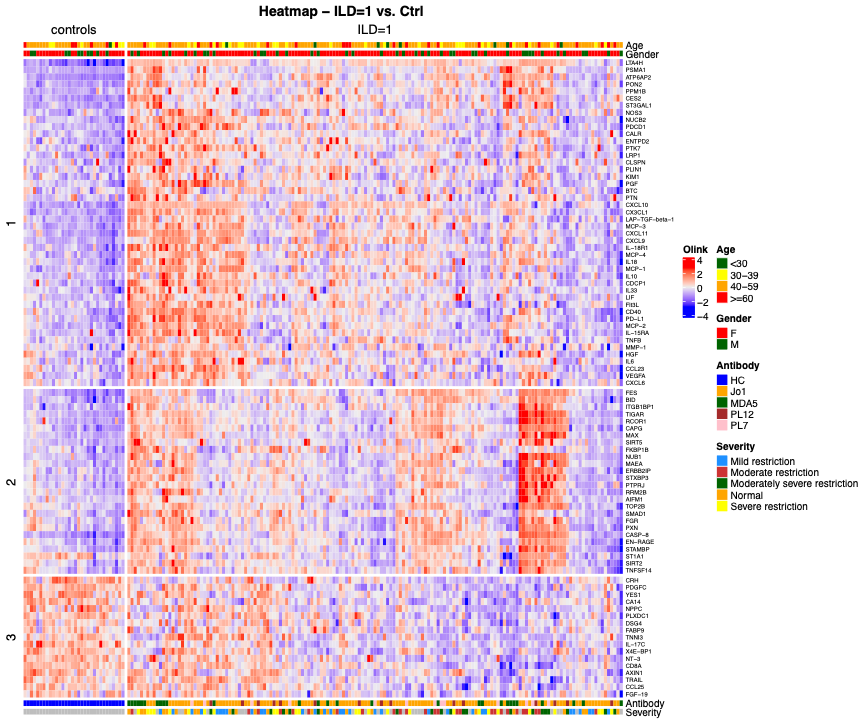

Heatmap of differentially expressed serum proteins comparing higher-ILD-risk autoantibody patients with ILD with matched healthy controls. Columns represent individual participants and rows represent proteins identified as differentially expressed in this comparison. Column annotations indicate age group, sex, autoantibody type, and ILD severity. Protein abundance is shown as normalized Olink NPX values scaled by protein, with higher relative expression shown in red and lower relative expression shown in blue.

Abbreviations: ILD, interstitial lung disease; NPX, normalized protein expression.

**Supplementary Figure S3. Differentially expressed proteins in higher-ILD-risk autoantibody patients without ILD compared with controls.**

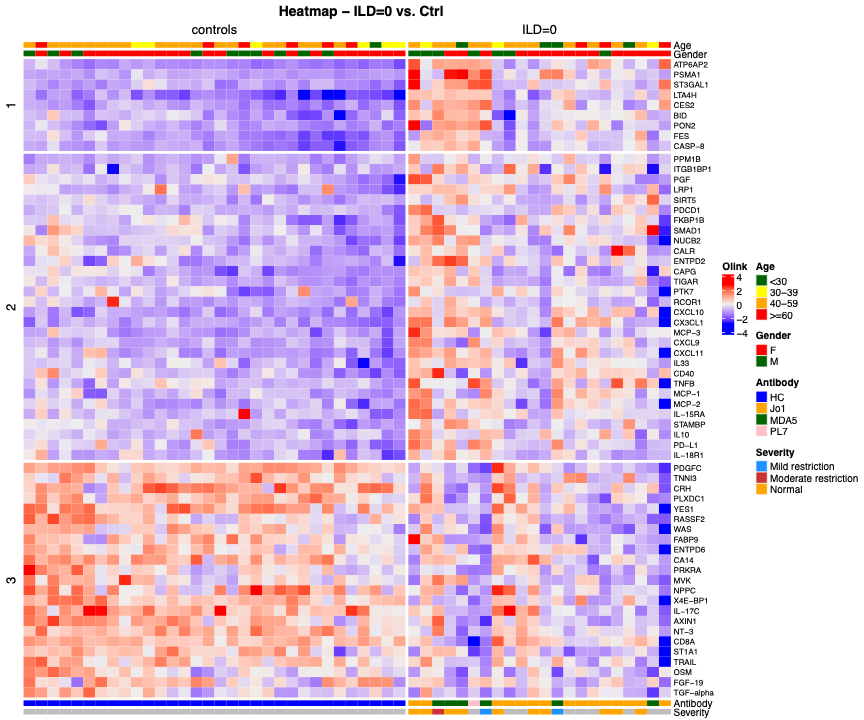

Heatmap of differentially expressed serum proteins comparing higher-ILD-risk autoantibody patients without ILD with matched healthy controls. Columns represent individual participants and rows represent proteins identified as differentially expressed in this comparison. Column annotations indicate age group, sex, autoantibody type, and ILD severity. Protein abundance is shown as normalized Olink NPX values scaled by protein, with higher relative expression shown in red and lower relative expression shown in blue.
Abbreviations: ILD, interstitial lung disease; NPX, normalized protein expression.

**Supplementary Figure S4. Hallmark pathway enrichment analysis of differentially expressed proteins in myositis autoantibody groups compared with controls.**

**A**

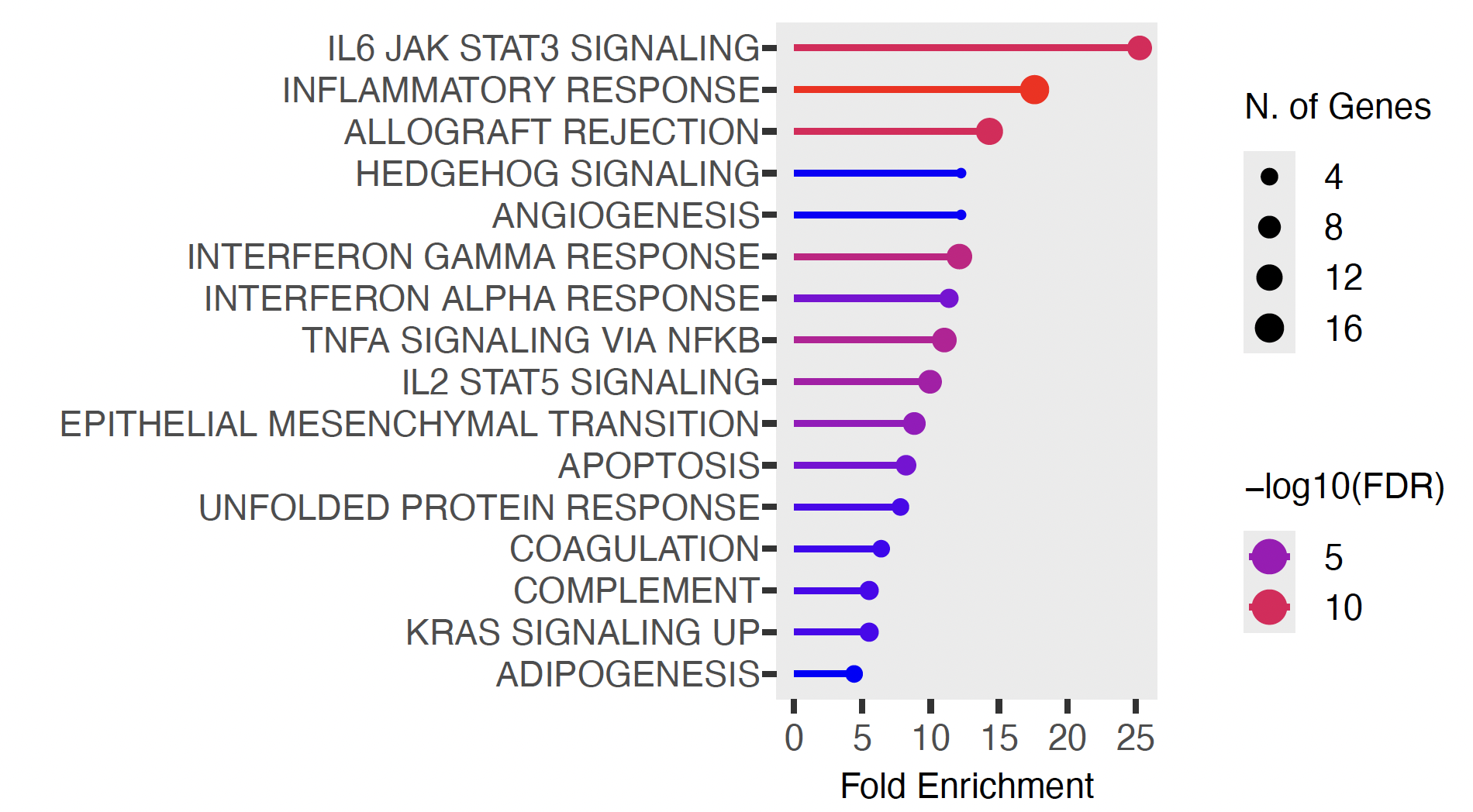

**B**

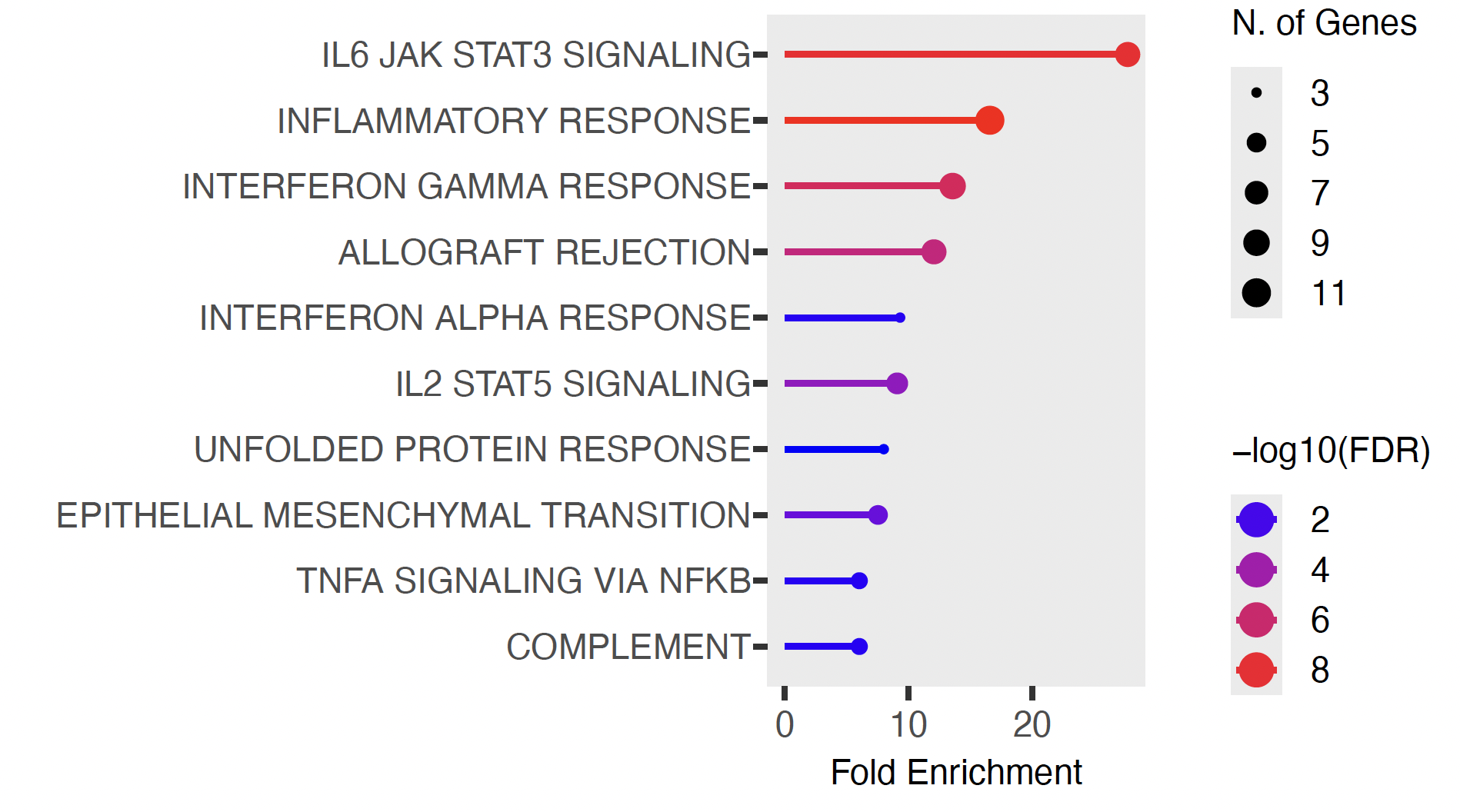

Hallmark pathway enrichment analysis was performed using differentially expressed proteins from comparisons of (A) higher-ILD-risk autoantibody groups versus healthy controls and (B) lower-ILD-risk autoantibody groups versus healthy controls. Pathways are ranked by fold enrichment. Point size represents the number of genes/proteins contributing to each pathway, and color indicates statistical significance as −log10 false discovery rate. Both higher-ILD-risk and lower-ILD-risk autoantibody groups showed enrichment of IL-6/JAK/STAT3 signaling, inflammatory response, interferon-related signaling, and TNF/NF-κB signaling, supporting a shared inflammatory pathway architecture across myositis autoantibody groups. Abbreviations: FDR, false discovery rate; ILD, interstitial lung disease.

**Supplementary Figure S5. Ingenuity Pathway Analysis of higher-ILD-risk group compared with healthy controls.**

A. Top Canonical Pathways

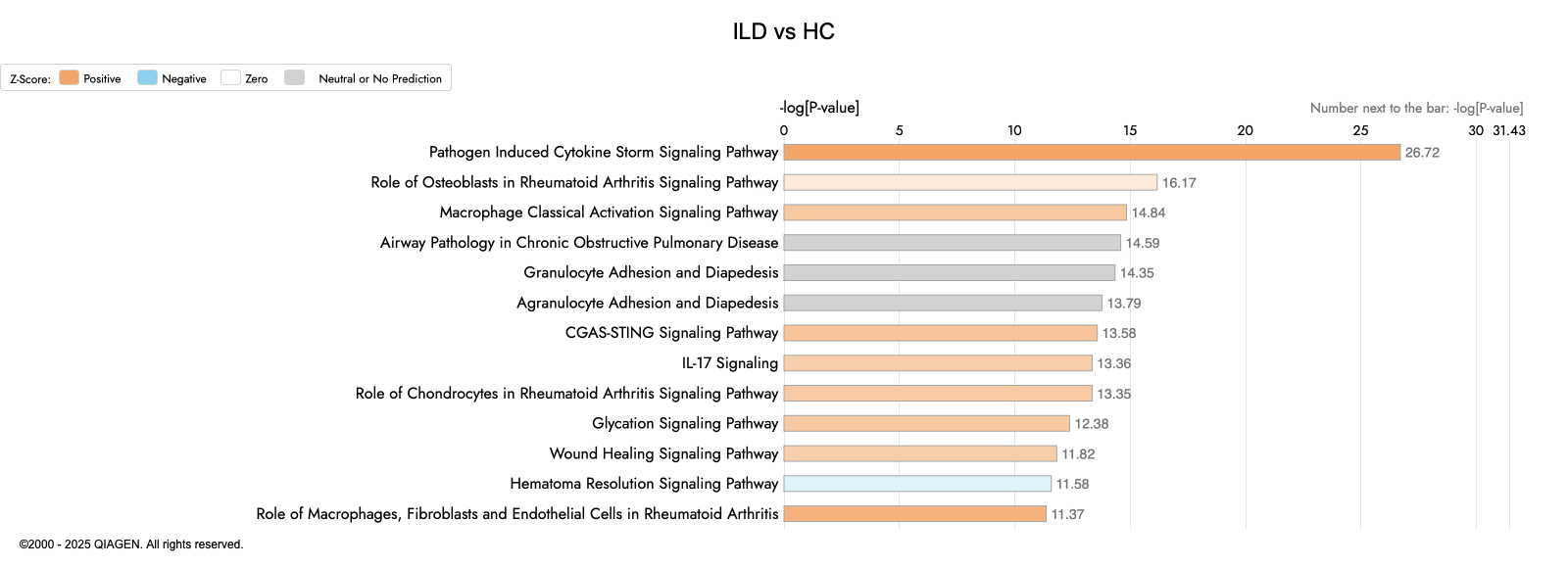

B. Graphical Summary

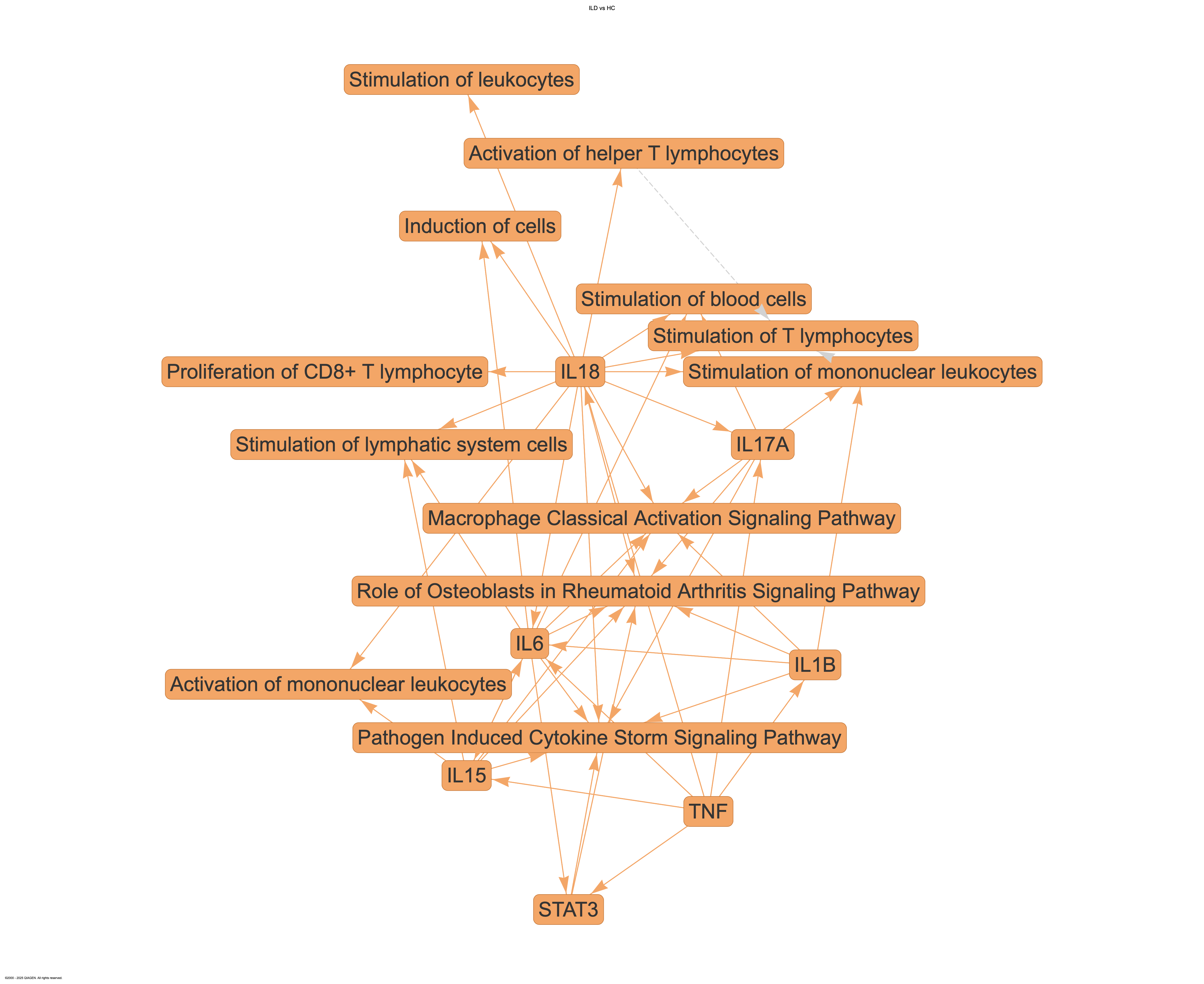

**Legend**

(A) Top canonical pathways identified by Ingenuity Pathway Analysis among differentially expressed proteins in the higher-ILD risk autoantibody group compared with healthy controls. Pathways are ranked by −log10(P value).

(B) Graphical summary integrating the principal molecules, canonical pathways, and predicted biological functions identified in this comparison. The network highlights cytokine-mediated immune activation involving IL-6, IL-15, IL-17A, IL-18, IL-1β, TNF, and STAT3, together with macrophage classical activation and pathogen-induced cytokine-storm signaling. Orange indicates predicted activation and blue indicates predicted inhibition.

**Supplementary Figure S6. Ingenuity Pathway Analysis of lower-ILD risk group compared with healthy controls.**

A. Top Canonical Pathways

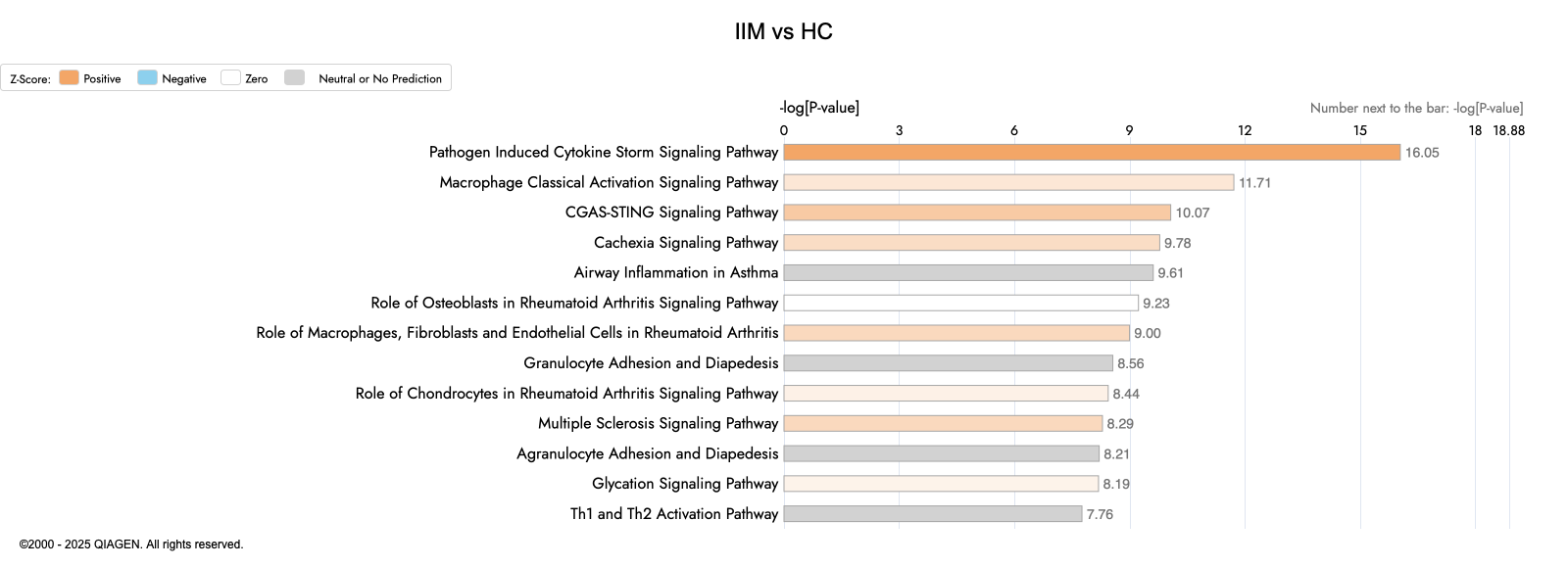

B. Graphical Summary

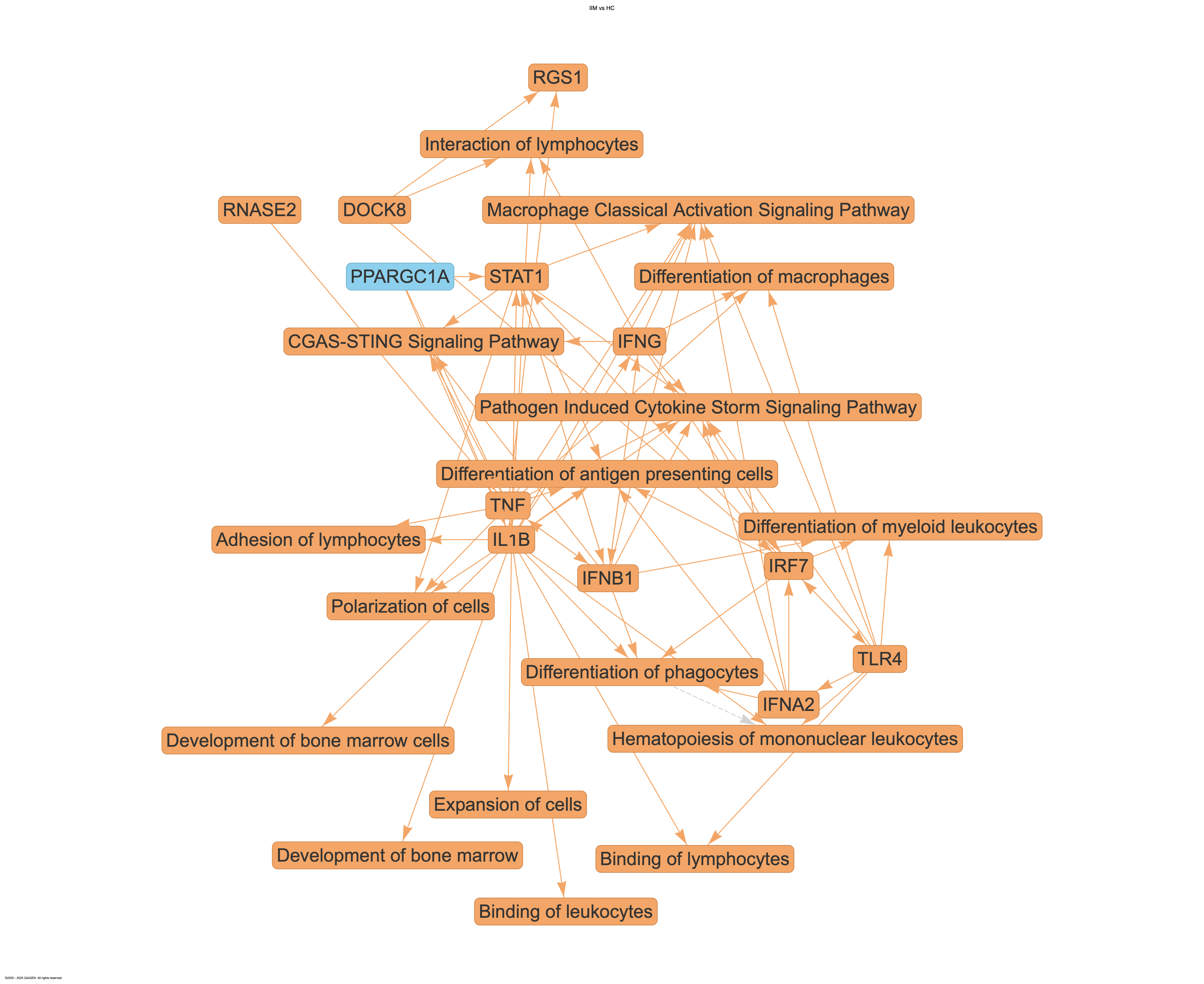

A. Top canonical pathways identified by Ingenuity Pathway Analysis among differentially expressed proteins in the lower-ILD risk autoantibody group compared with healthy controls. Pathways are ranked by −log10(P value).

B. Graphical summary integrating the principal molecules, canonical pathways, and predicted biological functions identified in this comparison. The network highlights interferon-related and innate immune signaling involving IFNG, IFNB1, IFNA2, STAT1, IRF7, TLR4, TNF, and IL-1β, together with macrophage activation, antigen-presenting-cell differentiation, and leukocyte development and adhesion. Orange indicates predicted activation and blue indicates predicted inhibition.

**Supplementary Figure S7. Correlations between quantitative autoantibody levels and baseline pulmonary function.**

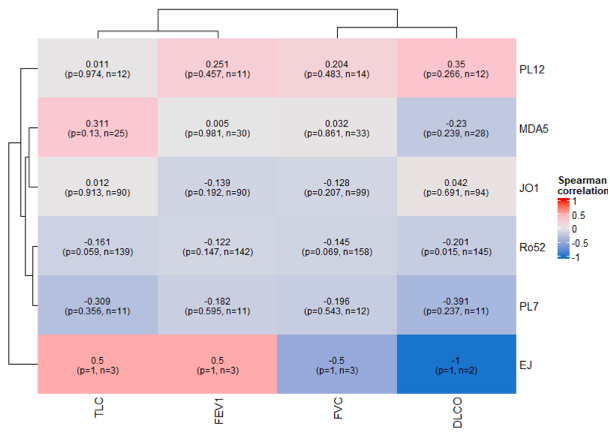

Heatmap showing Spearman correlations between quantitative autoantibody levels and baseline pulmonary function measures, including TLC, FEV1, FVC, and DLCO. Rows represent autoantibody groups and columns represent pulmonary function variables. Values within cells indicate Spearman correlation coefficients, with corresponding nominal P values and sample sizes shown in parentheses. Red indicates positive correlation and blue indicates negative correlation. Overall, quantitative autoantibody levels showed limited correlation with baseline pulmonary function, although Ro52 showed weak inverse correlations with pulmonary function measures, including DLCO. Abbreviations: DLCO, diffusing capacity of the lung for carbon monoxide; FEV1, forced expiratory volume in 1 second; FVC, forced vital capacity; TLC, total lung capacity.

**Supplementary Figure S8. Targeted heatmap of representative proteins across proteomic clusters.**

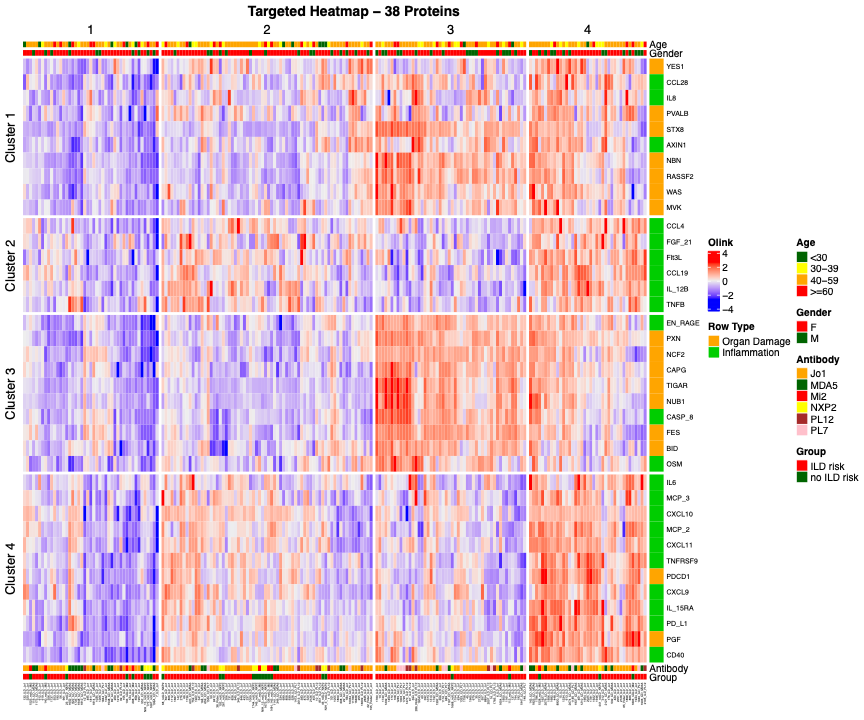

Heatmap showing relative expression of 38 representative serum proteins across the four unsupervised proteomic clusters. Participants are arranged by cluster, and proteins are grouped according to the proteomic program they most strongly represented. Colors indicate scaled Olink normalized protein expression values, with red representing higher and blue representing lower relative protein abundance. Annotation bars indicate age group, sex, myositis-specific autoantibody, and higher- versus lower-ILD-risk autoantibody group. The 38 proteins were selected based on cluster-associated differential expression and biological relevance to illustrate the dominant molecular programs identified in each endotype; they were not used to derive the clusters.

**Supplementary Figure S9. Quantitative anti-Jo-1 levels across proteomic clusters.**

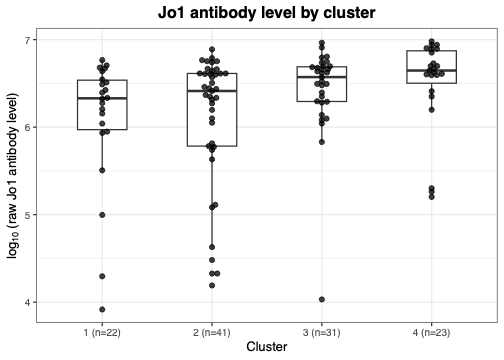

Boxplots show log10-transformed raw anti-Jo-1 levels among 117 anti-Jo-1-positive participants stratified by proteomic cluster: Cluster 1, n=22; Cluster 2, n=41; Cluster 3, n=31; and Cluster 4, n=23. Points represent individual participants, boxes represent the interquartile range, and horizontal lines indicate median values. In an exploratory comparison, anti-Jo-1 levels were higher in Clusters 3–4 combined than in Clusters 1–2 combined. Median raw anti-Jo-1 levels were 4,190,151 in Clusters 3–4 and 2,334,124 in Clusters 1–2. P=0.00069 by Wilcoxon rank-sum test of log10-transformed anti-Jo-1 levels.

**Supplementary Figure S10. Random survival forest variable-importance analysis for all-cause mortality.**

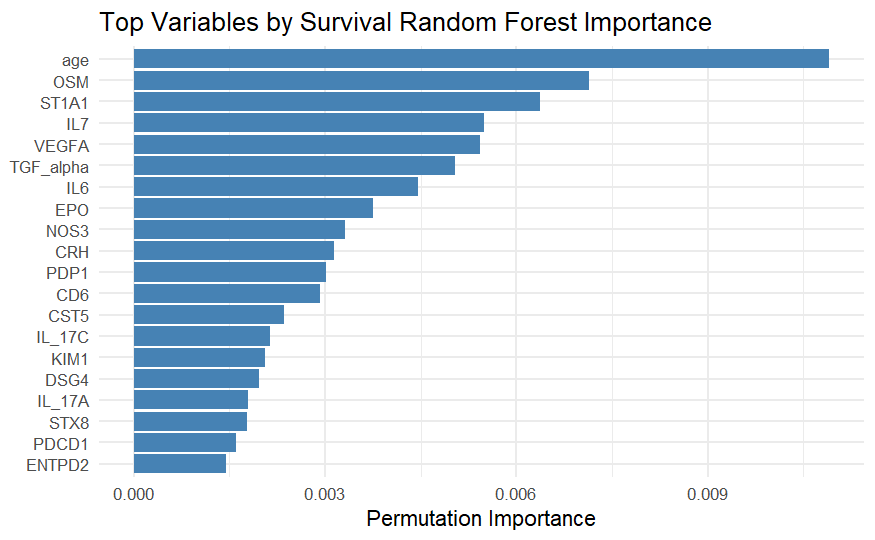

A random survival forest model was used as a complementary, non-parametric approach to prioritize variables associated with all-cause mortality. The bar plot shows the top-ranked variables according to predictive importance, with higher values indicating greater contribution to model prediction. Age was the highest-ranked variable, followed by several proteins also identified in Cox proportional hazards models, including OSM, IL-7, VEGFA, TGF-α, IL-6, CRH, PDP1, CD6, IL-17C, IL-17A, STX8, and PDCD1. These results were used to support exploratory prioritization of candidate mortality-associated proteins and composite protein-score construction.

**Supplementary Figure S11. All-cause survival according to quartiles of candidate mortality-associated proteins.**

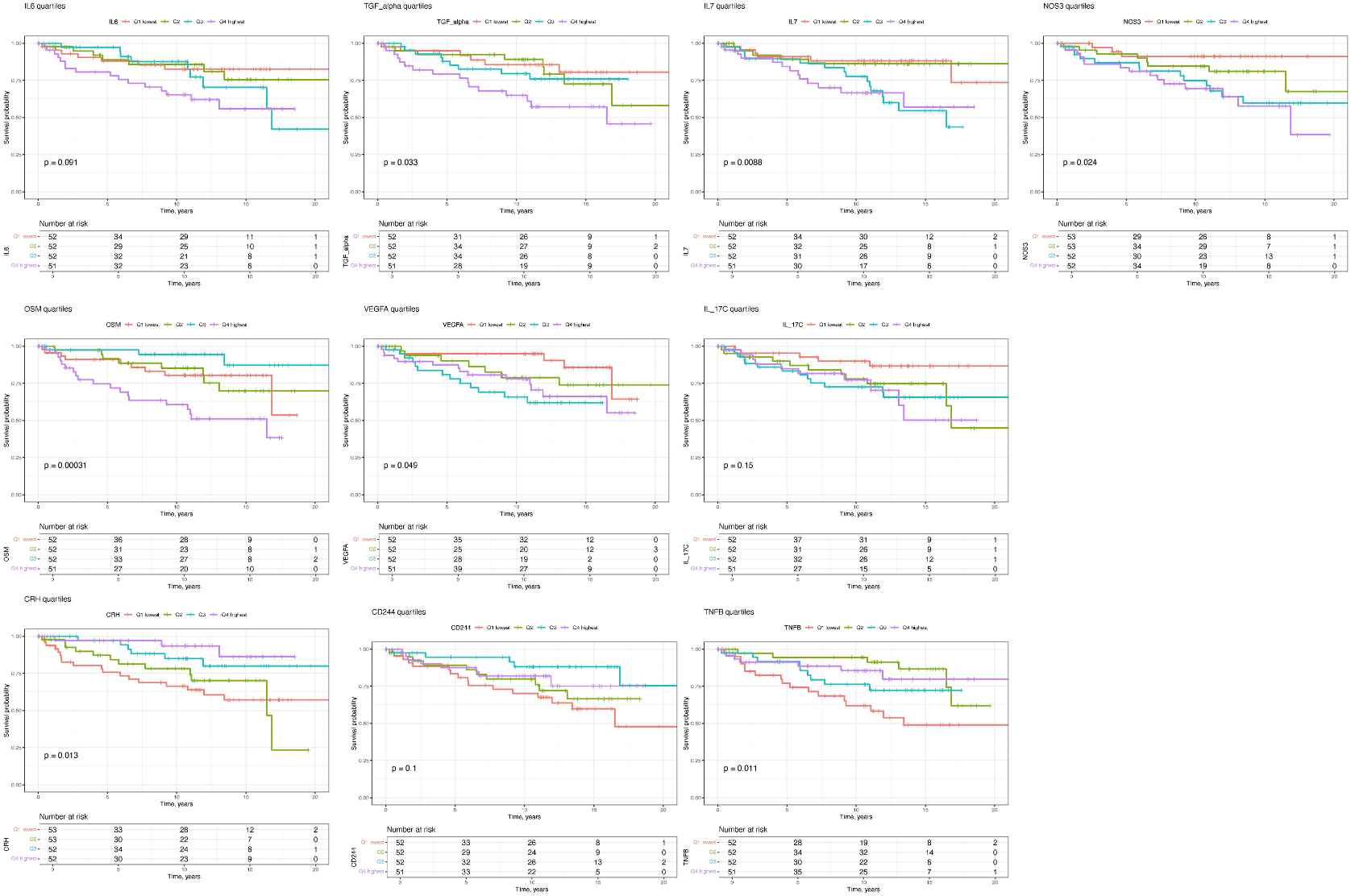

Kaplan-Meier curves show all-cause survival stratified by quartiles of baseline IL-7, OSM, VEGFA, IL-6, TGF-α, CRH, IL-17C, TNF-β, CD244, and NOS3 abundance. Q1 represents the lowest and Q4 the highest protein-abundance quartile. Analyses included participants with complete survival data and a non-missing measurement for the protein shown; 207 participants were included for IL-7, OSM, VEGFA, IL-6, TGF-α, IL-17C, TNF-β, and CD244, and 210 participants for CRH and NOS3. Tick marks indicate censored observations, and tables show the number of participants at risk. P values were calculated using unadjusted log-rank tests comparing survival across the four quartiles. These exploratory analyses were not adjusted for multiple comparisons and were performed to visualize potential nonlinear associations between individual proteins and survival.

**Supplementary Figure S12. Random forest variable-importance analysis for lung-function decline at 6 and 12 months.**

A. Standard definition - 6 months

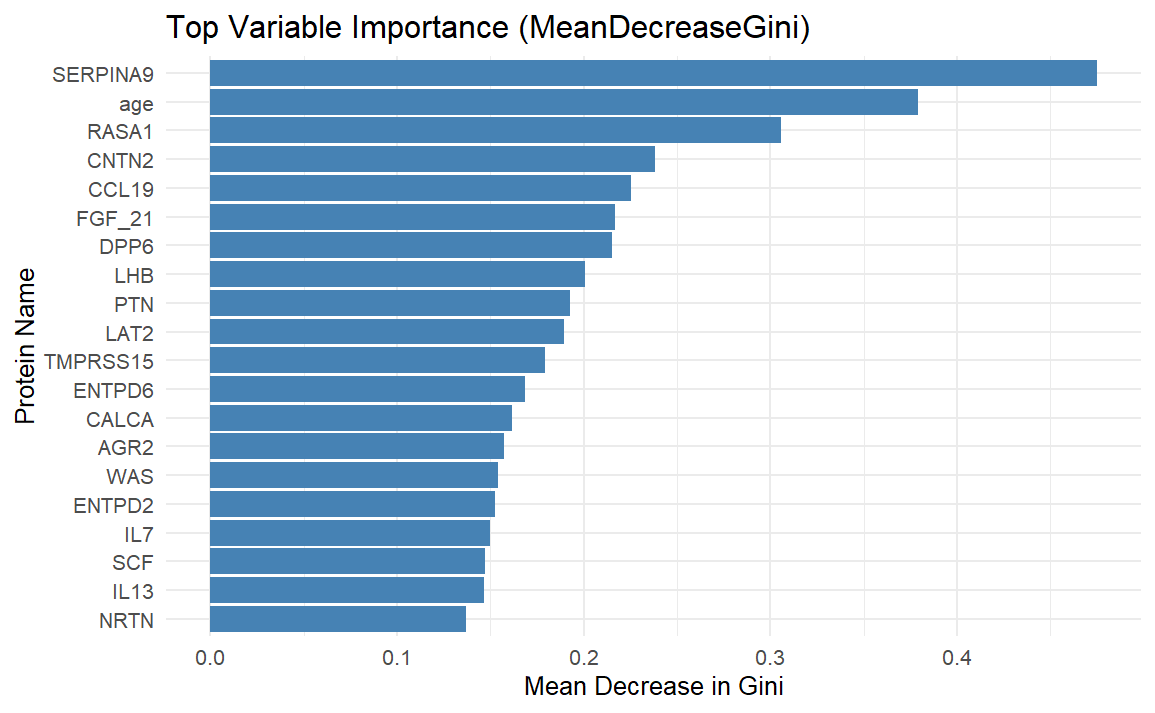

B. Less stringent definition - 6 months

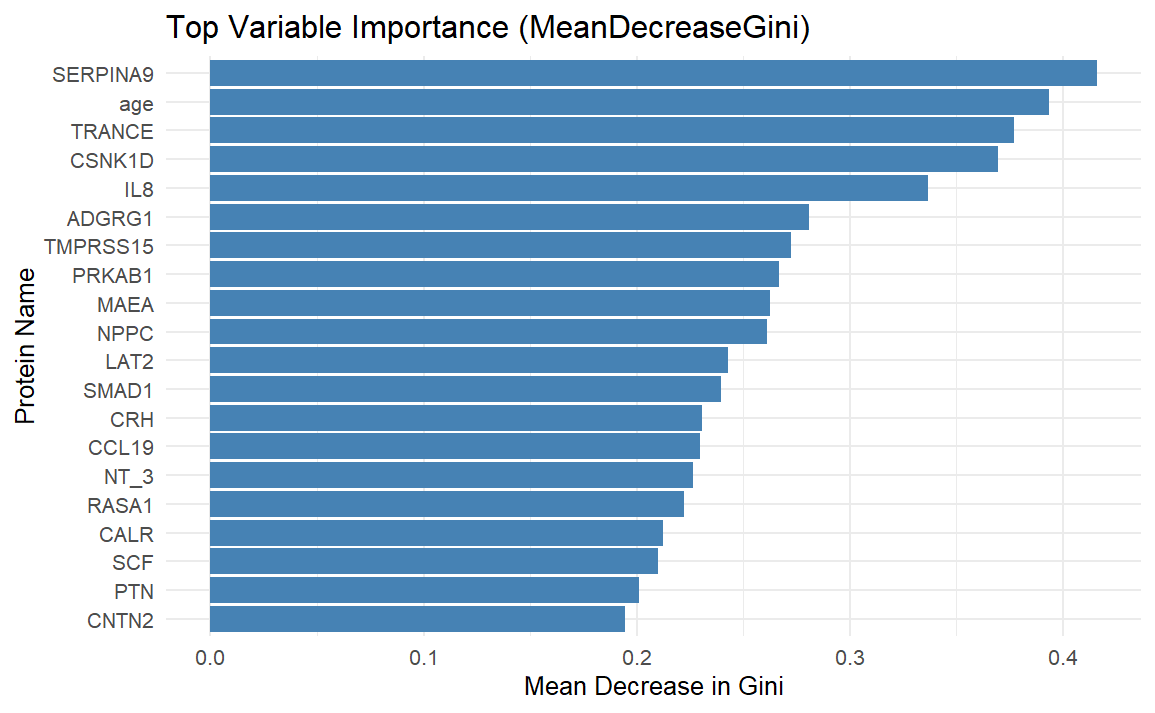

C. Standard definition - 12 months

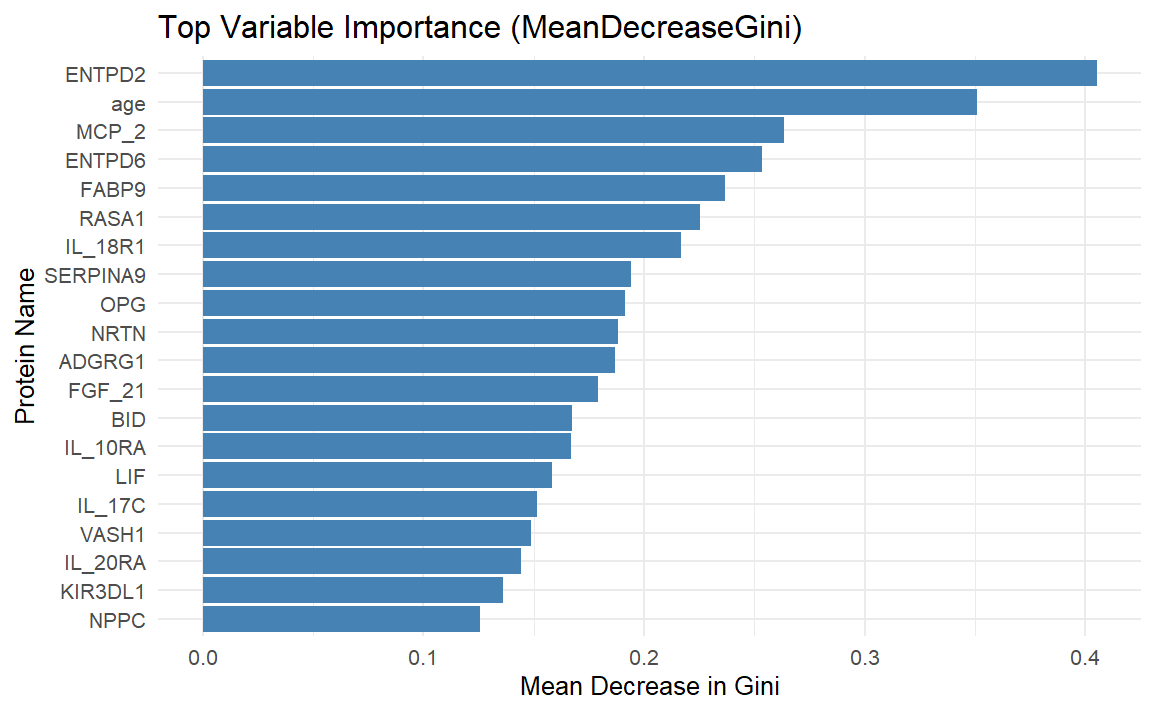

D. Less stringent definition - 12 months

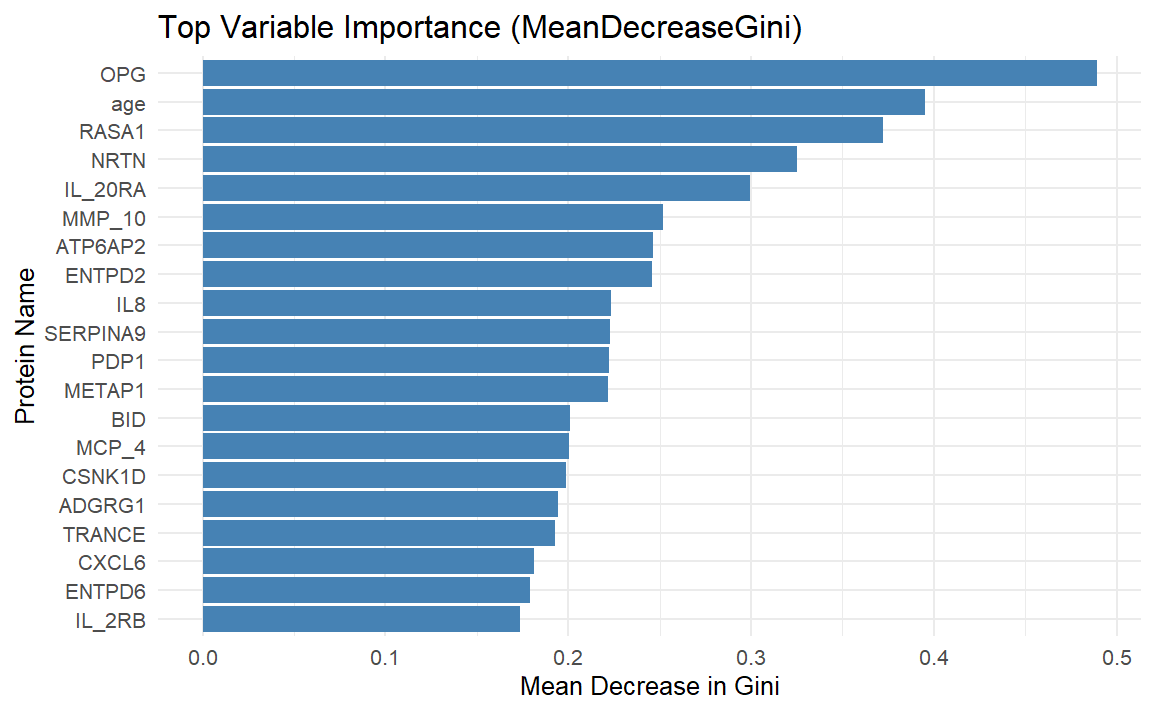

Random forest models evaluated baseline variables associated with lung-function decline versus all lung function categories at 6 months (A, B) and 12 months (C, D). Panels A and C use the standard definition: an absolute decrease from baseline of at least 10 percentage points in forced vital capacity (FVC) % predicted and/or at least 15 percentage points in diffusing capacity of the lung for carbon monoxide (DLCO) % predicted. Panels B and D use the less stringent definition: an absolute decrease of at least 5 percentage points in FVC % predicted and/or at least 10 percentage points in DLCO % predicted. Bars show mean decrease in Gini impurity (MeanDecreaseGini), a relative variable-importance measure; higher values indicate greater contribution to classification within the corresponding model but do not indicate the direction of association, effect size, or statistical significance. Analyses were exploratory.

**Supplementary Figure S13. Random forest variable-importance analysis for lung-function decline at 18 and 24 months.**

A. Standard definition - 18 months

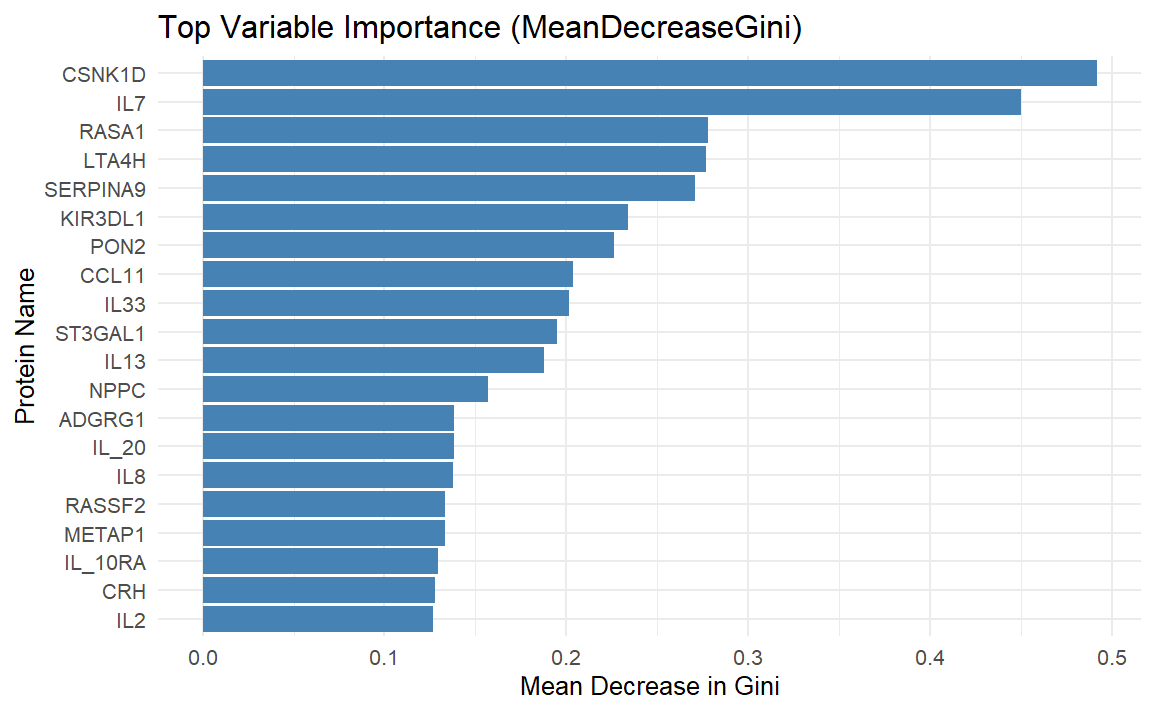

B. Less stringent definition - 18 months

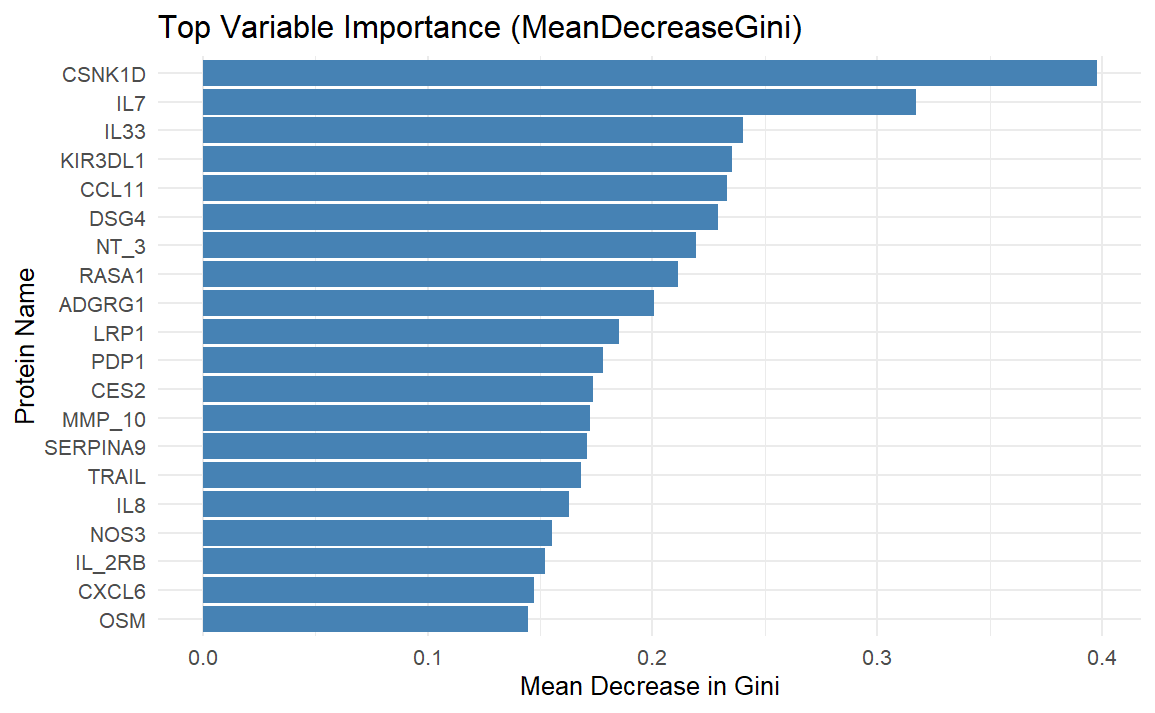

C. Standard definition - 24 months

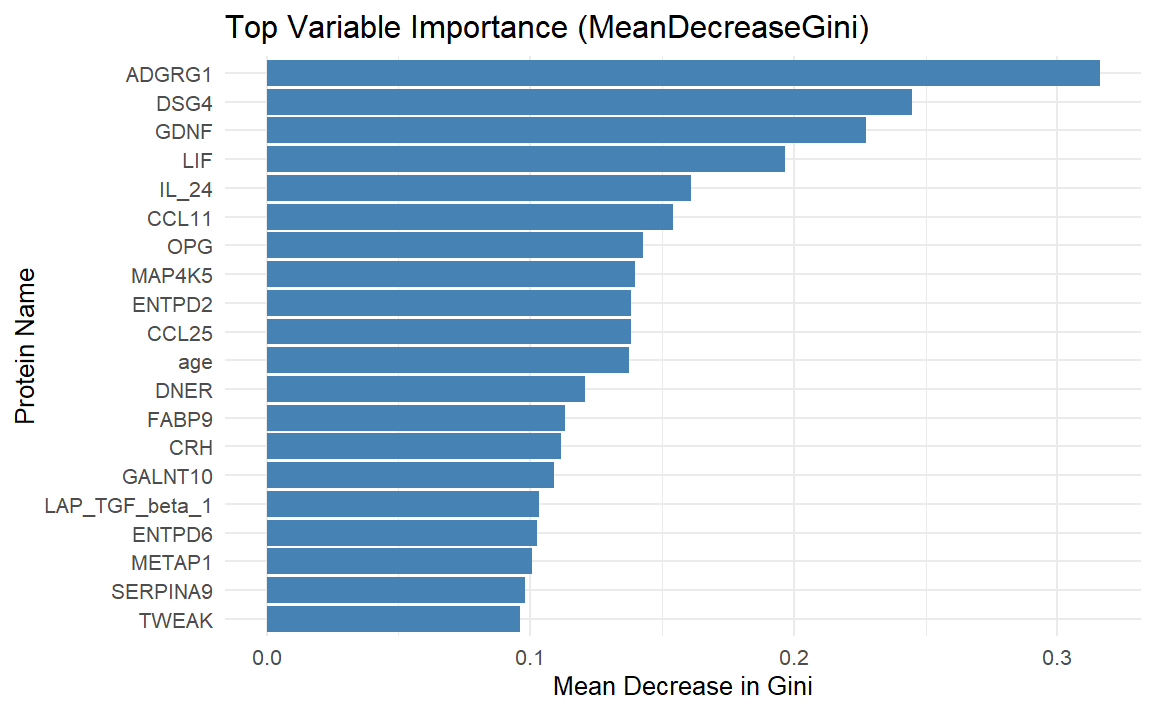

D. Less stringent definition - 24 months

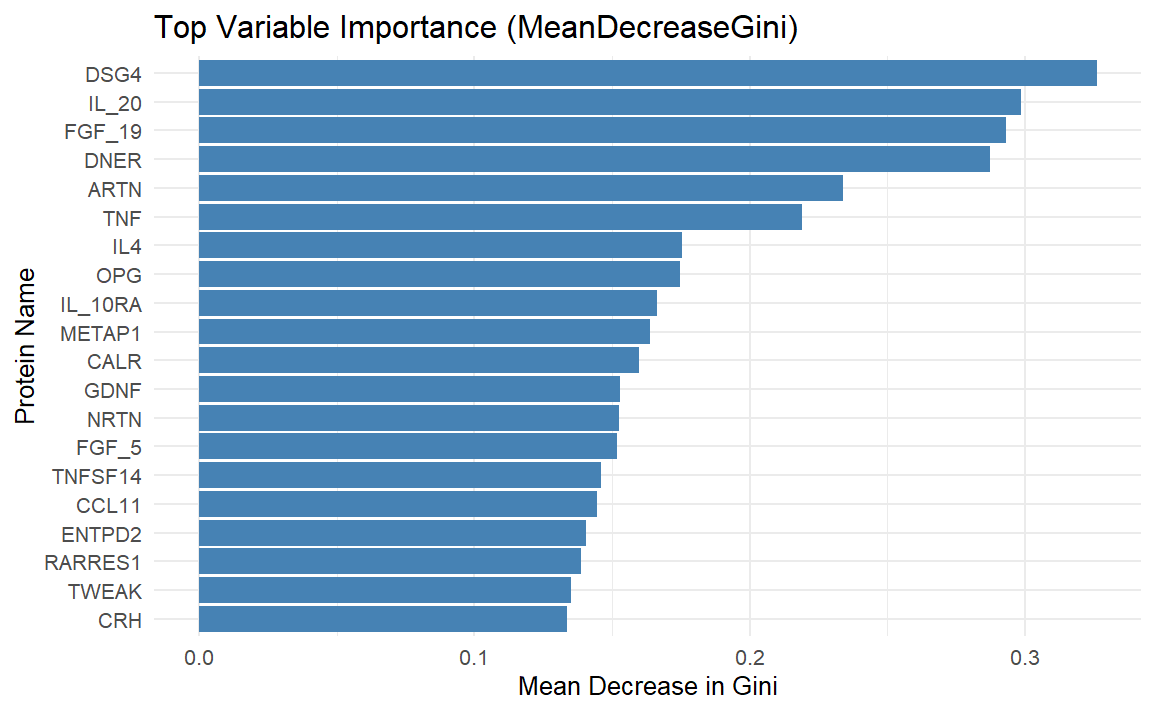

Random forest models evaluated baseline variables associated with lung-function decline versus all other lung-function categories at 18 months (A, B) and 24 months (C, D). Panels A and C use the standard definition: an absolute decrease from baseline of at least 10 percentage points in FVC % predicted and/or at least 15 percentage points in DLCO % predicted. Panels B and D use the less stringent definition: an absolute decrease of at least 5 percentage points in FVC % predicted and/or at least 10 percentage points in DLCO % predicted. Bars show mean decrease in Gini impurity (MeanDecreaseGini), a relative variable-importance measure; higher values indicate greater contribution to classification within the corresponding model but do not indicate the direction of association, effect size, or statistical significance. Analyses were exploratory.

**Supplementary Figure S14. Random forest variable-importance analysis for lung-function improvement at 6 and 12 months.**

A. Standard definition - 6 months

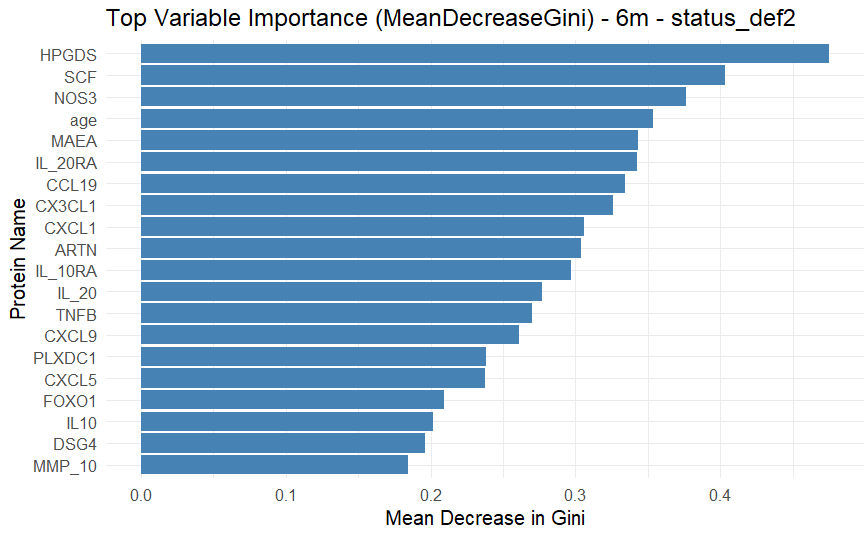

B. Less stringent definition - 6 months

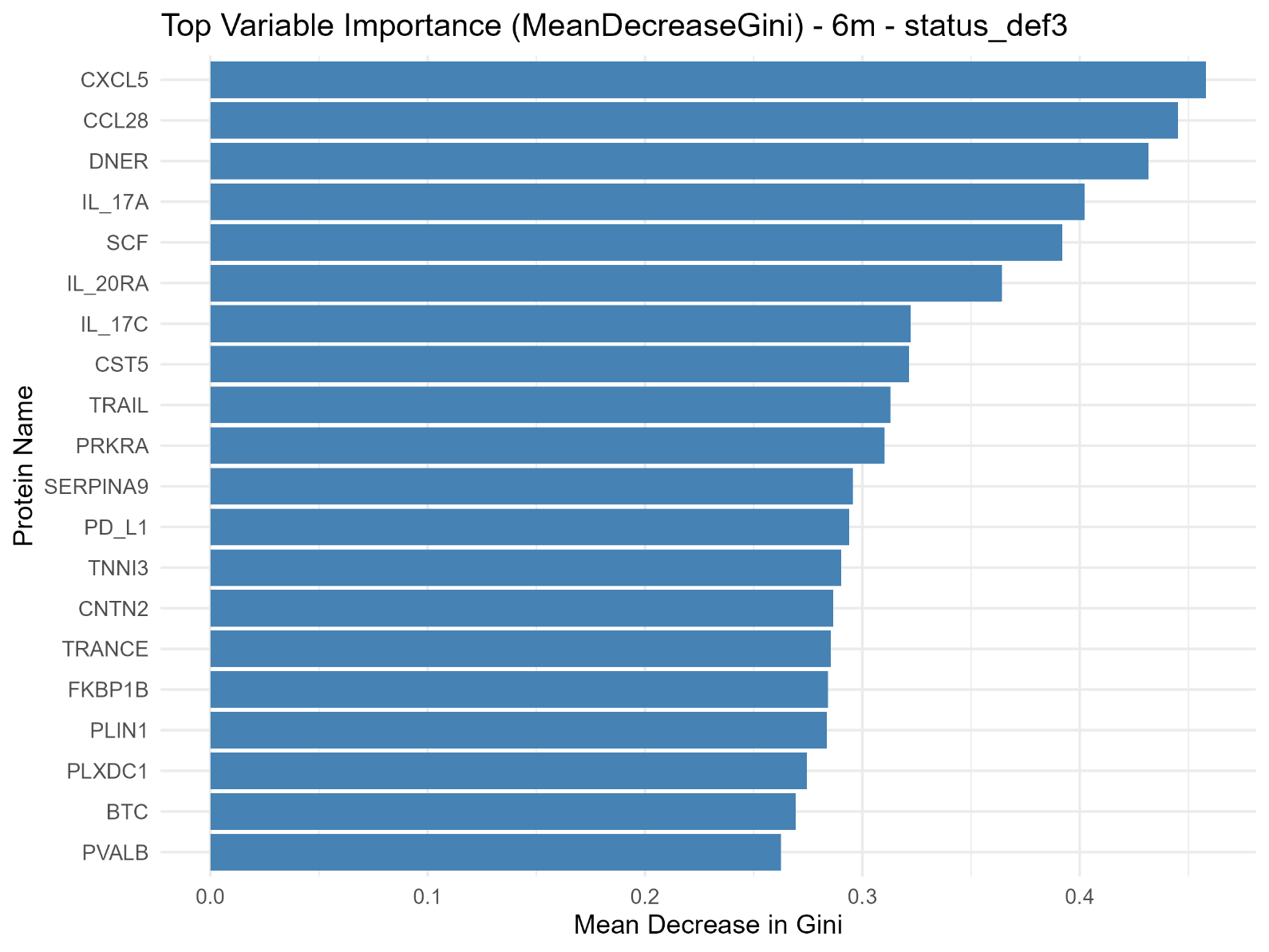

C. Standard definition - 12 months

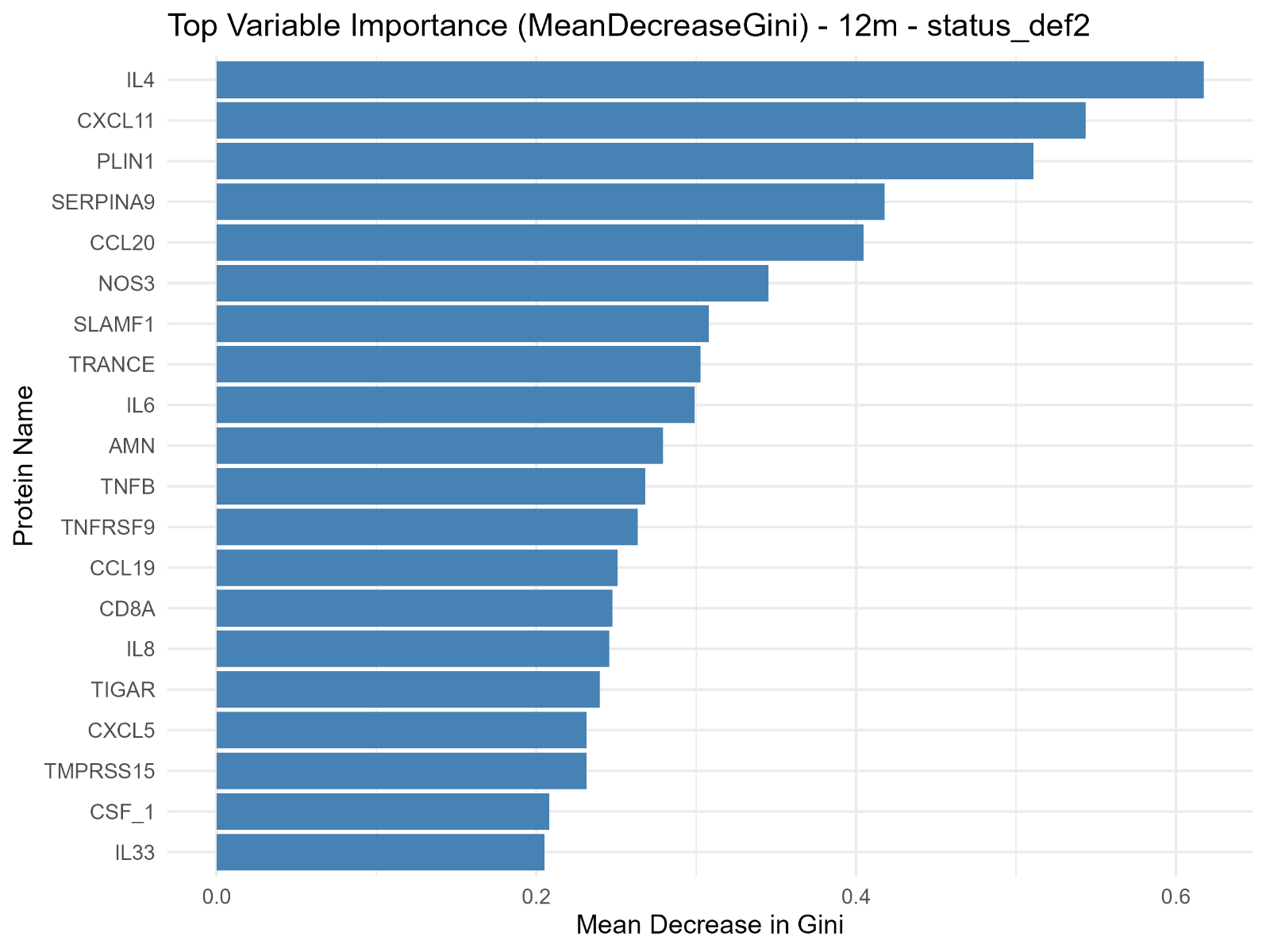

D. Less stringent definition - 12 months

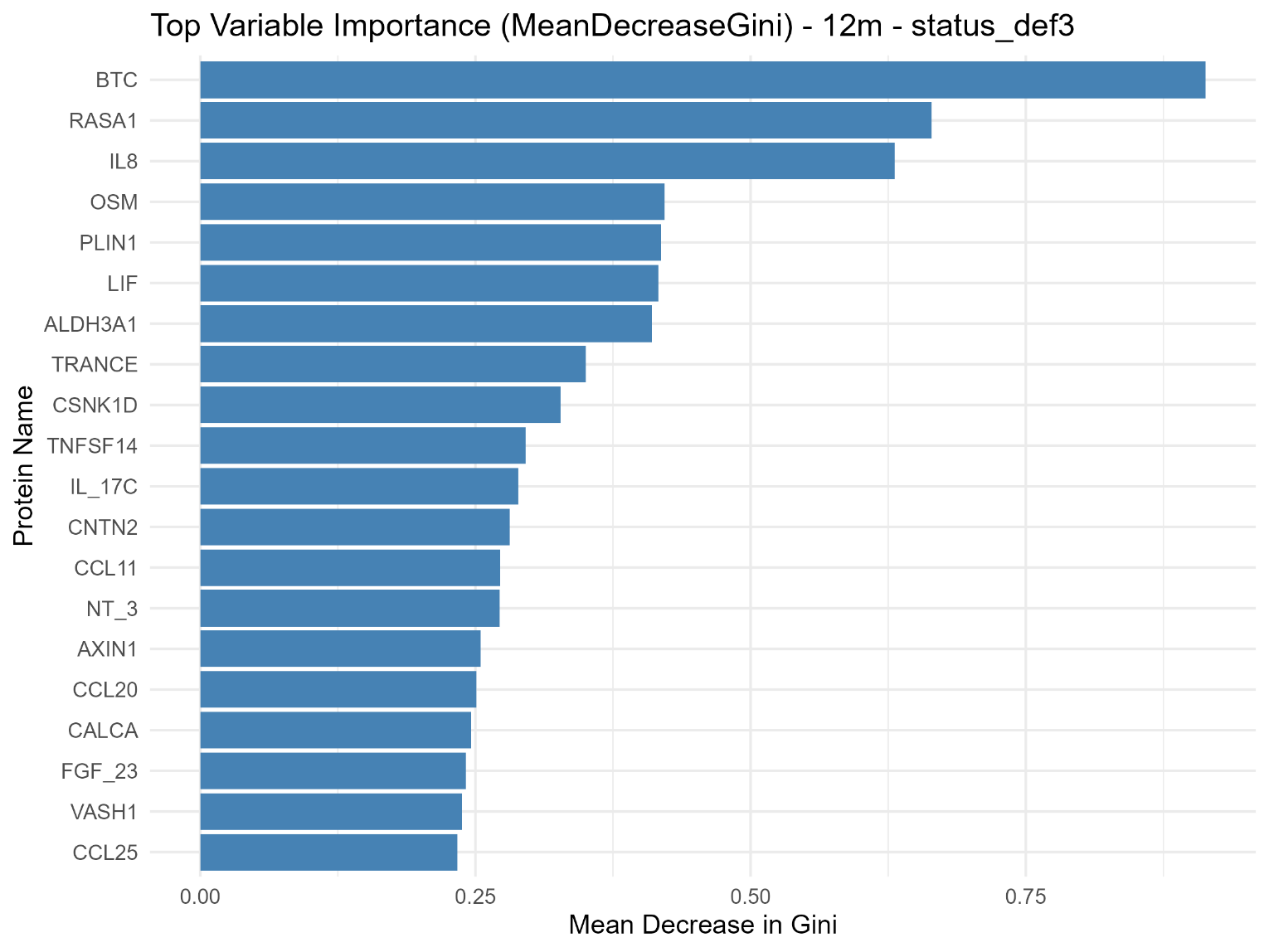

Random forest models evaluated baseline variables associated with lung-function improvement versus all other lung-function categories at 6 months (A, B) and 12 months (C, D). Panels A and C use the standard definition: an absolute increase from baseline of at least 10 percentage points in FVC % predicted and/or at least 15 percentage points in DLCO % predicted. Panels B and D use the less stringent definition: an absolute increase of at least 5 percentage points in FVC % predicted and/or at least 10 percentage points in DLCO % predicted. Bars show mean decrease in Gini impurity (MeanDecreaseGini), a relative variable-importance measure; higher values indicate greater contribution to classification within the corresponding model but do not indicate the direction of association, effect size, or statistical significance. Analyses were exploratory.

**Supplementary Figure S15. Random forest variable-importance analysis for lung-function improvement at 18 and 24 months.**

A. Standard definition - 18 months

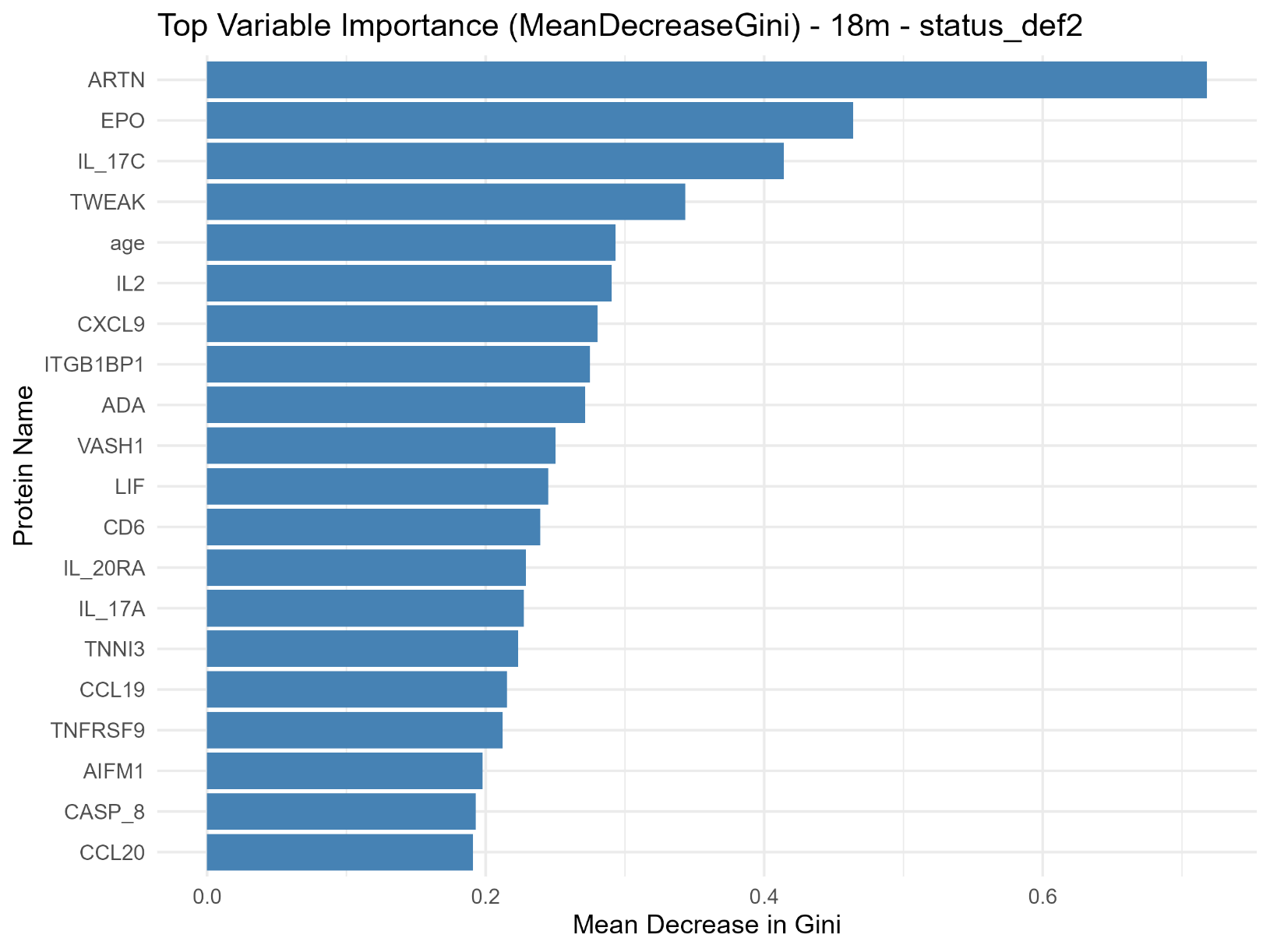

B. Less stringent definition - 18 months

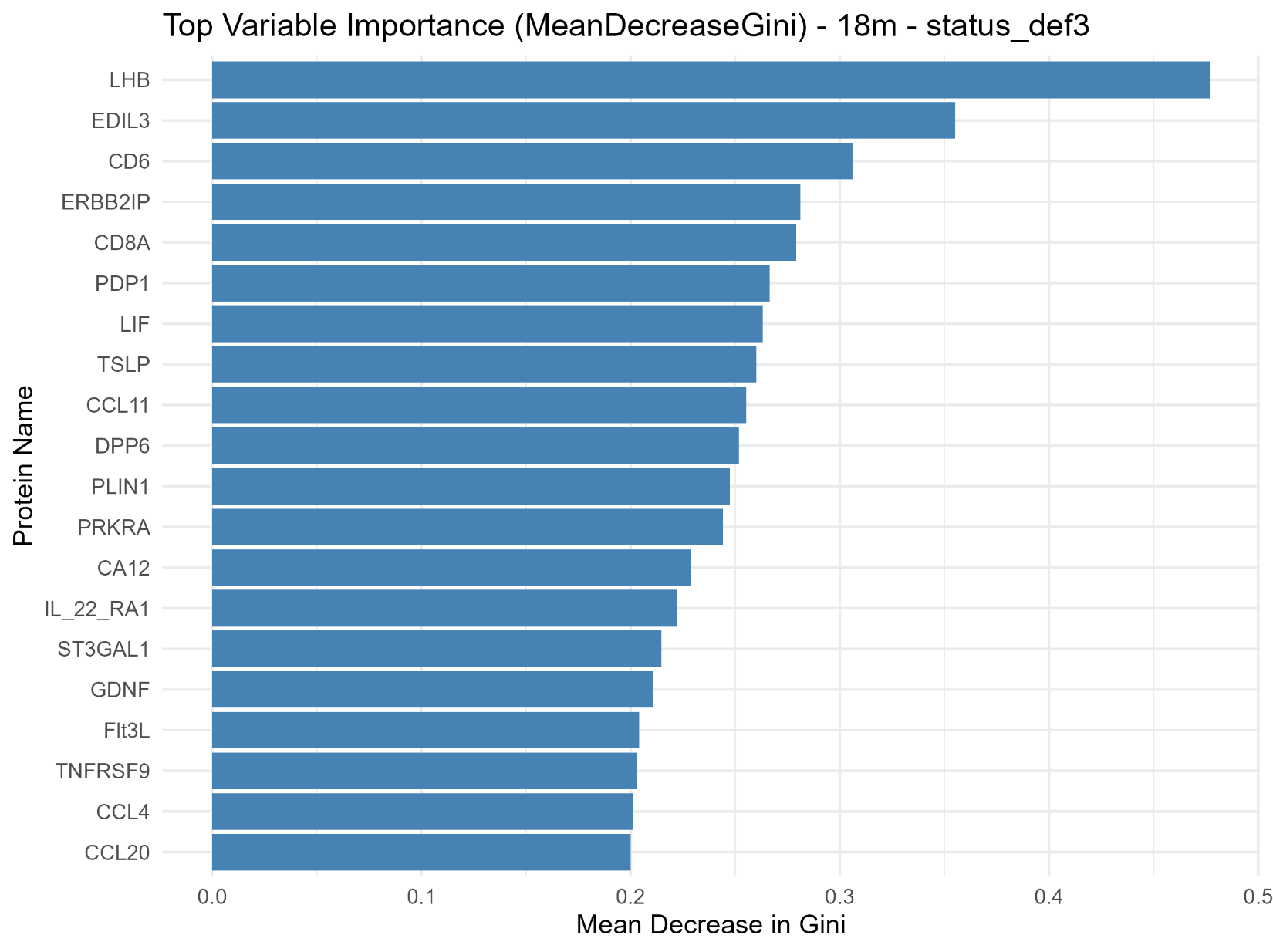

C. Standard definition - 24 months

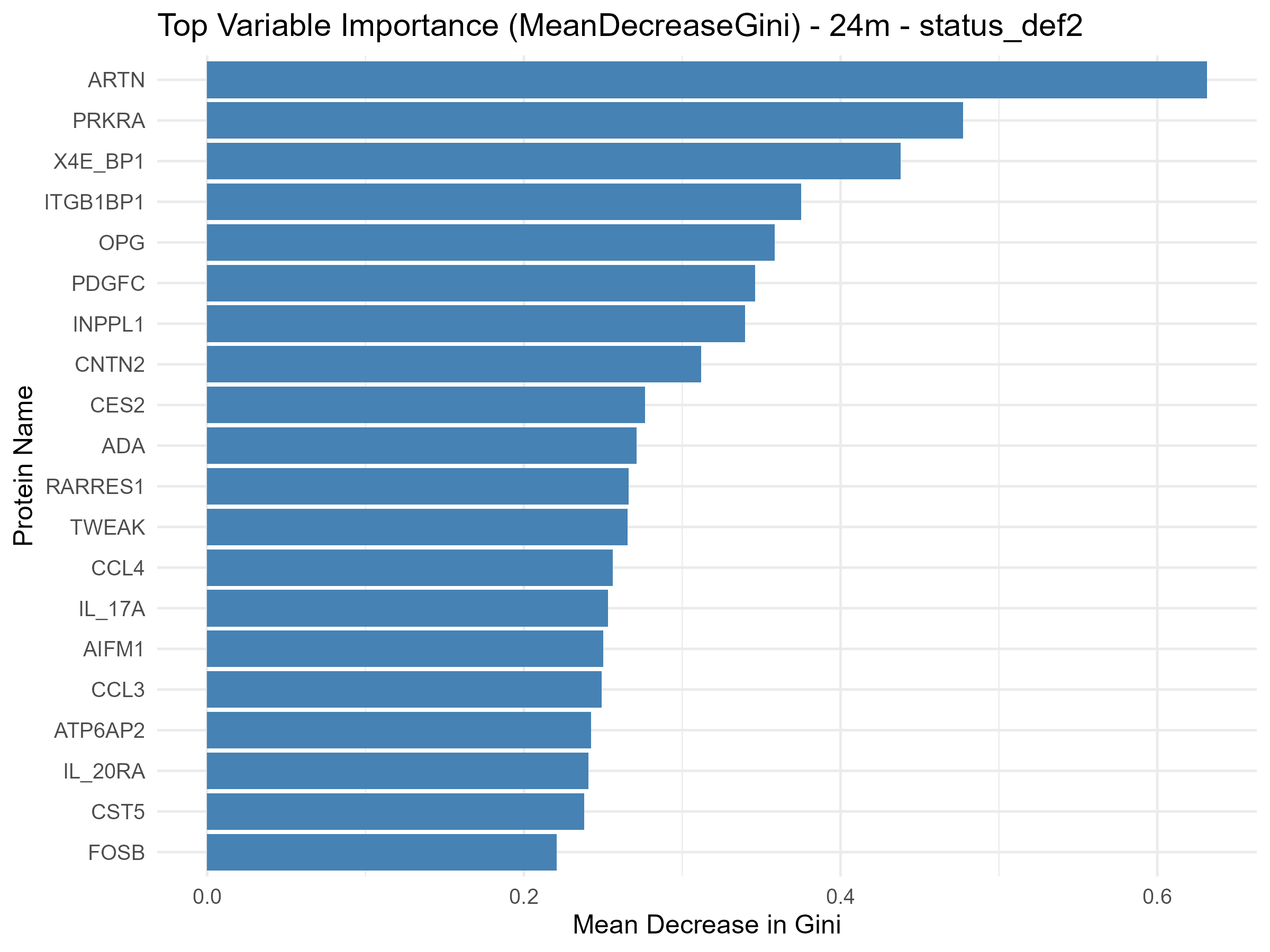

D. Less stringent definition - 24 months

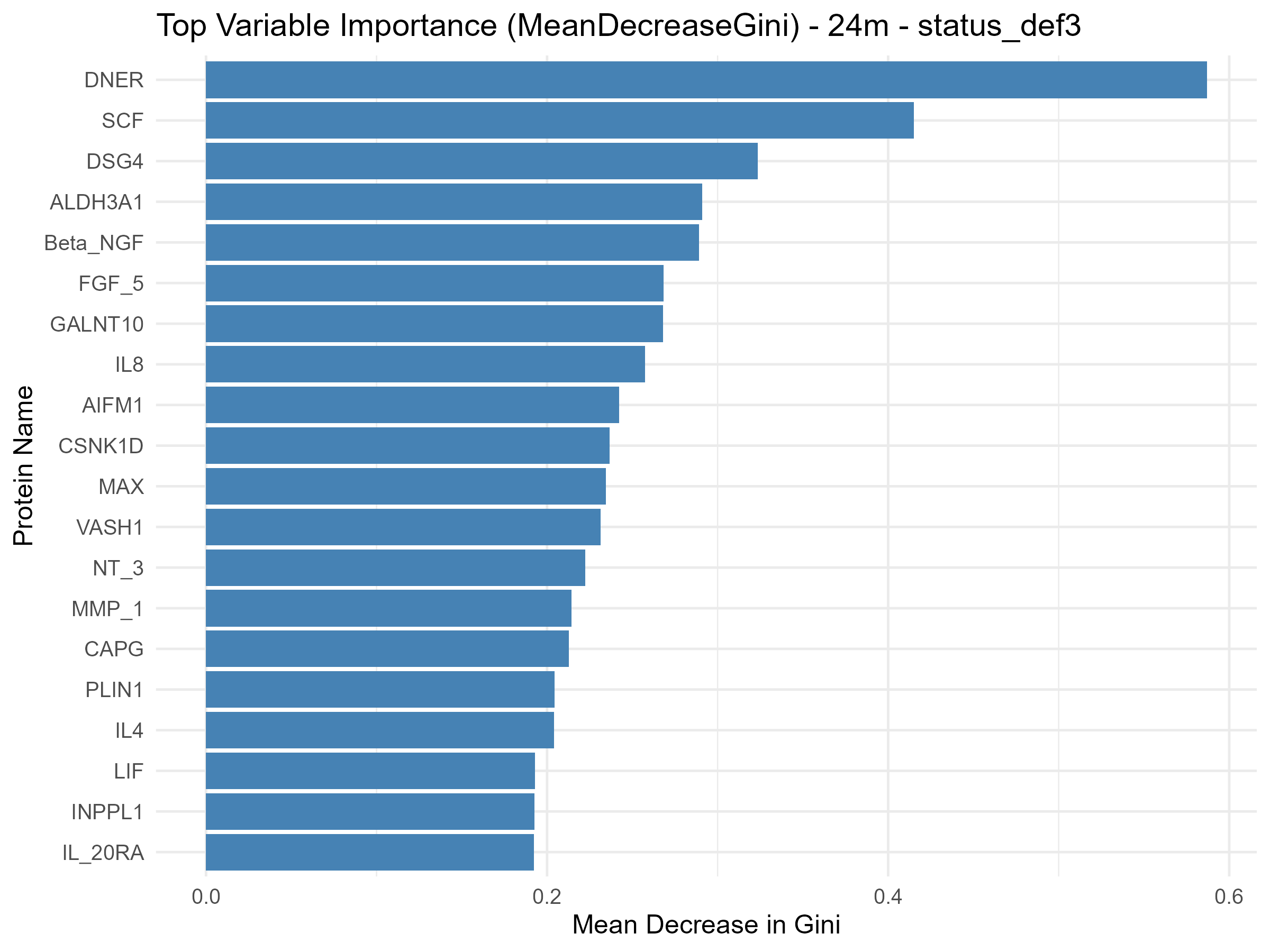

Random forest models evaluated baseline variables associated with lung-function improvement versus all other lung-function categories at 18 months (A, B) and 24 months (C, D). Panels A and C use the standard definition: an absolute increase from baseline of at least 10 percentage points in FVC % predicted and/or at least 15 percentage points in DLCO % predicted. Panels B and D use the less stringent definition: an absolute increase of at least 5 percentage points in FVC % predicted and/or at least 10 percentage points in DLCO % predicted. Bars show mean decrease in Gini impurity (MeanDecreaseGini), a relative variable-importance measure; higher values indicate greater contribution to classification within the corresponding model but do not indicate the direction of association, effect size, or statistical significance. Analyses were exploratory.

**Tables**

**Supplementary Table S1. Protein nomenclature.**

| Protein Name | Uniprot ID | OlinkID | Gene ID | Entry Name |
| --- | --- | --- | --- | --- |
| ADA | P00813 | OID00560 | 100 | ADA |
| ADGRG1 | Q9Y653 | OID01040 | 9289 | AGRG1 |
| AGR2 | O95994 | OID01045 | 10551 | AGR2 |
| AIFM1 | O95831 | OID01048 | 9131 | AIFM1 |
| ALDH3A1 | P30838 | OID01034 | 218 | AL3A1 |
| AMN | Q9BXJ7 | OID01037 | 81693 | AMNLS |
| ARTN | Q5T4W7 | OID00526 | 9048 | ARTN |
| ATP6AP2 | O75787 | OID01101 | 10159 | RENR |
| AXIN1 | O15169 | OID00487 | 8312 | AXIN1 |
| BAMBI | Q13145 | OID01049 | 25805 | BAMBI |
| BANK1 | Q8NDB2 | OID01085 | 55024 | BANK1 |
| Beta-NGF | P01138 | OID00519 | 4803 | NGF |
| BID | P55957 | OID01051 | 637 | BID |
| BTC | P35070 | OID01081 | 685 | BTC |
| CA12 | O43570 | OID01066 | 771 | CAH12 |
| CA14 | Q9ULX7 | OID01061 | 23632 | CAH14 |
| CALCA | P01258 | OID01095 | 796 | CALC |
| CALR | P27797 | OID01119 | 811 | CALR |
| CAPG | P40121 | OID01050 | 822 | CAPG |
| CASP-8 | Q14790 | OID00550 | 841 | CASP8 |
| CCL11 | P51671 | OID00505 | 6356 | CCL11 |
| CCL19 | Q99731 | OID00513 | 6363 | CCL19 |
| CCL20 | P78556 | OID00556 | 6364 | CCL20 |
| CCL23 | P55773 | OID00530 | 6368 | CCL23 |
| CCL25 | O15444 | OID00551 | 6370 | CCL25 |
| CCL28 | Q9NRJ3 | OID00539 | 56477 | CCL28 |
| CCL3 | P10147 | OID00532 | 6348 | CCL3 |
| CCL4 | P13236 | OID00498 | 6351 | CCL4 |
| CD244 | Q9BZW8 | OID00477 | 51744 | CD244 |
| CD40 | P25942 | OID00542 | 958 | TNR5 |
| CD5 | P06127 | OID00531 | 921 | CD5 |
| CD6 | P30203 | OID00499 | 923 | CD6 |
| CD8A | P01732 | OID05124 | 925 | CD8A |
| CDCP1 | Q9H5V8 | OID00476 | 64866 | CDCP1 |
| CES2 | O00748 | OID01055 | 8824 | EST2 |
| CLEC1A | Q8NC01 | OID01031 | 51267 | CLC1A |
| CLSPN | Q9HAW4 | OID01028 | 63967 | CLSPN |
| CNTN2 | Q02246 | OID01108 | 6900 | CNTN2 |
| CRH | P06850 | OID01096 | 1392 | CRF |
| CSF-1 | P09603 | OID00562 | 1435 | CSF1 |
| CSNK1D | P48730 | OID01035 | 1453 | KC1D |
| CST5 | P28325 | OID00491 | 1473 | CYTD |
| CX3CL1 | P78423 | OID00552 | 6376 | X3CL1 |
| CXCL1 | P09341 | OID00496 | 2919 | GROA |
| CXCL10 | P02778 | OID00535 | 3627 | CXL10 |
| CXCL11 | O14625 | OID00486 | 6373 | CXL11 |
| CXCL5 | P42830 | OID00520 | 6374 | CXCL5 |
| CXCL6 | P80162 | OID00534 | 6372 | CXCL6 |
| CXCL9 | Q07325 | OID00490 | 4283 | CXCL9 |
| DNER | Q8NFT8 | OID01213 | 92737 | DNER |
| DPP6 | P42658 | OID01047 | 1804 | DPP6 |
| DSG4 | Q86SJ6 | OID01029 | 147409 | DSG4 |
| EDIL3 | O43854 | OID01064 | 10085 | EDIL3 |
| EGFL7 | Q9UHF1 | OID01114 | 51162 | EGFL7 |
| EN-RAGE | P80511 | OID00541 | 6283 | S10AC |
| ENAH | Q8N8S7 | OID01042 | 55740 | ENAH |
| ENTPD2 | Q9Y5L3 | OID01071 | 954 | ENTP2 |
| ENTPD6 | O75354 | OID01089 | 955 | ENTP6 |
| EPO | P01588 | OID01043 | 2056 | EPO |
| ERBB2IP | Q96RT1 | OID01077 | 55914 | ERBIN |
| FABP9 | Q0Z7S8 | OID01115 | 646480 | FABP9 |
| FES | P07332 | OID01059 | 2242 | FES |
| FGF-19 | O95750 | OID00545 | 9965 | FGF19 |
| FGF-21 | Q9NSA1 | OID00512 | 26291 | FGF21 |
| FGF-23 | Q9GZV9 | OID00507 | 8074 | FGF23 |
| FGF-5 | P12034 | OID00509 | 2250 | FGF5 |
| FGR | P09769 | OID01103 | 2268 | FGR |
| FKBP1B | P68106 | OID01107 | 2281 | FKB1B |
| Flt3L | P49771 | OID00533 | 2323 | FLT3L |
| FOSB | P53539 | OID01118 | 2354 | FOSB |
| FOXO1 | Q12778 | OID01065 | 2308 | FOXO1 |
| GALNT10 | Q86SR1 | OID01080 | 55568 | GLT10 |
| GDNF | P39905 | OID00475 | 2668 | GDNF |
| HGF | P14210 | OID00522 | 3082 | HGF |
| HPGDS | O60760 | OID01105 | 27306 | HPGDS |
| IFN-gamma | P01579 | OID05547 | 3458 | IFNG |
| IL-1-alpha | P01583 | OID00493 | 3552 | IL1A |
| IL-10RA | Q13651 | OID00508 | 3587 | I10R1 |
| IL-10RB | Q08334 | OID00515 | 3588 | I10R2 |
| IL-12B | P29460 | OID00523 | 3593 | IL12B |
| IL-15RA | Q13261 | OID00514 | 3601 | I15RA |
| IL-17A | Q16552 | OID00485 | 3605 | IL17 |
| IL-17C | Q9P0M4 | OID00483 | 27189 | IL17C |
| IL-18R1 | Q13478 | OID00517 | 8809 | IL18R |
| IL-20 | Q9NYY1 | OID00537 | 50604 | IL20 |
| IL-20RA | Q9UHF4 | OID00489 | 53832 | I20RA |
| IL-22-RA1 | Q8N6P7 | OID00516 | 58985 | I22R1 |
| IL-24 | Q13007 | OID00524 | 11009 | IL24 |
| IL-2RB | P14784 | OID00492 | 3560 | IL2RB |
| IL10 | P22301 | OID00528 | 3586 | IL10 |
| IL13 | P35225 | OID00525 | 3596 | IL13 |
| IL18 | Q14116 | OID00501 | 3606 | IL18 |
| IL2 | P60568 | OID00495 | 3558 | IL2 |
| IL33 | O95760 | OID00543 | 90865 | IL33 |
| IL4 | P05112 | OID00546 | 3565 | IL4 |
| IL5 | P05113 | OID00559 | 3567 | IL5 |
| IL6 | P05231 | OID00482 | 3569 | IL6 |
| IL7 | P13232 | OID00478 | 3574 | IL7 |
| IL8 | P10145 | OID00471 | 3576 | IL8 |
| INPPL1 | O15357 | OID01056 | 3636 | SHIP2 |
| ITGB1BP1 | O14713 | OID01091 | 9270 | ITBP1 |
| KIM1 | Q96D42 | OID01075 | 26762 | HAVR1 |
| KIR3DL1 | P43629 | OID05125 | 3811 | KI3L1 |
| LAP-TGF-beta-1 | P01137 | OID00480 | 7040 | TGFB1 |
| LAT2 | Q9GZY6 | OID01074 | 7462 | NTAL |
| LHB | P01229 | OID01117 | 3972 | LSHB |
| LIF | P15018 | OID00547 | 3976 | LIF |
| LIF-R | P42702 | OID00511 | 3977 | LIFR |
| LRP1 | Q07954 | OID01030 | 4035 | LRP1 |
| LTA4H | P09960 | OID01063 | 4048 | LKHA4 |
| MAEA | Q7L5Y9 | OID01079 | 10296 | MAEA |
| MAGED1 | Q9Y5V3 | OID01083 | 9500 | MAGD1 |
| MAP4K5 | Q9Y4K4 | OID01090 | 11183 | M4K5 |
| MAX | P61244 | OID01088 | 4149 | MAX |
| MCP-1 | P13500 | OID00484 | 6347 | CCL2 |
| MCP-2 | P80075 | OID00549 | 6355 | CCL8 |
| MCP-3 | P80098 | OID00474 | 6354 | CCL7 |
| MCP-4 | Q99616 | OID00504 | 6357 | CCL13 |
| METAP1 | P53582 | OID01097 | 23173 | MAP11 |
| MMP-1 | P03956 | OID00510 | 4312 | MMP1 |
| MMP-10 | P09238 | OID00527 | 4319 | MMP10 |
| MVK | Q03426 | OID01039 | 4598 | KIME |
| NBN | O60934 | OID01087 | 4683 | NBN |
| NCF2 | P19878 | OID01102 | 4688 | NCF2 |
| NOS3 | P29474 | OID01053 | 4846 | NOS3 |
| NPPC | P23582 | OID01084 | 4880 | ANFC |
| NRTN | Q99748 | OID00548 | 4902 | NRTN |
| NT-3 | P20783 | OID00554 | 4908 | NTF3 |
| NUB1 | Q9Y5A7 | OID01092 | 51667 | NUB1 |
| NUCB2 | P80303 | OID01052 | 4925 | NUCB2 |
| OPG | O00300 | OID00479 | 4982 | TR11B |
| OSM | P13725 | OID00494 | 5008 | ONCM |
| PD-L1 | Q9NZQ7 | OID00518 | 29126 | PD1L1 |
| PDCD1 | Q15116 | OID01098 | 5133 | PDCD1 |
| PDGFC | Q9NRA1 | OID01044 | 56034 | PDGFC |
| PDP1 | Q9P0J1 | OID01072 | 54704 | PDP1 |
| PGF | P49763 | OID01493 | 5228 | PLGF |
| PLIN1 | O60240 | OID01067 | 5346 | PLIN1 |
| PLXDC1 | Q8IUK5 | OID01086 | 57125 | PLDX1 |
| PON2 | Q15165 | OID01082 | 5445 | PON2 |
| PPM1B | O75688 | OID01069 | 5495 | PPM1B |
| PRKAB1 | Q9Y478 | OID01046 | 5564 | AAKB1 |
| PRKRA | O75569 | OID01068 | 8575 | PRKRA |
| PSMA1 | P25786 | OID01104 | 5682 | PSA1 |
| PTK7 | Q13308 | OID01112 | 5754 | PTK7 |
| PTN | P21246 | OID01099 | 5764 | PTN |
| PTPRJ | Q12913 | OID01110 | 5795 | PTPRJ |
| PVALB | P20472 | OID01113 | 5816 | PRVA |
| PXN | P49023 | OID01032 | 5829 | PAXI |
| RARRES1 | P49788 | OID01093 | 5918 | TIG1 |
| RASA1 | P20936 | OID01078 | 5921 | RASA1 |
| RASSF2 | P50749 | OID01094 | 9770 | RASF2 |
| RCOR1 | Q9UKL0 | OID01062 | 23186 | RCOR1 |
| RRM2B | Q7LG56 | OID01111 | 50484 | RIR2B |
| SCF | P21583 | OID00500 | 4254 | SCF |
| SERPINA9 | Q86WD7 | OID01100 | 327657 | SPA9 |
| SIRT2 | Q8IXJ6 | OID00538 | 22933 | SIR2 |
| SIRT5 | Q9NXA8 | OID01057 | 23408 | SIR5 |
| SLAMF1 | Q13291 | OID00502 | 6504 | SLAF1 |
| SMAD1 | Q15797 | OID01033 | 4086 | SMAD1 |
| ST1A1 | P50225 | OID00557 | 6817 | ST1A1 |
| ST3GAL1 | Q11201 | OID01058 | 6482 | SIA4A |
| STAMBP | O95630 | OID00558 | 10617 | STABP |
| STX8 | Q9UNK0 | OID01076 | 9482 | STX8 |
| STXBP3 | O00186 | OID01106 | 6814 | STXB3 |
| TGF-alpha | P01135 | OID00503 | 7039 | TGFA |
| TIGAR | Q9NQ88 | OID01041 | 57103 | TIGAR |
| TMPRSS15 | P98073 | OID01116 | 5651 | ENTK |
| TNF | P01375 | OID05548 | 7124 | TNFA |
| TNFB | P01374 | OID00561 | 4049 | TNFB |
| TNFRSF9 | Q07011 | OID00553 | 3604 | TNR9 |
| TNFSF14 | O43557 | OID00506 | 8740 | TNF14 |
| TNNI3 | P19429 | OID01054 | 7137 | TNNI3 |
| TOP2B | Q02880 | OID01036 | 7155 | TOP2B |
| TRAIL | P50591 | OID00488 | 8743 | TNF10 |
| TRANCE | O14788 | OID00521 | 8600 | TNF11 |
| TSLP | Q969D9 | OID00497 | 85480 | TSLP |
| TWEAK | O43508 | OID00555 | 8742 | TNF12 |
| uPA | P00749 | OID00481 | 5328 | UROK |
| VASH1 | Q7L8A9 | OID01073 | 22846 | VASH1 |
| VEGFA | P15692 | OID00472 | 7422 | VEGFA |
| WAS | P42768 | OID01109 | 7454 | WASP |
| X4E-BP1 | Q13541 | OID00536 | 1978 | 4EBP1 |
| YES1 | P07947 | OID01070 | 7525 | YES |

Supplementary Table S2. Full baseline clinical characteristics stratified by autoantibody group

| **Characteristic** | **Overall**  N = 225 | **Jo-1**  N = 126 | **MDA5**  N = 39 | **PL-12**  N = 16 | **PL-7**  N = 15 | **EJ**  N = 3 | **Mi-2**  N = 11 | **NXP2**  N = 15 |
| --- | --- | --- | --- | --- | --- | --- | --- | --- |
| **Demographics** | | | | | | | | |
| Age at blood draw^1^ | 49.7 (40.4, 55.5) | 50.7 (39.4, 57.3) | 50.0 (42.6, 55.0) | 41.9 (34.6, 48.2) | 50.0 (42.7, 54.0) | 59.2 (54.5, 71.1) | 49.9 (47.8, 60.4) | 47.6 (39.8, 49.8) |
| Age at myositis diagnosis^1^ | 49.3 (38.7, 54.5) | 49.3 (35.6, 54.7) | 50.2 (43.0, 54.7) | 41.5 (33.4, 51.2) | 47.4 (40.2, 54.0) | 55.2 (53.7, 71.1) | 48.4 (46.0, 60.2) | 44.6 (36.1, 54.4) |
| Age at symptom onset^1^ | 46.3 (37.1, 53.3) | 46.3 (34.3, 53.6) | 48.9 (42.2, 53.1) | 40.0 (31.2, 44.4) | 46.0 (38.8, 50.6) | 50.8 (48.7, 68.4) | 48.1 (45.7, 59.8) | 47.0 (39.5, 49.7) |
| Age at respiratory symptom onset^1^ | 49.3 (40.7, 54.8) | 49.9 (41.7, 55.6) | 50.6 (43.0, 55.1) | 40.7 (37.9, 44.4) | 47.2 (38.7, 51.7) | 55.2 (53.4, 68.4) | NA (NA, NA) | NA (NA, NA) |
| Female^2^ | 158 (70%) | 90 (71%) | 26 (67%) | 14 (88%) | 9 (60%) | 1 (33%) | 6 (55%) | 12 (80%) |
| White^2^ | 139 (62%) | 91 (72%) | 26 (67%) | 0 (0%) | 5 (33%) | 1 (33%) | 7 (64%) | 9 (60%) |
| African American^2^ | 59 (26%) | 26 (21%) | 7 (18%) | 14 (88%) | 8 (53%) | 1 (33%) | 1 (9.1%) | 2 (13%) |
| Other race^2^ | 27 (12%) | 9 (7.1%) | 6 (15%) | 2 (13%) | 2 (13%) | 1 (33%) | 3 (27%) | 4 (27%) |
| **Clinical phenotype and outcome** | | | | | | | | |
| ILD present^2^ | 172 (77%) | 110 (89%) | 30 (77%) | 16 (100%) | 13 (87%) | 3 (100%) | 0 (0%) | 0 (0%) |
| UIP pattern on CT^3^ | 18 (11%) | 10 (9.5%) | 2 (7.1%) | 2 (13%) | 3 (23%) | 1 (33%) | 0 (NA%) | 0 (NA%) |
| NSIP/OP/GGO pattern on CT^3^ | 95 (58%) | 60 (57%) | 16 (57%) | 8 (53%) | 9 (69%) | 2 (67%) | 0 (NA%) | 0 (NA%) |
| ILD severity^3^ |  |  |  |  |  |  |  |  |
| mild | 76 (58%) | 54 (68%) | 14 (58%) | 4 (29%) | 2 (20%) | 2 (67%) | 0 (NA%) | 0 (NA%) |
| moderate | 29 (22%) | 19 (24%) | 2 (8.3%) | 3 (21%) | 5 (50%) | 0 (0%) | 0 (NA%) | 0 (NA%) |
| severe | 26 (20%) | 7 (8.8%) | 8 (33%) | 7 (50%) | 3 (30%) | 1 (33%) | 0 (NA%) | 0 (NA%) |
| Myositis subtype^2^ |  |  |  |  |  |  |  |  |
| Dermatomyositis | 127 (69%) | 58 (59%) | 28 (85%) | 10 (83%) | 7 (54%) | 1 (33%) | 10 (100%) | 13 (87%) |
| Polymyositis | 53 (29%) | 41 (41%) | 0 (0%) | 2 (17%) | 6 (46%) | 2 (67%) | 0 (0%) | 2 (13%) |
| Amyopathic dermatomyositis | 5 (2.7%) | 0 (0%) | 5 (15%) | 0 (0%) | 0 (0%) | 0 (0%) | 0 (0%) | 0 (0%) |
| Proximal weakness^2^ | 157 (89%) | 97 (93%) | 23 (70%) | 12 (92%) | 12 (86%) | 3 (100%) | 5 (100%) | 5 (100%) |
| Distal weakness^2^ | 30 (22%) | 14 (19%) | 6 (21%) | 2 (18%) | 2 (25%) | 0 (0%) | 2 (40%) | 4 (80%) |
| **Mortality** | | | | | | | | |
| Deceased during follow-up^2^ | 43 (19%) | 25 (20%) | 5 (13%) | 6 (38%) | 5 (33%) | 1 (33%) | 1 (9.1%) | 0 (0%) |
| **Pulmonary function** | | | | | | | | |
| PFT restriction severity^3^ |  |  |  |  |  |  |  |  |
| none | 34 (24%) | 22 (25%) | 8 (32%) | 0 (0%) | 0 (0%) | 0 (0%) | 2 (100%) | 2 (100%) |
| mild | 22 (15%) | 18 (20%) | 4 (16%) | 0 (0%) | 0 (0%) | 0 (0%) | 0 (0%) | 0 (0%) |
| moderate | 35 (24%) | 16 (18%) | 8 (32%) | 5 (42%) | 5 (45%) | 1 (33%) | 0 (0%) | 0 (0%) |
| moderately severe | 19 (13%) | 12 (14%) | 3 (12%) | 2 (17%) | 1 (9.1%) | 1 (33%) | 0 (0%) | 0 (0%) |
| severe | 33 (23%) | 20 (23%) | 2 (8.0%) | 5 (42%) | 5 (45%) | 1 (33%) | 0 (0%) | 0 (0%) |
| FVC % predicted^3^ | 70.3 (52.1, 80.1) | 72.2 (58.1, 81.2) | 75.1 (61.1, 86.1) | 55.6 (48.6, 67.9) | 59.1 (50.5, 69.7) | 53.6 (50.3, 76.0) | 93.4 (88.3, 98.4) | 71.8 (46.0, 102.9) |
| FEV1 % predicted^3^ | 71.0 (56.0, 82.9) | 72.5 (58.1, 83.0) | 74.5 (62.6, 83.6) | 65.0 (35.0, 70.1) | 59.6 (43.4, 70.0) | 59.9 (56.3, 78.0) | 91.7 (90.7, 92.7) | 69.2 (44.5, 99.7) |
| TLC % predicted^3^ | 65.9 (52.0, 79.0) | 67.9 (51.3, 79.5) | 68.0 (63.5, 86.5) | 56.2 (40.4, 65.9) | 56.8 (47.0, 64.3) | 59.1 (48.3, 61.0) | 82.9 (81.9, 83.8) | 111.1 (96.3, 125.8) |
| DLCO % predicted^3^ | 59.6 (44.0, 82.1) | 60.1 (46.0, 82.3) | 73.1 (50.6, 83.3) | 38.5 (36.4, 56.1) | 41.1 (34.7, 51.0) | 55.7 (49.0, 62.4) | 99.2 (92.1, 106.2) | 110.6 (105.3, 115.8) |
| **Laboratory features** | | | | | | | | |
| ALT^1^ | 36.0 (24.0, 66.0) | 37.5 (25.0, 72.0) | 39.0 (20.0, 74.0) | 24.0 (14.0, 34.0) | 37.0 (34.0, 57.0) | 26.5 (23.0, 30.0) | 143.0 (143.0, 143.0) | 23.0 (23.0, 23.0) |
| AST^1^ | 31.0 (23.0, 75.0) | 30.5 (23.0, 77.0) | 36.5 (25.0, 77.0) | 23.0 (16.0, 27.0) | 32.0 (28.5, 41.0) | 22.5 (19.0, 26.0) | 79.0 (76.0, 82.0) | 18.0 (18.0, 18.0) |
| CK^1^ | 166.0 (68.0, 741.0) | 261.0 (100.0, 1,182.0) | 56.0 (38.0, 101.0) | 136.0 (54.0, 260.5) | 424.5 (109.0, 683.0) | 189.0 (111.0, 197.0) | 618.0 (599.0, 1,600.0) | 109.0 (30.0, 149.0) |
| CRP^1^ | 1.0 (0.3, 3.5) | 0.9 (0.3, 2.7) | 0.7 (0.1, 1.9) | 7.3 (0.5, 9.2) | 2.6 (1.1, 5.2) | NA (NA, NA) | 0.2 (0.1, 0.3) | 1.3 (0.2, 3.5) |
| ESR^1^ | 28.0 (11.0, 52.0) | 20.0 (9.0, 44.0) | 39.5 (27.0, 48.0) | 61.5 (48.0, 78.0) | 35.0 (21.0, 67.0) | 37.5 (28.0, 47.0) | NA (NA, NA) | 42.0 (7.0, 52.0) |
| **Immunosuppressive medication use** | | | | | | | | |
| No medication records available^2^ | 24 (11%) | 15 (12%) | 2 (5.1%) | 2 (13%) | 2 (13%) | 0 (0%) | 2 (18%) | 1 (6.7%) |
| No immunosuppression^2^ | 7 (3.5%) | 5 (4.5%) | 1 (2.7%) | 1 (7.1%) | 0 (0%) | 0 (0%) | 0 (0%) | 0 (0%) |
| Prednisone^2^ | 168 (84%) | 92 (83%) | 33 (89%) | 11 (79%) | 9 (69%) | 3 (100%) | 8 (89%) | 12 (86%) |
| Mycophenolate mofetil^2^ | 56 (28%) | 25 (23%) | 16 (43%) | 2 (14%) | 6 (46%) | 2 (67%) | 2 (22%) | 3 (21%) |
| IVIG^2^ | 37 (18%) | 16 (14%) | 8 (22%) | 3 (21%) | 1 (7.7%) | 1 (33%) | 2 (22%) | 6 (43%) |
| Azathioprine^2^ | 75 (37%) | 46 (41%) | 13 (35%) | 4 (29%) | 7 (54%) | 1 (33%) | 1 (11%) | 3 (21%) |
| Methotrexate^2^ | 48 (24%) | 30 (27%) | 9 (24%) | 1 (7.1%) | 0 (0%) | 0 (0%) | 4 (44%) | 4 (29%) |
| Rituximab^2^ | 14 (7.0%) | 10 (9.0%) | 0 (0%) | 2 (14%) | 1 (7.7%) | 0 (0%) | 1 (11%) | 0 (0%) |
| Cyclophosphamide^2^ | 9 (4.5%) | 5 (4.5%) | 3 (8.1%) | 1 (7.1%) | 0 (0%) | 0 (0%) | 0 (0%) | 0 (0%) |
| Tacrolimus^2^ | 5 (2.5%) | 1 (0.9%) | 2 (5.4%) | 2 (14%) | 0 (0%) | 0 (0%) | 0 (0%) | 0 (0%) |
| Methylprednisolone^2^ | 12 (6.0%) | 6 (5.4%) | 1 (2.7%) | 3 (21%) | 1 (7.7%) | 0 (0%) | 0 (0%) | 1 (7.1%) |
| Cyclosporine^2^ | 1 (0.5%) | 1 (0.9%) | 0 (0%) | 0 (0%) | 0 (0%) | 0 (0%) | 0 (0%) | 0 (0%) |
| Anakinra^2^ | 1 (0.5%) | 1 (0.9%) | 0 (0%) | 0 (0%) | 0 (0%) | 0 (0%) | 0 (0%) | 0 (0%) |
| Infliximab^2^ | 3 (1.5%) | 3 (2.7%) | 0 (0%) | 0 (0%) | 0 (0%) | 0 (0%) | 0 (0%) | 0 (0%) |
| Etanercept^2^ | 3 (1.5%) | 2 (1.8%) | 1 (2.7%) | 0 (0%) | 0 (0%) | 0 (0%) | 0 (0%) | 0 (0%) |
| Adalimumab^2^ | 1 (0.5%) | 1 (0.9%) | 0 (0%) | 0 (0%) | 0 (0%) | 0 (0%) | 0 (0%) | 0 (0%) |
| **Immunosuppressive combination** | | | | | | | | |
| Prednisone only | 29 (15%) | 12 (11%) | 6 (17%) | 4 (31%) | 1 (7.7%) | 0 (0%) | 3 (33%) | 3 (21%) |
| No prednisone + 1 immunosuppressive | 16 (8.2%) | 7 (6.6%) | 2 (5.6%) | 2 (15%) | 3 (23%) | 0 (0%) | 1 (11%) | 1 (7.1%) |
| No prednisone + 2 immunosuppressives | 4 (2.1%) | 2 (1.9%) | 1 (2.8%) | 0 (0%) | 0 (0%) | 0 (0%) | 0 (0%) | 1 (7.1%) |
| No prednisone + ≥3 immunosuppressives | 1 (0.5%) | 1 (0.9%) | 0 (0%) | 0 (0%) | 0 (0%) | 0 (0%) | 0 (0%) | 0 (0%) |
| Prednisone + 1 immunosuppressive | 90 (46%) | 55 (52%) | 13 (36%) | 3 (23%) | 8 (62%) | 2 (67%) | 3 (33%) | 6 (43%) |
| Prednisone + 2 immunosuppressives | 34 (18%) | 20 (19%) | 8 (22%) | 2 (15%) | 0 (0%) | 1 (33%) | 1 (11%) | 2 (14%) |
| Prednisone + ≥3 immunosuppressives | 20 (10%) | 9 (8.5%) | 6 (17%) | 2 (15%) | 1 (7.7%) | 0 (0%) | 1 (11%) | 1 (7.1%) |

^1^Median (Q1, Q3).

^2^n (%).

^3^Missing values in the Overall IIM cohort for CT/PFT variables: CT pattern: 61, ILD severity: 94, PFT restriction grade: 94, FVC: 74, FEV1: 88, TLC: 94, DLCO: 89.

PFT values are shown as percent predicted. Abbreviations: ALT, alanine aminotransferase; AST, aspartate aminotransferase; CK, creatine kinase; CRP, C-reactive protein; DLCO, diffusing capacity for carbon monoxide; FEV1, forced expiratory volume in 1 second; FVC, forced vital capacity; ILD, interstitial lung disease; IVIG, intravenous immunoglobulin; NSIP, nonspecific interstitial pneumonia; OP, organizing pneumonia; GGO, ground-glass opacity; TLC, total lung capacity; UIP, usual interstitial pneumonia.

The lower-ILD-risk group included Mi-2, NXP2, and one anti-TIF1γ-positive patient; the anti-TIF1γ patient is not shown separately because n=1.

Denominators vary because of missing data; percentages are calculated among participants with available data for each variable.

Supplementary Table S3. Full clinical characteristics and outcomes stratified by proteomic cluster

| **Characteristic** | **Cluster 1**  N = 45 | **Cluster 2**  N = 70 | **Cluster 3**  N = 50 | **Cluster 4**  N = 39 |
| --- | --- | --- | --- | --- |
| **Demographics** | | | | |
| Age at blood draw^1^ | 49.9 (38.3, 56.0) | 48.6 (41.2, 54.7) | 47.9 (41.4, 57.5) | 50.2 (40.0, 55.1) |
| Age at myositis diagnosis^1^ | 46.4 (34.0, 52.7) | 49.2 (40.2, 54.5) | 46.6 (35.5, 54.9) | 49.3 (38.6, 54.3) |
| Age at symptom onset^1^ | 45.6 (36.4, 53.3) | 44.4 (37.7, 52.8) | 46.2 (34.0, 54.6) | 44.1 (33.9, 52.7) |
| Age at respiratory symptom onset^1^ | 49.4 (40.7, 53.5) | 49.2 (42.1, 54.0) | 46.9 (38.8, 56.9) | 46.5 (39.3, 53.0) |
| Female^2^ | 35 (78%) | 55 (79%) | 33 (66%) | 25 (64%) |
| White^2^ | 28 (62%) | 45 (64%) | 29 (58%) | 27 (69%) |
| African American^2^ | 13 (29%) | 16 (23%) | 17 (34%) | 8 (21%) |
| Other race^2^ | 4 (8.9%) | 9 (13%) | 4 (8.0%) | 4 (10%) |
| **Clinical phenotype and outcome** | | | | |
| ILD present^2^ | 32 (71%) | 46 (66%) | 45 (92%) | 33 (87%) |
| UIP pattern on CT^3^ | 2 (6.9%) | 3 (6.7%) | 8 (18%) | 1 (3.1%) |
| NSIP/OP/GGO pattern on CT^3^ | 19 (66%) | 23 (51%) | 27 (61%) | 19 (59%) |
| ILD severity^3^ |  |  |  |  |
| Mild | 13 (57%) | 22 (61%) | 19 (51%) | 17 (71%) |
| Moderate | 4 (17%) | 8 (22%) | 12 (32%) | 3 (13%) |
| Severe | 6 (26%) | 6 (17%) | 6 (16%) | 4 (17%) |
| Myositis subtype^2^ |  |  |  |  |
| Dermatomyositis | 26 (70%) | 40 (71%) | 24 (60%) | 23 (68%) |
| Polymyositis | 9 (24%) | 16 (29%) | 15 (38%) | 9 (26%) |
| Amyopathic dermatomyositis | 2 (5.4%) | 0 (0%) | 1 (2.5%) | 2 (5.9%) |
| Proximal weakness^2^ | 30 (83%) | 44 (96%) | 39 (87%) | 29 (85%) |
| Distal weakness^2^ | 5 (18%) | 10 (26%) | 4 (12%) | 8 (36%) |
| **Mortality** | | | | |
| Deceased during follow-up^2^ | 4 (8.9%) | 10 (14%) | 17 (34%) | 8 (21%) |
| **Pulmonary function** | | | | |
| PFT restriction severity^3^ |  |  |  |  |
| None | 4 (14%) | 12 (32%) | 6 (17%) | 10 (37%) |
| Mild | 7 (25%) | 4 (11%) | 2 (5.7%) | 5 (19%) |
| Moderate | 9 (32%) | 7 (18%) | 8 (23%) | 6 (22%) |
| Moderately severe | 2 (7.1%) | 5 (13%) | 9 (26%) | 1 (3.7%) |
| Severe | 6 (21%) | 10 (26%) | 10 (29%) | 5 (19%) |
| FVC % predicted^3^ | 68.2 (50.4, 77.1) | 75.3 (65.7, 87.3) | 60.0 (50.4, 67.8) | 75.6 (55.5, 85.6) |
| FEV1 % predicted^3^ | 71.9 (55.6, 78.4) | 74.0 (69.5, 85.0) | 60.0 (52.0, 70.0) | 72.1 (63.2, 83.1) |
| TLC % predicted^3^ | 66.7 (58.7, 78.1) | 65.9 (46.1, 87.0) | 58.8 (48.0, 69.0) | 71.3 (61.0, 84.0) |
| DLCO % predicted^3^ | 59.6 (42.0, 81.4) | 62.7 (47.2, 76.0) | 52.1 (42.9, 76.1) | 58.7 (46.0, 82.1) |
| **Laboratory features** | | | | |
| ALT^1^ | 51.0 (29.5, 73.0) | 38.0 (24.0, 66.0) | 36.0 (26.0, 65.0) | 31.0 (19.0, 37.0) |
| AST^1^ | 38.5 (24.5, 80.0) | 28.0 (23.0, 63.5) | 33.0 (23.0, 77.0) | 28.0 (20.0, 68.0) |
| CK^1^ | 122.0 (66.0, 377.0) | 165.0 (58.0, 963.0) | 236.5 (97.5, 616.0) | 182.0 (84.0, 1,606.0) |
| CRP^1^ | 1.5 (0.3, 2.4) | 0.8 (0.4, 2.5) | 2.4 (0.5, 6.6) | 0.7 (0.3, 1.9) |
| ESR^1^ | 25.5 (10.0, 44.0) | 23.0 (11.0, 50.0) | 31.5 (14.0, 63.0) | 32.0 (8.5, 47.0) |
| **Immunosuppressive medication use** | | | | |
| No medication records available^2^ | 2 (4.4%) | 9 (13%) | 9 (18%) | 3 (7.7%) |
| No immunosuppression^2^ | 1 (2.3%) | 2 (3.3%) | 2 (4.9%) | 1 (2.8%) |
| Prednisone^2^ | 38 (88%) | 45 (74%) | 36 (88%) | 31 (86%) |
| Mycophenolate mofetil^2^ | 12 (28%) | 16 (26%) | 10 (24%) | 12 (33%) |
| IVIG^2^ | 8 (19%) | 11 (18%) | 5 (12%) | 8 (22%) |
| Azathioprine^2^ | 15 (35%) | 21 (34%) | 16 (39%) | 14 (39%) |
| Methotrexate^2^ | 11 (26%) | 15 (25%) | 13 (32%) | 8 (22%) |
| Rituximab^2^ | 4 (9.3%) | 3 (4.9%) | 2 (4.9%) | 3 (8.3%) |
| Cyclophosphamide^2^ | 3 (7.0%) | 2 (3.3%) | 3 (7.3%) | 1 (2.8%) |
| Tacrolimus^2^ | 3 (7.0%) | 1 (1.6%) | 0 (0%) | 1 (2.8%) |
| Methylprednisolone^2^ | 3 (7.0%) | 4 (6.6%) | 0 (0%) | 4 (11%) |
| Cyclosporine^2^ | 0 (0%) | 0 (0%) | 1 (2.4%) | 0 (0%) |
| Anakinra^2^ | 0 (0%) | 0 (0%) | 1 (2.4%) | 0 (0%) |
| Infliximab^2^ | 0 (0%) | 0 (0%) | 1 (2.4%) | 2 (5.6%) |
| Etanercept^2^ | 1 (2.3%) | 2 (3.3%) | 0 (0%) | 0 (0%) |
| Adalimumab^2^ | 1 (2.3%) | 0 (0%) | 0 (0%) | 0 (0%) |
| **Immunosuppressive combination** | | | | |
| Prednisone only | 5 (12%) | 13 (22%) | 4 (10%) | 3 (8.6%) |
| No prednisone + 1 immunosuppressive | 3 (7.1%) | 9 (15%) | 3 (7.7%) | 1 (2.9%) |
| No prednisone + 2 immunosuppressives | 1 (2.4%) | 3 (5.1%) | 0 (0%) | 0 (0%) |
| No prednisone + ≥3 immunosuppressives | 0 (0%) | 1 (1.7%) | 0 (0%) | 0 (0%) |
| Prednisone + 1 immunosuppressive | 21 (50%) | 20 (34%) | 21 (54%) | 19 (54%) |
| Prednisone + 2 immunosuppressives | 7 (17%) | 8 (14%) | 7 (18%) | 8 (23%) |
| Prednisone + ≥3 immunosuppressives | 5 (12%) | 5 (8.5%) | 4 (10%) | 4 (11%) |

^1^Median (Q1, Q3). ^2^n (%). ^3^Total missing values across proteomic clusters for CT/PFT variables: CT pattern: 51, ILD severity: 81, PFT restriction grade: 85, FVC: 67, FEV1: 81, TLC: 85, DLCO: 80. PFT values are shown as percent predicted. Abbreviations as in Supplementary Table S2.

Denominators vary because of missing data; percentages are calculated among participants with available data for each variable.

**Supplementary Table S4. Individual protein Cox proportional hazards models for all-cause mortality.**

| **Protein Name** | **HR** | **95% CI lower** | **95% CI upper** | ***P* value** |
| --- | --- | --- | --- | --- |
| EPO | 1.41 | 1.15 | 1.72 | 0.001 |
| CCL20 | 1.47 | 1.15 | 1.87 | 0.002 |
| TGF-alpha | 1.64 | 1.19 | 2.27 | 0.003 |
| CXCL1 | 1.76 | 1.22 | 2.55 | 0.003 |
| PDP1 | 4.68 | 1.68 | 13.09 | 0.003 |
| CD6 | 0.59 | 0.42 | 0.84 | 0.003 |
| IL6 | 1.38 | 1.11 | 1.72 | 0.004 |
| CCL28 | 1.68 | 1.18 | 2.38 | 0.004 |
| ST1A1 | 1.96 | 1.24 | 3.12 | 0.004 |
| VEGFA | 1.94 | 1.23 | 3.07 | 0.004 |
| CXCL5 | 1.91 | 1.22 | 2.99 | 0.005 |
| OSM | 1.41 | 1.08 | 1.85 | 0.013 |
| IL-17A | 1.38 | 1.07 | 1.79 | 0.014 |
| HGF | 1.71 | 1.11 | 2.63 | 0.015 |
| IL-2RB | 1.83 | 1.07 | 3.14 | 0.027 |
| CRH | 0.70 | 0.52 | 0.96 | 0.027 |
| CALR | 1.82 | 1.05 | 3.15 | 0.032 |
| TNFSF14 | 1.46 | 1.03 | 2.08 | 0.034 |
| STX8 | 1.45 | 1.02 | 2.07 | 0.036 |
| PRKAB1 | 1.49 | 1.02 | 2.17 | 0.037 |
| IL-24 | 1.62 | 1.03 | 2.56 | 0.038 |
| IL7 | 1.46 | 1.02 | 2.08 | 0.039 |
| IL-17C | 1.46 | 1.01 | 2.10 | 0.044 |
| PDCD1 | 0.67 | 0.46 | 0.99 | 0.045 |
| FGF-23 | 1.36 | 1.01 | 1.83 | 0.046 |
| MMP-10 | 1.447412943 | 1.005800125 | 2.082923014 | 0.046465311 |
| SMAD1 | 1.667704583 | 1.004437603 | 2.768951069 | 0.048028453 |
| SLAMF1 | 1.484185903 | 1.000021244 | 2.202761 | 0.049983466 |

Individual protein-level Cox proportional hazards models were used to evaluate associations between baseline protein expression and all-cause mortality. Each row represents a separate multivariable model adjusted for age, sex, and ILD status, with one protein included as the predictor of interest. Hazard ratios (HRs), 95% confidence intervals, and nominal p-values are shown. Proteins are ordered by ascending p-value. These analyses were used as exploratory protein-level screening and were interpreted together with random-forest variable-importance analyses to prioritize proteins for composite mortality-score construction.
